# Blood Biomarkers of Alzheimer’s Disease Neuropathology, Cognition, and Response to a Multidomain Lifestyle Intervention: Secondary analysis of the FINGER Randomized Controlled Dementia Risk Reduction Trial

**DOI:** 10.64898/2026.10.02.26364486

**Authors:** Anna Rosenberg, Ari Solomon, Anna Matton, Jenni Lehtisalo, Andrea L. Benedet, Francesca Mangialasche, Esko Levälahti, Burak Arslan, Makrina Daniilidou, Markku Peltonen, Ilaria Pola, Juha Rinne, Kübra Tan, Tiia Ngandu, Henrik Zetterberg, Miia Kivipelto

## Abstract

Interventions targeting lifestyle and health behaviors support cognition in older adults at risk of dementia. It remains unclear if the cognitive benefit of such interventions is modified by Alzheimer’s disease (AD)-related neuropathology, and if lifestyle interventions lead to changes in blood AD biomarkers. Here, we analyzed core AD plasma biomarkers (Aβ, p-tau181, p-tau217, p-tau231, NFL, GFAP) in the two-year FINGER randomized controlled multidomain lifestyle trial, which demonstrated cognitive efficacy in an at-risk older general population (N=1260). We found that 85% of participants had p-tau217 below the pathological threshold at baseline (Lumipulse assay). Higher biomarker levels were associated with less favorable cognitive trajectories over two years, but their ability to predict brain amyloid accumulation on PET scans was modest (e.g., p-tau217 AUC 0.72). We found no indication that baseline biomarker levels significantly modified the cognitive benefits of the intervention. Changes in biomarkers were small over two years, with most participants (82%) remaining below the pathological p-tau217 threshold. Biomarker trajectories were similar in the intervention and control groups, suggesting that the cognitive benefits of the two-year lifestyle intervention in at-risk individuals without substantial impairment may primarily involve mechanistic pathways not captured by AD blood biomarkers. These findings provide new evidence to clarify the role of AD blood biomarkers for risk stratification and prediction of response to lifestyle interventions, which is crucial to inform future dementia precision prevention strategies. ClinicalTrials.gov ID NCT01041989.

## INTRODUCTION

Dementia, most commonly caused by Alzheimer’s disease (AD), represents a major global health challenge (1). Its underlying neuropathology unfolds over decades, long before clinical symptoms emerge (2), defining a window for early intervention to prevent cognitive decline. Biomarkers measured in blood now enable a minimally invasive assessment of disease-related neuropathology, including beta-amyloid (Aβ) accumulation, tau phosphorylation, neurodegeneration, and neuroinflammation (3). Plasma-based phosphorylated (p)-tau species, such as tau phosphorylated at threonine 217 (p-tau217) and p-tau231, reflect both Aβ and tau pathologies (4) and begin to increase already in early disease stages (5). These markers, particularly p-tau217, show strong concordance with current gold-standard Aβ measures in cerebrospinal fluid (CSF) and positron emission tomography (PET) (6,7), and are therefore already used in some clinical settings to support AD diagnosis in symptomatic individuals (8). Other markers like neurofilament light chain (NFL) and glial fibrillary acidic protein (GFAP) reflect neurodegeneration and astrocytic reactivity/ neuroinflammation which are AD-related but not AD-specific processes (9). Beyond their concordance with established measures of neuropathology, elevated levels of AD-related blood biomarkers predict e.g., faster longitudinal accumulation of Aβ and tau on PET (10) and dementia-related structural brain changes (11), as well as cognitive decline (12,13) and incident cognitive impairment and dementia (14–17).

While AD-related blood biomarkers show promising diagnostic and prognostic value in symptomatic individuals, evidence regarding their performance and dynamics in cognitively unimpaired individuals and at preclinical disease stages remains more limited. This gap is critical, as effective dementia risk reduction likely requires targeting at-risk individuals before symptom onset. Preventive strategies may include pharmacological approaches, but also interventions addressing the modifiable lifestyle, vascular, and metabolic risk factors of dementia (18), as was highlighted also in the 2026 updated World Health Organization (WHO) guidelines for dementia risk reduction (19). The Finnish Geriatric Intervention Study to Prevent Cognitive Impairment and Disability (FINGER) was among the first large-scale randomized controlled trials (RCT) to show that a two-year multidomain intervention combining dietary counselling, physical activity, cognitive training, and vascular/metabolic risk management improves cognition and reduces the risk of decline in community-dwelling older adults at increased risk of dementia but without substantial cognitive impairment at study entry (20). These findings have since been replicated in large trials in the United States (21) and Latin America (22), and several trials are ongoing worldwide (23).

In this study, we investigated a broad panel of core AD-related blood biomarkers (Aβ42 and 40, p-tau181, p-tau217, p-tau231, GFAP, and NFL) in the FINGER RCT. Using baseline and two-year plasma samples analyzed with two state-of-the-art methods (NULISAseq™ CNS Disease 120 panel and Fujirebio Lumipulse assay for p-tau217), we tested whether baseline blood biomarker levels predicted cognitive response to the multidomain lifestyle intervention, and whether the intervention led to changes in blood biomarkers. While the new blood biomarkers are increasingly used in pharmacological AD RCTs as exploratory endpoints (24,25), our study provides, to our knowledge, the first evidence from a large-scale lifestyle-based dementia risk reduction RCT with demonstrated cognitive efficacy. We also examined associations between blood biomarkers, cognitive performance, and Aβ-PET positivity, extending the currently limited evidence in at-risk older adults.

## METHODS

### Study setting and population

This study is a secondary analysis of the FINGER RCT (ClinicalTrials.gov ID NCT01041989). FINGER investigated the efficacy of a multidomain lifestyle intervention in community-dwelling older adults at risk of dementia, in six centers in Finland (N=1260, mean age 69 years at baseline). FINGER received ethical approval from the Coordinating Ethics Committee of Helsinki and Uusimaa Hospital District, and all participants provided written informed consent. Screening and baseline assessments took place in September 2009–November 2011, and the core two-year RCT ended in February 2014. We performed the analyses of this study in 2025–2026. FINGER protocol (26), baseline population characteristics (27), and main trial results (20) are published elsewhere.

FINGER participants were recruited from previous national population-based surveys, and the main inclusion criteria were age 60–77 years, Cardiovascular Risk Factors, Aging, and Dementia (CAIDE) Risk Score (28) of at least six points, and cognitive performance at the mean level or slightly lower than expected for age. Main exclusion criteria were dementia (previously diagnosed or suspected based on screening assessments), substantial cognitive impairment (Mini-Mental State Examination, MMSE <20), and any conditions affecting safe engagement in the intervention. These criteria selected individuals representative of the general Finnish older population with several dementia risk factors, but without pronounced cognitive impairment (20,27).

Participants were randomized 1:1 to intervention group (intensive multidomain lifestyle program, n=631) or control group (regular health advice, n=629). Computer-generated allocation was performed at each site in blocks of four (two participants randomly allocated to each group) at each site after baseline by the study nurse. Double blinding was pursued as much as possible. The outcome assessors were blinded and not involved in the intervention, and group allocation was not actively disclosed to participants. Primary trial outcome was change in cognition measured with a total composite z-score of a Neuropsychological Test Battery (NTB). Secondary cognitive outcomes included domain-specific NTB z-scores for memory, executive functioning, and processing speed. After two years, the intervention group showed significantly more cognitive improvement than the control group and a decreased risk of cognitive decline (20). The PET sub-study (conducted at baseline) included 46 participants who were scanned with ^11^C-Pittsburgh compound B (PiB)-PET to visually assess Aβ status (29).

### Intervention

All participants received lifestyle information and feedback on their vascular risk factors at baseline. After randomization, participants assigned to control received regular health advice and information about their risk factors and blood test results at the main study visits (at six months, one year and two years), together with advice to contact their health care provider if needed. Participants assigned to intervention received a multidomain program including dietary counselling, exercise, cognitive stimulation, and monitoring and management of vascular and metabolic factors. The intervention, described in detail elsewhere (20,26), was delivered by a multidisciplinary team of professionals, following national and international guidelines.

In brief, the dietary intervention consisted of three individual and six to eight group sessions to tailor participant’s diet. The exercise component included progressive strength training (one to three weekly group sessions) and independent, yet individually planned, aerobic training (two to five weekly sessions). The cognitive intervention included ten group sessions (e.g., educational content) and independent, progressive computer-based cognitive training (in two six-month blocks, training frequency three times per week). Program was designed to train executive processes, memory, and mental speed, with tasks different from the ones included in the outcome assessments. Management of metabolic and vascular risk factors aimed at improving blood pressure, blood lipids, glucose, and body weight, in line with national care guidelines. Six meetings were scheduled for additional measurements and lifestyle guidance. Medications were not prescribed, but participants were advised to contact their health care provider if needed.

### Cognitive assessment

Cognitive assessment (NTB) was conducted by blinded study psychologists at baseline, one year, and two years. The primary outcome of the trial was change in NTB total score reflecting global cognition (the mean of 14 individual tests, at least eight required for calculation). Scores at each timepoint were calculated as z-scores standardized to baseline mean and SD (higher score = better performance). Z-scores for executive functioning, processing speed, and memory domains were secondary outcomes. Abbreviated memory score was also calculated (four tests with more complex processing and longer recall delay). Cognitive decline was defined as a negative z-score change from baseline to two years. Decline in each cognitive outcome was analyzed separately.

### Blood sample collection and analysis

Fasting venous blood samples were collected at baseline, six months, one year, and two years. Separated serum and plasma were frozen immediately and transferred monthly from each site to the laboratory of the Finnish Institute for Health and Welfare (study coordinator) where the samples were aliquoted and stored at −70°C until further analyses. Apolipoprotein E *(APOE)* genotype was determined using standard techniques (30). Estimated glomerular filtration rate (eGFR, indicator of renal function) was calculated in this study based on serum creatinine, sex, and age with the 2021 CKD-EPI equation. AD biomarkers were analyzed in all available banked plasma samples from baseline (n=1225) and two years (n=1105). Samples were shipped in February 2024 and analyzed at the Clinical Neurochemistry Laboratory in Mölndal, Gothenburg, Sweden. The assays performed were the Fujirebio Lumipulse (for p-tau217), clinically validated as previously described (31), and NULISAseq™ CNS Disease 120 Panel (Alamar Biosciences; for Aβ, p-tau species, NFL, and GFAP). The NULISA (NUcleic acid Linked Immuno-Sandwich Assay) provides an ultrasensitive, multiplexed detection and quantification of 124 proteins (32). This panel has shown comparable performance to other established methods in predicting Aβ pathology (33,34).

Participants’ samples were balanced (randomization group allocation and age) across study plates while keeping longitudinal samples of a single individual on the same plate. Sample and data analysis was performed according to kit manufacturer protocols, with the generation of NULISA Protein Quantification (NPQ) values on log2 scale (32,35). Therefore, NPQ units reflect relative protein abundances and differences are interpreted as log2 (fold change), whereas the Lumipulse p-tau217 assay yields absolute concentrations (pg/ml).

### Statistical analysis

Analyses were conducted using Stata, version 18.0. All statistical tests were two-sided with a significance threshold of p<0.05. Estimates are presented with 95% confidence intervals (CIs).

Baseline characteristics were compared between the intervention and control groups using t-tests (continuous variables) and χ² tests (categorical variables). Blood biomarker correlations were assessed using Spearman rank correlation (Spearman’s ρ). The ability of blood biomarkers (continuous levels) to discriminate Aβ PET positivity at baseline was evaluated using logistic regression models, with performance quantified using the area under the receiver operating characteristic curve (AUC) and corresponding 95% CIs.

Cross-sectional baseline associations between participant characteristics and biomarker levels, as well as between biomarker levels and cognition, were examined with linear regression models. Results for biomarker outcomes are reported as estimated differences between two values of the predictor (mean and mean + 1 unit for continuous variables or between category levels for categorical variables). For cognition, effect estimates are expressed as differences in cognitive z-scores corresponding to 1 standard deviation (SD) differences in biomarker levels. Effect modification of baseline biomarker-cognition associations by age, sex, apolipoprotein E (*APOE*) genotype, and vascular risk factor burden (assessed using the validated Finnish FINRISK score, as described in a previous study (36)) were evaluated by including biomarker × characteristic interaction terms in the models. Interaction significance was assessed using likelihood ratio tests.

Associations between baseline biomarker levels and cognitive trajectories over two years were analyzed using linear mixed-effects models with repeated cognitive measurements (baseline, one year, and two years), accounting for within- and between-person variability over time. Time was modeled as a continuous variable (years since baseline), and models included subject-specific random intercepts and slopes. In line with the FINGER main trial approach, analyses were conducted in the modified intention-to-treat population (participants with at least one post-baseline cognitive assessment) (20). Baseline biomarker levels were included as predictors, and models also included time, randomization group, and their interactions (group × time, biomarker × group, biomarker × time, and biomarker × group × time). From these models, we derived estimates for the associations between baseline biomarker levels and cognitive change in the whole cohort, irrespective of randomization (results reported as differences in cognitive change corresponding to 1 SD differences in baseline biomarker levels). The biomarker × group × time interaction was examined to assess whether baseline biomarker levels modified the intervention effects on cognition, i.e., whether the differences in cognitive change between intervention and control groups differed by baseline biomarker levels. This approach aligns with our previous FINGER subgroup analyses (30,36). Here, the biomarker × group × time interaction (the main result) was derived from a model where biomarker was entered as a continuous variable, but for visualization purposes and to facilitate the interpretation of results, estimates of intervention effects on cognition are shown across biomarker strata (below vs. above median levels). Lumipulse p-tau217 analysis was additionally performed by categorizing participants according to established cut-offs (normal <0.22 pg/ml, intermediate 0.22–0.34 pg/ml, pathological >0.34 pg/ml (31,37); intermediate and pathological groups were combined in the analysis). To explore the potential impact of *APOE* genotype on the associations between biomarker levels and intervention effects on cognition, models were extended to include a four-way *APOE* × biomarker × group × time interaction.

We also examined the associations between baseline biomarker levels and risk of cognitive decline over two years, in the whole cohort (irrespective of randomization), using logistic regression. We analyzed biomarkers as continuous variables, and Lumipulse p-tau217 additionally as a categorical variable (normal, intermediate, pathological). Cognitive decline was defined as any decrease in NTB z-score from baseline to two-year follow-up. Decline in each cognitive outcome (global cognition, memory, abbreviated memory, executive functioning, processing speed) was analyzed separately. For Lumipulse p-tau217 status, odds ratios (ORs) are reported relative to the reference category (normal).

Intervention effects on two-year biomarker trajectories were analyzed using linear mixed-effects models with biomarker levels as the outcome and time and randomization group as fixed effects, including their interaction. Analyses were restricted to participants with biomarker measurements available at both baseline and two years.

In all regression analyses, we used marginal prediction methods to derive interpretable effect estimates. Biomarker values were transformed for analyses to reduce skewness and converted back to original scale for presentation of the results. All analyses apart from the Aβ-PET analysis were adjusted for baseline age, sex, years of education, study site, *APOE* genotype (ε4 carrier vs. non-carrier), body mass index (BMI), and estimated glomerular filtration rate (eGFR) as an indicator of renal function. Any missing covariate values were replaced with the mean value, to retain all observations in the analyses. Models with risk of cognitive decline as outcome were additionally adjusted for the baseline cognitive score. Models investigating longitudinal biomarker changes were further adjusted for two-year change in eGFR and its interaction with randomization group, and change in BMI was included as an additional covariate in a sensitivity analysis.

## RESULTS

### Population characteristics

Blood biomarkers were analyzed for all FINGER study participants with available plasma samples (n=1225 out of 1260 randomized at baseline, n=1105 at two years). Baseline characteristics are shown in Table 1. Participants were on average 68.8 (SD 4.7) years old and had 10.0 (SD 3.4) years of education. 46.9% were women, and a third were carriers of the apolipoprotein E (*APOE)* ε4 allele. The intervention and control groups did not significantly differ in any characteristics. Median Lumipulse p-tau217 concentration in the whole cohort was 0.10 [interquartile range 0.08– 0.15]. A total of 85.4% were within the normal range (<0.22 pg/ml). Eight percent had intermediate levels (0.22–0.34 pg/ml) and 6.6% had pathological levels (>0.34 pg/ml) (37).

**Table 1.** Population characteristics.

|  | n with data | All participants (N=1225) | Intervention (n=617) | Control (n=608) | p |
| --- | --- | --- | --- | --- | --- |
| **Demographics** |  |  |  |  |  |
| Age, years | 1225 | 68.8 (4.7) | 69.0 (4.7) | 68.7 (4.7) | 0.25 |
| Women | 1225 | 574 (46.9%) | 282 (45.7%) | 292 (48.0%) | 0.42 |
| Education, years | 1223 | 10.0 (3.4) | 9.9 (3.5) | 10.0 (3.4) | 0.82 |
| **Cognition** |  |  |  |  |  |
| NTB Global cognition | 1224 | -0.01 (0.58) | -0.03 (0.56) | 0.02 (0.59) | 0.14 |
| NTB Memory | 1224 | -0.00 (0.67) | -0.03 (0.68) | 0.02 (0.66) | 0.20 |
| NTB Abbreviated memory | 1203 | -0.00 (0.76) | -0.02 (0.78) | 0.02 (0.74) | 0.32 |
| NTB Executive functioning | 1223 | -0.02 (0.68) | -0.04 (0.66) | 0.00 (0.69) | 0.29 |
| NTB Processing speed | 1224 | -0.00 (0.82) | -0.04 (0.79) | 0.03 (0.85) | 0.19 |
| MMSE | 1221 | 26.7 (2.1) | 26.7 (2.1) | 26.7 (2.1) | 0.77 |
| **APOE, Aβ-PET** |  |  |  |  |  |
| APOE ε4 carrier | 1153 | 384/1153 (33.3%) | 188/581 (32.4%) | 196/572 (34.3%) | 0.49 |
| Positive PiB-PET scan (visual read) | 46 | 19/46 (41.3%) | 7/24 (29.2%) | 12/22 (54.6%) | 0.08 |
| **Clinical characteristics** |  |  |  |  |  |
| Body mass index | 1216 | 28.2 (4.6) | 28.3 (4.5) | 28.1 (4.7) | 0.47 |
| eGFR, ml/min/1.73 m<sup>2</sup> | 1224 | 82.6 (13.5) | 81.8 (13.8) | 83.3 (13.1) | 0.05 |
| Vascular risk | 1201 | 1.5 [1.1–2.3] | 1.5 [1.1–2.3] | 1.5 [1.1–2.2] | 0.77 |
| **AD blood biomarkers** |  |  |  |  |  |
| p-tau217 (Lumipulse), pg/ml | 1221 | 0.10 [0.08-0.15] | 0.10 [0.08-0.15] | 0.10 [0.08-0.15] | 0.41 |
| Normal (<0.22) |  | 1043 (85.4%) | 525 (85.2%) | 518 (85.6%) |  |
| Intermediate (0.22–0.34) |  | 98 (8.0%) | 55 (8.9%) | 43 (7.1%) | 0.33 |
| Pathological (>0.34) |  | 80 (6.6%) | 36 (5.8%) | 44 (7.3%) |  |
| p-tau181, NPQ | 1224 | 12.73 [12.39-13.09] | 12.73 [12.38-13.10] | 12.72 [12.39-13.09] | 0.53 |
| p-tau217, NPQ | 1224 | 10.83 [10.54-11.18] | 10.82 [10.53-11.17] | 10.84 [10.55-11.18] | 0.68 |
| p-tau231, NPQ | 1224 | 12.95 [12.60-13.35] | 12.97 [12.60-13.35] | 12.94 [12.60-13.35] | 0.92 |
| Aβ42, NPQ | 1224 | 12.50 [12.12-12.77] | 12.54 [12.12-12.81] | 12.47 [12.12-12.74] | 0.17 |
| Aβ40, NPQ | 1224 | 12.42 [12.13-12.75] | 12.43 [12.12-12.79] | 12.42 [12.13-12.71] | 0.37 |
| Aβ42/40 | 1224 | 0.02 [-0.31-0.33] | 0.04 [-0.30-0.37] | 0.01 [-0.32-0.31] | 0.70 |
| GFAP, NPQ | 1224 | 14.44 [13.97-14.90] | 14.45 [14.00-14.90] | 14.41 [13.95-14.90] | 0.11 |
| NFL, NPQ | 1224 | 14.33 [14.00-14.68] | 14.37 [14.01-14.70] | 14.30 [13.99-14.66] | 0.55 |

Data are presented as n (%), mean (SD), and median [interquartile range], for all FINGER participants included in the biomarker analysis. P-tau217 was measured with the Lumipulse assay (pg/ml) and the NULISaseq™ CNS Disease Panel 120 (NULISA Protein Quantification unit, NPQ; log2-scale with values reflecting relative protein abundances). Other markers were measured with the NULISaseq™ CNS Disease Panel 120. Aβ42/40: Aβ42 minus Aβ40 due to log2 scale. NTB scores are expressed as standardized z scores, MMSE as total score. Group differences were tested using t-test (for continuous variables) or χ<sup>2</sup> test (for categorical variables), with p-values <0.05 considered statistically significant.
Aβ=beta-amyloid; AD=Alzheimer's disease; APOE=apolipoprotein E; eGFR=estimated glomerular filtration rate; GFAP=glial fibrillary acidic protein; MMSE=Mini Mental State Examination; NFL=neurofilament light chain; NTB=neuropsychological test battery; PiB-PET=Pittsburgh compound B positron emission tomography; p-tau=phosphorylated tau.

Biomarker correlations at baseline are shown in Supplementary material (Table 1). P-tau181, p-tau217 (both assays), p-tau231, NFL, and GFAP all correlated with each other. Strongest correlations were observed among the p-tau markers measured with NULISA. NFL and GFAP were most strongly correlated with each other.

### Blood biomarkers, participant characteristics, and Aβ-PET positivity at baseline

Areas under the curve (AUCs) for discriminating between Aβ-positive and negative individuals in the FINGER Aβ-PET subcohort (n=46) are shown in Figure 1. P-tau217 showed the highest accuracy (AUC 0.75 for NULISA, 0.72 for Lumipulse assay). AUC for Aβ42 was 0.72; the next best performing biomarker was p-tau231 (AUC 0.66). Results for models including age are shown in Supplementary material (Fig. 1). The n (%) of Aβ-PET positives among those with normal, intermediate, and pathological plasma p-tau217 (Lumipulse) was 11/36 (31%), 4/6 (67%), and 4/4 (100%), respectively.

**Figure 1.**
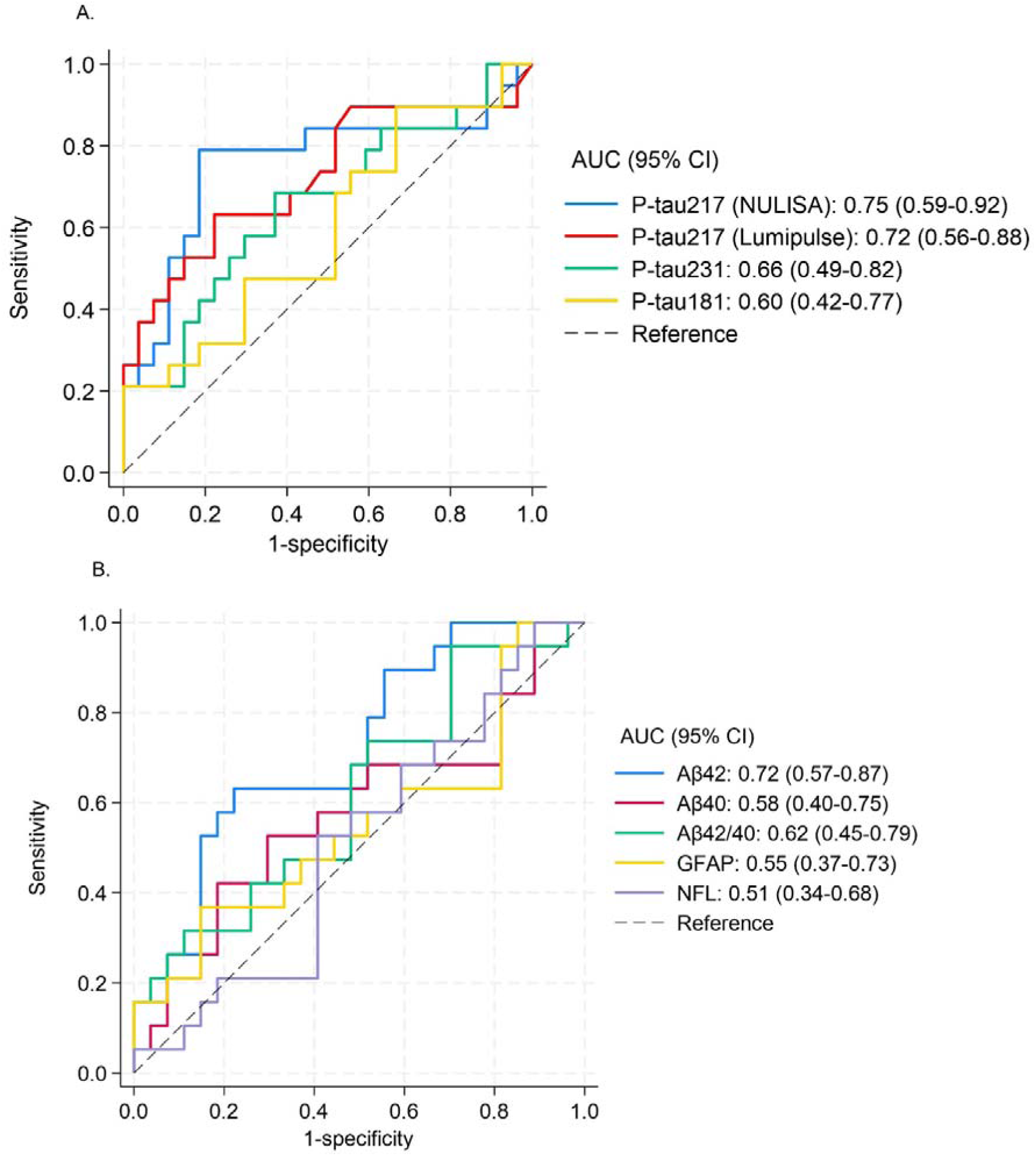
Predictive performance for Aβ-PET positivity Predictive performance of p-tau markers is shown in panel A, and Aβ, GFAP, and NFL in panel B. Biomarkers were examined as continuous variables. Unadjusted logistic regression; discrimination quantified by AUCs (95% CI). PiB-PET scan and visual read of Aβ positivity were available for 46 participants at baseline. P-tau217 was measured with both Lumipulse assay and NULISAseq™ CNS Disease Panel 120, all other markers were measured with NULISAseq™ CNS Disease Panel 120. Aβ=beta-amyloid; AUC=area under the curve; GFAP=glial fibrillary acidic protein; NFL=neurofilament light chain; PiB-PET=Pittsburgh compound B positron emission tomography; p-tau=phosphorylated tau.

Associations with participant characteristics with known impact on blood biomarker levels are shown in Supplementary material (Tables 2, 3). After adjusting for the other characteristics, higher age was associated with higher p-tau217, p-tau181, p-tau231, NFL, and GFAP, and women had higher GFAP. *APOE* ε4 carriers had lower Aβ42 and Aβ42/40, and higher p-tau217, p-tau181, p-tau231, and GFAP but not NFL. Body mass index (BMI) and estimated glomerular filtration rate (eGFR) as an indicator of renal function were negatively associated with p-tau217, NFL, and GFAP. eGFR was additionally associated with p-tau181, p-tau231, Aβ42, and Aβ40. The directions of the associations align with previous reports (38).

**Table 2.** Change in biomarker levels in *APOE4* carriers and non-carriers.

|  | Estimated mean change from baseline,<br>*APOE4* carriers |  |  | Estimated mean change from baseline,<br>*APOE4* non-carriers |  |  | Difference between *APOE4*<br>carriers and non-carriers |  |  |
| --- | --- | --- | --- | --- | --- | --- | --- | --- | --- |
|  | Estimate (95% CI) | SE | p | Estimate (95% CI) | SE | p | Estimate (95% CI) | SE | p |
| **p-tau217**<br>**(Lumipulse)** | 0.018 (0.012 to 0.024) | 0.003 | **<0.001** | 0.009 (0.006 to 0.011) | 0.001 | **<0.001** | 0.009 (0.003 to 0.016) | 0.003 | **0.006** |
| **p-tau181** | 0.133 (0.074 to 0.191) | 0.030 | **<0.001** | 0.055 (0.013 to 0.097) | 0.021 | **0.01** | 0.078 (0.007 to 0.149) | 0.036 | **0.03** |
| **p-tau217**<br>**(NULISA)** | 0.155 (0.107 to 0.202) | 0.024 | **<0.001** | 0.036 (0.004 to 0.067) | 0.016 | **0.03** | 0.119 (0.063 to 0.175) | 0.029 | **<0.001** |
| **p-tau231** | 0.143 (0.088 to 0.198) | 0.028 | **<0.001** | 0.049 (0.010 to 0.088) | 0.020 | **0.02** | 0.095 (0.028 to 0.161) | 0.034 | **0.005** |
| **Aβ42** | 0.076 (0.021 to 0.131) | 0.028 | **0.01** | 0.042 (0.003 to 0.082) | 0.020 | **0.03** | 0.034 (-0.033 to 0.100) | 0.034 | 0.33 |
| **Aβ40** | -0.013 (-0.067 to 0.040) | 0.027 | 0.62 | -0.023 (-0.062 to 0.015) | 0.020 | 0.23 | 0.010 (-0.055 to 0.075) | 0.033 | 0.76 |
| **Aβ42/40** | 0.090 (0.020 to 0.160) | 0.036 | **0.01** | 0.092 (0.042 to 0.142) | 0.025 | **<0.001** | -0.002 (-0.087 to 0.083) | 0.043 | 0.96 |
| **GFAP** | 0.151 (0.104 to 0.197) | 0.023 | **<0.001** | 0.129 (0.094 to 0.163) | 0.018 | **<0.001** | 0.022 (-0.035 to 0.079) | 0.029 | 0.45 |
| **NFL** | 0.170 (0.112 to 0.229) | 0.030 | **<0.001** | 0.099 (0.057 to 0.141) | 0.021 | **<0.001** | 0.071 (-0.000 to 0.143) | 0.036 | 0.05 |

Estimates represent biomarker change from baseline to two years in *APOE4* carriers (n=340) and non-carriers (n=694 Lumipulse p-tau217, n=696 other biomarkers), irrespective of randomization. Estimates were obtained from linear mixed-effects models, using marginal prediction methods to derive interpretable effect estimates. Positive estimates indicate increase in biomarker levels. Biomarkers were analyzed on a transformed scale; estimates are presented on the original scale (Lumipulse p-tau217 pg/ml, NULISA markers NPQ log2 fold change). Analyses were adjusted for baseline age, sex, education, study site, body mass index (BMI), estimated glomerular filtration rate (eGFR), 2-year change in eGFR, and group\*eGFR change interaction. Statistically significant p-values <0.05 are bolded.
Aβ =beta-amyloid; APOE=apolipoprotein E; CI= confidence interval; GFAP=glial fibrillary acidic protein; NFL=neurofilament light chain; p-tau=phosphorylated tau; SE=standard error.

**Table 3.** Change in biomarker levels in the intervention and control groups.

|  | Estimated mean change from baseline,<br>Intervention group |  |  | Estimated mean change from baseline,<br>Control group |  |  | Difference between<br>Intervention and Control groups |  |  |
| --- | --- | --- | --- | --- | --- | --- | --- | --- | --- |
|  | Estimate (95% CI) | SE | p | Estimate (95% CI) | SE | p | Estimate (95% CI) | SE | p |
| **p-tau217**<br>**(Lumipulse)** | 0.012 (0.008 to 0.015) | 0.002 | **<0.001** | 0.010 (0.006 to 0.013) | 0.002 | **<0.001** | 0.002 (-0.003 to 0.007) | 0.002 | 0.37 |
| **p-tau181** | 0.083 (0.037 to 0.129) | 0.023 | **<0.001** | 0.079 (0.033 to 0.126) | 0.024 | **0.001** | 0.003 (-0.062 to 0.069) | 0.033 | 0.92 |
| **p-tau217**<br>**(NULISA)** | 0.092 (0.056 to 0.128) | 0.018 | **<0.001** | 0.057 (0.021 to 0.093) | 0.018 | **0.002** | 0.035 (-0.016 to 0.086) | 0.026 | 0.18 |
| **p-tau231** | 0.083 (0.039 to 0.126) | 0.022 | **<0.001** | 0.075 (0.031 to 0.119) | 0.022 | **0.001** | 0.007 (-0.055 to 0.069) | 0.031 | 0.82 |
| **Aβ42** | 0.038 (-0.006 to 0.081) | 0.022 | 0.09 | 0.066 (0.022 to 0.110) | 0.022 | **0.003** | -0.028 (-0.090 to 0.034) | 0.032 | 0.37 |
| **Aβ40** | -0.022 (-0.065 to 0.022) | 0.022 | 0.33 | -0.015 (-0.058 to 0.028) | 0.022 | 0.50 | -0.007 (-0.068 to 0.055) | 0.031 | 0.83 |
| **Aβ42/40** | 0.087 (0.032 to 0.142) | 0.028 | **0.002** | 0.088 (0.032 to 0.143) | 0.028 | **0.002** | -0.001 (-0.079 to 0.077) | 0.040 | 0.98 |
| **GFAP** | 0.134 (0.096 to 0.172) | 0.019 | **<0.001** | 0.135 (0.096 to 0.173) | 0.020 | **<0.001** | -0.001 (-0.055 to 0.053) | 0.028 | 0.97 |
| **NFL** | 0.129 (0.081 to 0.176) | 0.024 | **<0.001** | 0.100 (0.053 to 0.148) | 0.024 | **<0.001** | 0.029 (-0.039 to 0.096) | 0.034 | 0.40 |

Estimates represent biomarker change from baseline to 2 years in the intervention and control groups and group differences in these changes. Intervention group n=553 (Lumipulse p-tau217) and n=554 (other biomarkers); control group n=548 (Lumipulse p-tau217) and n=550 (other biomarkers). Estimates were obtained from linear mixed-effects models, using marginal prediction methods to derive interpretable effect estimates. Positive estimates indicate increase in biomarker levels. Biomarkers were analyzed on a transformed scale; estimates are presented on the original scale (Lumipulse p-tau217 pg/ml, NULISA markers NPQ log2 fold change). Analyses were adjusted for baseline age, sex, education, study site, *APOE* genotype, body mass index (BMI), estimated glomerular filtration rate (eGFR), 2-year change in eGFR, and group\*eGFR change interaction. Statistically significant p-values <0.05 are bolded.
Aβ =beta-amyloid; CI=confidence interval; GFAP=glial fibrillary acidic protein; NFL=neurofilament light chain; p-tau=phosphorylated tau; SE=standard error.

### Blood biomarkers and cognitive performance

Figure 2 shows the associations between baseline blood biomarker levels (continuous variables) and cognitive performance (Neuropsychological Test Battery, NTB, z-score for global cognition, primary FINGER trial outcome). Results for other cognitive outcomes (NTB z-scores for memory, abbreviated memory, executive functioning, and processing speed) are shown in Supplementary material (Fig. 2–5).

**Figure 2.**
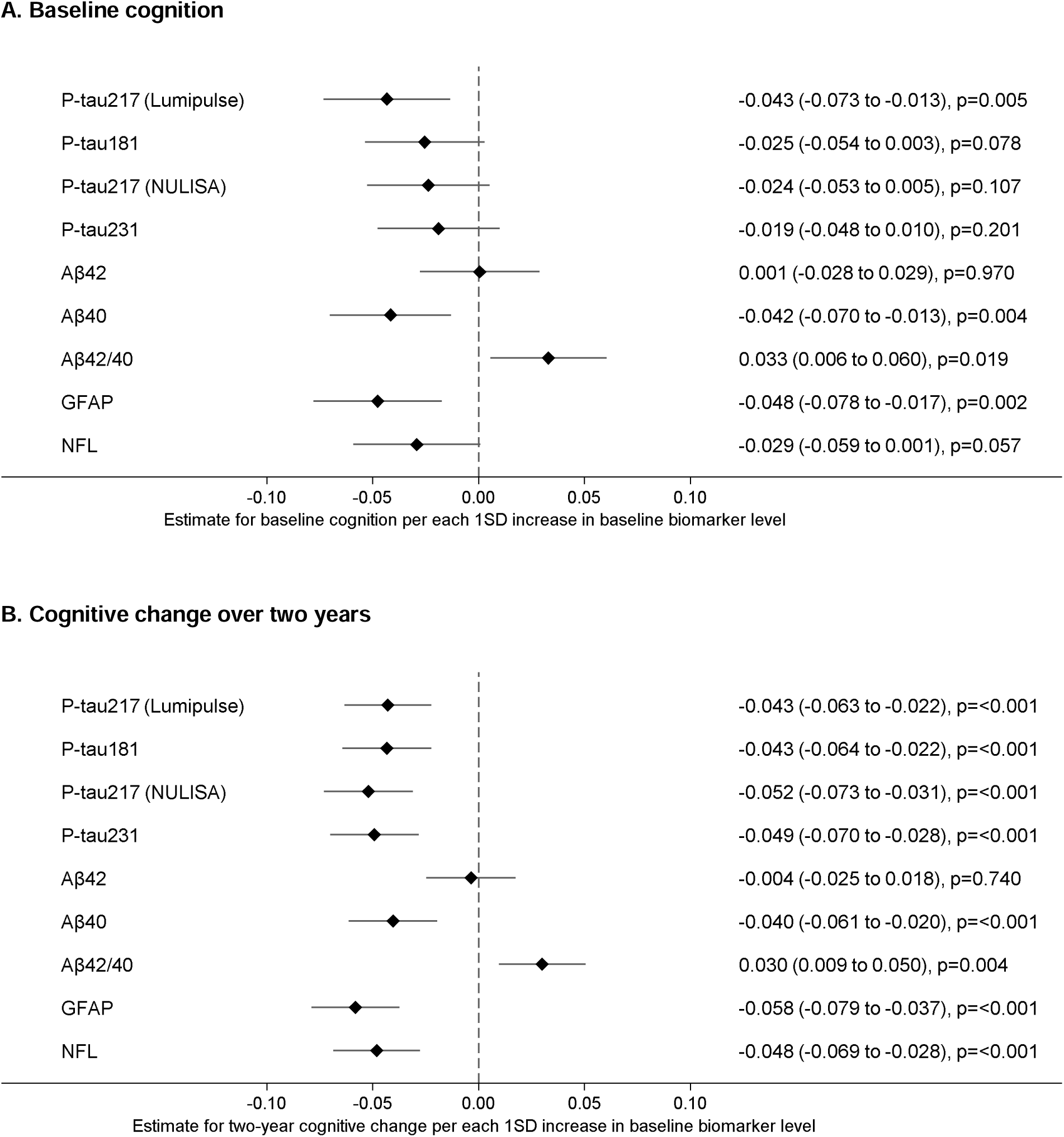
Associations between baseline biomarker levels and cognition (NTB Global cognition, primary trial outcome) Estimates (95% CI) were obtained from linear regression models (for baseline cognition, A) and mixed-effects models (for cognitive change, B), using marginal prediction methods to derive interpretable effect estimates. Results are interpreted as differences in baseline cognitive performance (A) or cognitive change in two years (B), per 1 standard deviation (SD) higher baseline biomarker level, in the whole cohort (irrespective of randomization). Negative estimates indicate worse cognitive performance or less favorable cognitive trajectory (less improvement) with increasing baseline biomarker levels. Analyses were adjusted for baseline age, sex, education, study site, *APOE* genotype, body mass index (BMI), and estimated glomerular filtration rate (eGFR) as an indicator of renal function. Baseline analysis n=1220 (Lumipulse p-tau217) and n=1223 (other biomarkers); longitudinal analysis n=1154 (Lumipulse p-tau217) and n=1157 (other biomarkers). Aβ=beta-amyloid; GFAP=glial fibrillary acidic protein; NFL=neurofilament light chain; NTB=neuropsychological test battery; p-tau=phosphorylated tau.

Cross-sectionally at baseline, higher levels of Lumipulse p-tau217, Aβ40, and GFAP, and lower Aβ42/40 were associated with poorer global cognition (Fig. 2A). Higher Lumipulse p-tau217 and GFAP were also associated with poorer memory (Suppl. Fig. 2A); only p-tau217 showed an association with an abbreviated memory score including selected tests with more complex processing and longer recall delay (Suppl. Fig. 3A). Higher Aβ40 and GFAP were associated with poorer executive functioning and processing speed (Suppl. Fig. 4A, 5A). Aβ42/40 was also associated with executive functioning, and NULISA p-tau217 with processing speed. No statistically significant associations were found for Aβ42, p-tau181, ptau231, or NFL in any cognitive outcome.

We found no indication that the observed cross-sectional associations between baseline biomarker levels and cognition were significantly different between *APOE* ε4 carriers and non-carriers, by age, or by the level of FINRISK cardiovascular risk score as an indicator of vascular risk factor burden (biomarker × *APOE,* biomarker × age, and biomarker × vascular risk factor burden interactions p>0.05). However, a signal was observed for a potentially stronger association between biomarkers and cognition in *APOE* ε4 carriers (Suppl. Fig. 6). Similarly, the associations between p-tau markers and memory appeared somewhat stronger in younger participants and participants with lower vascular risk factor burden (Suppl. Fig. 7,8). Regarding sex, there were some statistically significant interactions with Aβ. Higher Aβ40 associated with worse global cognition and memory in women but not in men (Suppl. Fig. 9). Similarly, Aβ42 showed a negative association with cognition in women but not in men. In general, despite statistically non-significant interactions, there was a signal suggesting that higher biomarker levels potentially associated with worse cognition, particularly memory and processing speed, more strongly in women (Suppl. Fig. 9).

Longitudinally, higher baseline biomarker levels were generally associated with less favorable cognitive trajectories (less cognitive improvement over two years), irrespective of randomization group. In global cognition and memory, this was observed for all biomarkers, apart from Aβ42 (Fig. 2B and Suppl. Fig. 2B). NULISA p-tau217, p-tau231, and GFAP showed statistically significant associations also with abbreviated memory (Suppl. Fig. 3B). All biomarkers apart from the Aβ markers were associated with change in executive functioning (Suppl. Fig. 4B). P-tau217, p-tau231, Aβ40, GFAP, and NFL were associated with change in processing speed (Suppl. Fig. 5B). Aβ42 was the only biomarker not associated with change in any cognitive outcome.

Results of biomarker associations with risk of decline in global cognition, memory, executive functioning, and processing speed from baseline to two years are shown in Supplementary material (Table 4, Fig. 10). Having pathological p-tau217 (Lumipulse, >0.34 pg/ml) was associated with approximately twofold higher odds of decline in the memory domain (reference group normal <0.22 pg/ml, Suppl. Table 4). In analyses with continuous biomarker levels, both p-tau217 markers predicted decline in executive functioning (Suppl. Fig. 10). P-tau217 Lumipulse, Aβ42, Aβ42/40, and GFAP showed statistically significant, or borderline significant, associations with risk of decline in processing speed.

### Blood biomarkers and intervention effects on cognition

Next, we studied if baseline blood biomarker levels influenced the intervention effects on cognition. Results for global cognition are shown in Figure 3 and Supplementary material (Table 5); results for the other cognitive outcomes are shown in Supplementary material (Tables 6-9). We found that the intervention effects did not differ between individuals with lower vs. higher biomarker levels in any cognitive outcome (all randomization group × time × biomarker interactions p>0.05; Fig. 3, Suppl. Tables 5-9). Stratified analysis showed that the estimates for the intervention-control difference in cognitive change were consistently positive, i.e., in favor of the intervention group, both in participants with lower and higher biomarker levels (Fig. 3, Suppl. Tables 5-9). As for the p-tau markers and outcomes global cognition, executive functioning, and processing speed, we observed some statistically significant intervention-control differences, in favor of the intervention group, in the lower (less pathological) but not the higher (more pathological) biomarker-level group (Fig. 3, Suppl. Tables 5, 8, 9). Same was observed for GFAP in executive functioning and processing speed (Suppl. Tables 8, 9). For Aβ and global cognition, a statistically significantly positive estimate was found specifically in the higher Aβ40 group, and consequently, in the lower Aβ42/40 group (Fig. 3, Suppl. Table 5). Results for *APOE* ε4 carriers vs. non-carriers are presented in Supplementary material (Fig. 11); all *APOE* × randomization group × time × biomarker interactions were statistically non-significant.

**Figure 3.**
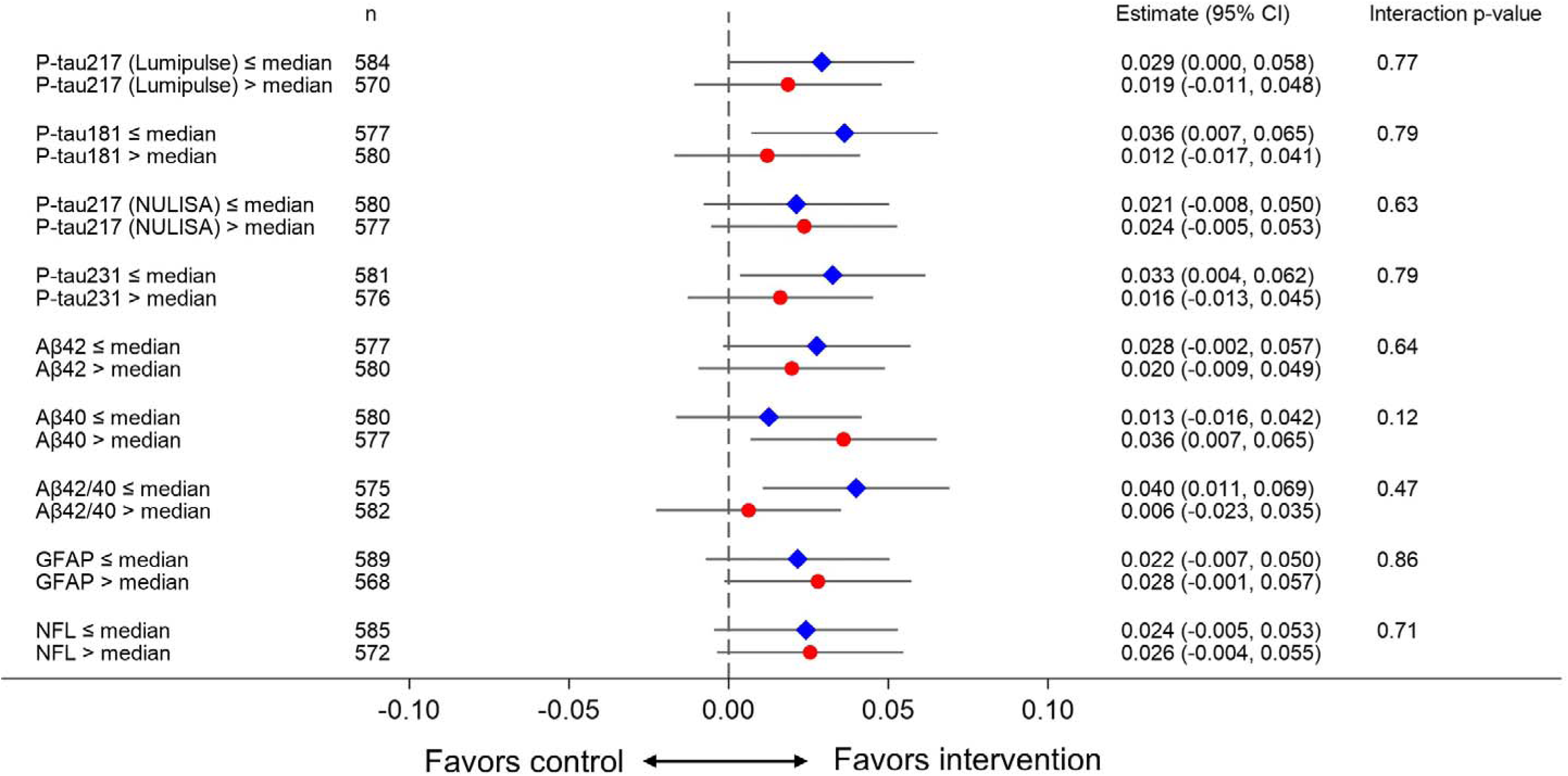
Intervention effect modification by baseline biomarker levels (outcome NTB Global cognition, primary trial outcome) Estimates (95% CI) were obtained from linear mixed-effects models, using marginal prediction methods to derive interpretable effect estimates. Estimates are interpreted as difference between the intervention and control groups in cognitive change per year; positive estimates indicate more favorable cognitive change in the intervention group. Biomarker × group × time interactions were examined to assess whether baseline biomarker levels modified the intervention effects on cognition, i.e., whether the differences in cognitive change between intervention and control groups differed by baseline biomarker levels. Interactions with p>0.05 indicate that the intervention effect did not significantly differ by baseline biomarker levels. Interactions were derived from models where biomarkers were entered as continuous variables, but for visualization, estimates for intervention effect are presented within biomarker strata (below median = blue diamond, above median = red circle). Analyses were adjusted for age, sex, education, study site, *APOE* genotype, body mass index (BMI), and estimated glomerular filtration rate (eGFR). Aβ=beta-amyloid; GFAP=glial fibrillary acidic protein; NFL=neurofilament light chain; NTB=neuropsychological test battery; p-tau=phosphorylated tau.

### Changes in blood biomarkers

Changes in blood biomarker levels from baseline to two years in *APOE* ε4 carriers vs. non-carriers and in the intervention vs. control groups, are shown in Tables 2 and 3. Biomarker levels increased in both *APOE* ε4 carriers and non-carriers, but ε4 carriers showed greater increase than non-carriers in p-tau markers (Table 2). Changes in Aβ and GFAP were similar. Overall, biomarker changes were subtle in this cohort, as after two years, most participants were still below the pathological threshold for Lumipulse p-tau217 (normal n=907, 82.1%, intermediate n=103, 9.3%, and pathological n=95, 8.6%). Estimated changes in p-tau217 status from baseline to two years are shown in Supplementary material (Suppl. Table 10).

The biomarker trajectories did not differ between the intervention and control groups (Table 3), and the results remained unchanged in a sensitivity analysis accounting additionally for BMI change (Suppl. Table 11). Also, biomarker changes were similar irrespective of adherence to intervention (rate of participation in scheduled activities, Suppl. Table 12). There were no significant differences between *APOE* ε4 carriers and non-carriers in the intervention effects on biomarkers (*APOE* × randomization group × time interactions p>0.05, Suppl. Table 13).

## DISCUSSION

To our knowledge, FINGER is the first large-scale lifestyle dementia risk reduction RCT where core AD-related blood biomarkers have been investigated in at-risk older adults without substantial cognitive impairment. We observed that the majority of study participants, 85%, had normal Lumipulse p-tau217, indicating a largely preclinical population with relatively low AD biomarker burden. Higher, more pathological levels of blood biomarkers were overall associated with worse baseline cognition and less favorable cognitive change over two years. The beneficial effects of the FINGER intervention on cognition did not significantly differ between individuals with lower and higher blood biomarker levels. Over two years, biomarker levels increased, though the rate of change was subtle and only a small proportion crossed the pathology threshold for Lumipulse p-tau217. Biomarker trajectories were similar in the intervention and control groups.

The observed associations between blood biomarkers and cognition support their emerging role in identifying cognitively unimpaired older individuals with an increased risk of decline. This aligns with previous recent evidence (39), although not all findings have been consistent (40). In our cohort, more pathological baseline biomarker levels were consistently associated with less favorable cognitive trajectories over two years, and having pathological p-tau217 at baseline approximately doubled the odds of subsequent decline in memory performance. Despite these associations, blood biomarkers, including p-tau217, showed only modest performance for identifying Aβ-PET positivity, with lower discrimination than typically reported in clinical cohorts (7). This likely reflects the preclinical stage as previously demonstrated (41) and/or the contribution of vascular and other non-AD pathologies in the FINGER at-risk population. Thus, while blood biomarkers may be informative for risk stratification, they remain insufficiently accurate as stand-alone screening tools for detecting Aβ pathology in cognitively unimpaired populations.

The longitudinal biomarker analyses indicated that the FINGER intervention did not significantly influence trajectories of core AD-related blood biomarkers over two years. Changes in all studied biomarkers were similar in the multidomain lifestyle intervention and control (regular health advice) groups, and accounting for adherence (rate of participation in scheduled activities) did not affect the results. The cognitive benefits of multidomain lifestyle interventions are thus unlikely to be explained by measurable changes in core AD-related pathology over this time frame, and may instead be driven by cardiometabolic, inflammatory, or cognitive-reserve pathways (42). Consistent with this interpretation, the two-year US POINTER multidomain lifestyle intervention with demonstrated cognitive efficacy did not affect the accumulation of Aβ and tau pathology on PET (43). Also, no intervention effect on blood p-tau181 was reported in the three-year French MAPT multidomain lifestyle RCT (44). However, the suitability and sensitivity of AD blood biomarkers to monitor intervention effects will require further clarification. In the evoke and evoke+ RCTs investigating the efficacy of semaglutide in early AD, the treatment-related changes observed in CSF AD biomarkers were not fully reflected in blood-based measures, and the reasons for this discrepancy remain unclear (45).

An important observation in our study was that most FINGER participants had normal blood biomarker levels at baseline (85% with blood p-tau217 below the pathological threshold). A two-year intervention time frame may thus be too limited for significant detectable biomarker changes to occur in this population. Overall, irrespective of randomization, we found increases in p-tau markers, GFAP, and NFL, consistent with age-related accumulation of pathology (46), patterns observed in placebo groups of RCTs investigating anti-amyloid therapies (24,25), and the few available longitudinal studies in general population-based cohorts (47,48). Interpretation of Aβ remains more challenging given the known limitations of this blood marker, e.g., variability and sensitivity to pre-analytical sample handling and peripheral factors (9). Despite the observed increases, only few FINGER participants crossed the pathology threshold for Lumipulse p-tau217 during the trial, suggesting only subtle progression of pathology in this early at-risk population. It remains therefore unclear to what extent multidomain lifestyle interventions could lead to biomarker changes in individuals with pathological biomarker profiles.

A key open question in multidomain lifestyle interventions is whether individuals with underlying AD pathology can benefit from them. In our study, baseline blood biomarker levels did not significantly modify the cognitive response to the intervention. This aligns mostly with findings from the French MAPT (44) and Japanese J-MINT RCTs (49), although evidence remains limited (44,49–51). Similarly, the US-POINTER RCT found no evidence that the intervention effects on cognition differed by baseline Aβ or tau burden on PET (43). In our stratified analyses, we observed potentially greater FINGER intervention benefits among individuals with less pathological p-tau and GFAP levels, although these patterns were not fully consistent, and the results of exploratory subgroup analyses should be interpreted with caution. These results could suggest that older adults with more advanced neuropathological changes may require additional interventions, such as pharmacological treatments, to achieve optimal cognitive benefits. Our main findings nevertheless indicate that multidomain lifestyle interventions like FINGER support cognitive functioning in at-risk older adults even in the early stages of AD neuropathology.

This study has several major strengths. The cohort is a large, well-characterized at-risk cohort that is broadly representative of the Finnish general population of the same age. Cognitive assessment was comprehensive and was conducted using a sensitive neuropsychological test battery which is suitable for capturing subtle cognitive changes during early at-risk stages. Blood biomarkers were assessed at two timepoints using validated state-of-the-art methods, covering a broad panel of core AD-related biomarkers and including Lumipulse p-tau217 which is already used in clinical practice and for which established cut-offs are available. Importantly, we were able to investigate both intervention response and longitudinal biomarker trajectories in a large dementia risk reduction RCT with demonstrated cognitive efficacy. Furthermore, the inclusion of an Aβ PET imaging subsample, although relatively small, allowed comparison of blood biomarkers with the gold-standard measure of Aβ pathology. Some limitations should also be considered. First, the use of the novel NULISA platform and its reliance on relative biomarker quantification may limit direct comparability of the biomarker findings with other studies. Second, the two-year time frame may not capture substantial changes in AD neuropathology or the longer-term effects of healthy lifestyle changes in this early at-risk population. Longer-term follow-up and biomarker analyses will be important to further elucidate these relationships.

To conclude, the two-year FINGER multidomain lifestyle intervention benefitted cognition in at-risk older adults without substantial impairment irrespective of the baseline levels of core AD-related blood biomarkers. These cognitive benefits may primarily involve mechanistic pathways other than those captured by the core AD blood biomarkers. Overall, changes in core AD blood biomarkers over two years were limited in this relatively healthy, early at-risk cohort from the general population. While biomarker levels were associated with cognition, their ability to detect underlying Aβ pathology was more limited than in clinical patient cohorts. Together, these findings support the cautious use of AD blood biomarkers for risk stratification in preclinical and/or at-risk populations. Future studies integrating blood biomarkers of vascular, metabolic, inflammatory, and synaptic pathways will be essential to further clarify the mechanisms of multidomain lifestyle interventions. It will also be important to replicate and extend these findings worldwide across ethnically, culturally, and socioeconomically diverse populations and settings. This is needed to inform the design of future dementia prevention strategies.

## Data Availability

FINGER dataset can be requested from University of Eastern Finland. De-identified data can be shared with academic researchers who provide a research plan accepted by the study group (estimated time frame one month). Data transfer agreements must be completed before accessing the data. Enquiries should be directed to.

## ETHICAL APPROVAL

FINGER received ethical approval from the Coordinating Ethics Committee of Helsinki and Uusimaa Hospital District in Finland (94/13/03/00/09), and all participants provided written informed consent.

