## Supplementary material for "Blood Biomarkers of Alzheimer’s Disease Neuropathology, Cognition, and Response to a Multidomain Lifestyle Intervention: Secondary analysis of the FINGER Randomized Controlled Dementia Risk Reduction Trial"

**Table 1**. Correlation matrix for the blood biomarkers (baseline levels)

| | **p-tau217 (Lumipulse)** | **p-tau181** | **p-tau217 (NULISA)** | **p-tau231** | **NFL** | **GFAP** | **Aβ42** | **Aβ40** | **Aβ42/40** |
|---|---|---|---|---|---|---|---|---|---|
| **p-tau217 (Lumipulse)** | 1.00 | | | | | | | | |
| **p-tau181** | 0.56** | 1.00 | | | | | | | |
| **p-tau217 (NULISA)** | 0.61** | 0.68** | 1.00 | | | | | | |
| **p-tau231** | 0.66** | 0.93** | 0.73** | 1.00 | | | | | |
| **NFL** | 0.35** | 0.36** | 0.26** | 0.39** | 1.00 | | | | |
| **GFAP** | 0.40** | 0.29** | 0.33** | 0.37** | 0.52** | 1.00 | | | |
| **Aβ42** | -0.01 | 0.17** | 0.09* | 0.14** | 0.18** | 0.03 | 1.00 | | |
| **Aβ40** | 0.17** | 0.18** | 0.10** | 0.20** | 0.38** | 0.29** | 0.37** | 1.00 | |
| **Aβ42/40** | -0.16** | -0.02 | -0.06* | -0.07* | -0.15** | -0.23** | 0.52** | -0.52** | 1.00 |

Table shows the Spearman rank correlation coefficients (ρ) and associated p-values, *$p<0.05$, ** $p<0.001$. Lumipulse p-tau217 n=1221, other markers (NULISAseq™ CNS Disease Panel 120) n=1224.

Aβ=beta-amyloid; GFAP=glial fibrillary acidic protein; NFL=neurofilament light chain; p-tau=phosphorylated tau.

**Table 2.** Baseline associations of participant characteristics and biomarker levels (p-tau217, p-tau181, p-tau231)

| | p-tau217 (Lumipulse) | | p-tau181 | | p-tau217 (NULISA) | | p-tau231 | |
|---|---|---|---|---|---|---|---|---|
| | Estimate (95% CI) | p | Estimate (95% CI) | p | Estimate (95% CI) | p | Estimate (95% CI) | p |
| ***UNADJUSTED*** | | | | | | | | |
| Age, years | 0.003 (0.002 to 0.003) | **<0.001** | 0.032 (0.025 to 0.039) | **<0.001** | 0.022 (0.016 to 0.028) | **<0.001** | 0.035 (0.027 to 0.042) | **<0.001** |
| Women (vs. men) | 0.006 (0.000 to 0.012) | **0.04** | -0.017 (-0.087 to 0.053) | 0.63 | -0.009 (-0.067 to 0.050) | 0.77 | 0.011 (-0.061 to 0.083) | 0.77 |
| Education, years | -0.001 (-0.001 to 0.000) | 0.23 | -0.008 (-0.018 to 0.003) | 0.15 | -0.005 (-0.014 to 0.003) | 0.20 | -0.010 (-0.020 to 0.001) | 0.06 |
| *APOE ε4* carriers (vs. non-carriers) | 0.030 (0.022 to 0.037) | **<0.001** | 0.148 (0.071 to 0.225) | **<0.001** | 0.250 (0.187 to 0.314) | **<0.001** | 0.247 (0.169 to 0.325) | **<0.001** |
| BMI, kg/m$^2$ | -0.001 (-0.002 to -0.000) | **0.002** | -0.003 (-0.011 to 0.004) | 0.37 | -0.010 (-0.017 to -0.004) | **0.002** | -0.007 (-0.014 to 0.001) | 0.10 |
| eGFR, ml/min/1.73m$^2$ | -0.001 (-0.001 to -0.001) | **<0.001** | -0.009 (-0.011 to -0.006) | **<0.001** | -0.008 (-0.010 to -0.006) | **<0.001** | -0.011 (-0.013 to -0.008) | **<0.001** |
| ***ADJUSTED*** | | | | | | | | |
| Age, years | 0.002 (0.001 to 0.003) | **<0.001** | 0.027 (0.019 to 0.035) | **<0.001** | 0.017 (0.011 to 0.024) | **<0.001** | 0.029 (0.021 to 0.036) | **<0.001** |
| Women (vs. men) | 0.002 (-0.003 to 0.008) | 0.44 | -0.055 (-0.124 to 0.013) | 0.12 | -0.039 (-0.095 to 0.017) | 0.17 | -0.033 (-0.102 to 0.035) | 0.34 |
| Education, years | -0.000 (-0.001 to 0.001) | 0.59 | -0.001 (-0.011 to 0.009) | 0.86 | -0.002 (-0.011 to 0.006) | 0.57 | -0.003 (-0.013 to 0.007) | 0.58 |
| *APOE ε4* carriers (vs. non-carriers) | 0.032 (0.024 to 0.039) | **<0.001** | 0.166 (0.092 to 0.242) | **<0.001** | 0.266 (0.204 to 0.328) | **<0.001** | 0.267 (0.192 to 0.341) | **<0.001** |
| BMI, kg/m$^2$ | -0.001 (-0.002 to -0.000) | **0.001** | -0.002 (-0.009 to 0.006) | 0.65 | -0.009 (-0.015 to -0.003) | **0.01** | -0.005 (-0.012 to 0.003) | 0.20 |
| eGFR, ml/min/1.73m$^2$ | -0.001 (-0.001 to -0.001) | **<0.001** | -0.006 (-0.009 to -0.003) | **<0.001** | -0.006 (-0.009 to -0.004) | **<0.001** | -0.008 (-0.010 to -0.005) | **<0.001** |

Estimates represent differences in baseline biomarker levels comparing mean vs mean + 1 unit of the predictor (or differences between categories), while holding other covariates constant at mean (adjusted models; all other characteristics included as covariates). Estimates were obtained from linear regression models, using marginal prediction methods to derive interpretable effect estimates. Biomarkers were analyzed on a transformed scale; estimates are presented on the original scale (Lumipulse p-tau217 unit pg/ml, NULISAseq™ CNS Disease Panel 120 unit NPQ, NULISA Protein Quantification Unit, log2-scale values reflecting relative protein abundances). eGFR: indicator of renal function, with higher values indicating better function.

*APOE*=apolipoprotein E; BMI=body mass index; eGFR=estimated glomerular filtration rate; p-tau=phosphorylated tau.

**Table 3.** Baseline associations of participant characteristics and biomarker levels (Aβ, GFAP, NFL)

| | **Aβ42** | | **Aβ40** | | **Aβ42/40** | | **GFAP** | | **NFL** | |
|---|---|---|---|---|---|---|---|---|---|---|
| | Estimate (95% CI) | p | Estimate (95% CI) | p | Estimate (95% CI) | p | Estimate (95% CI) | p | Estimate (95% CI) | p |
| ***UNADJUSTED*** | | | | | | | | | | |
| Age, years | 0.012 (0.004 to 0.020) | **0.002** | 0.018 (0.011 to 0.024) | **<0.001** | -0.004 (-0.012 to 0.005) | 0.41 | 0.049 (0.041 to 0.057) | **<0.001** | 0.046 (0.038 to 0.054) | **<0.001** |
| Women (vs. men) | 0.057 (-0.014 to 0.128) | 0.11 | 0.086 (0.022 to 0.149) | **0.01** | -0.042 (-0.121 to 0.038) | 0.30 | 0.229 (0.151 to 0.306) | **<0.001** | 0.009 (-0.071 to 0.089) | 0.83 |
| Education, years | -0.008 (-0.018 to 0.002) | 0.13 | -0.012 (-0.022 to -0.003) | **0.01** | 0.005 (-0.006 to 0.016) | 0.39 | -0.021 (-0.032 to -0.009) | **0.001** | -0.015 (-0.027 to -0.004) | **0.01** |
| *APOE ε4* carriers (vs. non-carriers) | -0.186 (-0.264 to -0.108) | **<0.001** | 0.012 (-0.058 to 0.081) | 0.75 | -0.218 (-0.304 to -0.133) | **<0.001** | 0.180 (0.097 to 0.264) | **<0.001** | -0.027 (-0.112 to 0.057) | 0.52 |
| BMI, kg/m$^2$ | -0.001 (-0.008 to 0.007) | 0.87 | 0.005 (-0.002 to 0.011) | 0.20 | -0.004 (-0.013 to 0.005) | 0.37 | -0.021 (-0.030 to -0.013) | **<0.001** | -0.025 (-0.033 to -0.016) | **<0.001** |
| eGFR, ml/min/1.73m$^2$ | -0.011 (-0.014 to -0.009) | **<0.001** | -0.014 (-0.016 to -0.011) | **<0.001** | 0.002 (-0.001 to 0.005) | 0.25 | -0.016 (-0.018 to -0.013) | **<0.001** | -0.018 (-0.021 to -0.015) | **<0.001** |
| ***ADJUSTED*** | | | | | | | | | | |
| Age, years | 0.000 (-0.008 to 0.008) | 0.97 | 0.005 (-0.002 to 0.012) | 0.18 | -0.003 (-0.012 to 0.006) | 0.46 | 0.036 (0.028 to 0.044) | **<0.001** | 0.031 (0.023 to 0.039) | **<0.001** |
| Women (vs. men) | 0.025 (-0.045 to 0.095) | 0.48 | 0.035 (-0.026 to 0.096) | 0.26 | -0.029 (-0.108 to 0.050) | 0.47 | 0.188 (0.116 to 0.259) | **<0.001** | -0.034 (-0.108 to 0.039) | 0.36 |
| Education, years | -0.004 (-0.015 to 0.006) | 0.42 | -0.009 (-0.018 to 0.000) | 0.05 | 0.005 (-0.006 to 0.017) | 0.37 | -0.010 (-0.021 to 0.001) | 0.07 | -0.007 (-0.018 to 0.004) | 0.20 |
| *APOE ε4* carriers (vs. non-carriers) | -0.180 (-0.256 to -0.103) | **<0.001** | 0.014 (-0.052 to 0.080) | 0.68 | -0.214 (-0.300 to -0.128) | **<0.001** | 0.194 (0.119 to 0.270) | **<0.001** | -0.011 (-0.087 to 0.065) | 0.77 |
| BMI, kg/m$^2$ | -0.003 (-0.011 to 0.004) | 0.41 | 0.002 (-0.004 to 0.009) | 0.50 | -0.004 (-0.012 to 0.005) | 0.38 | -0.002 (-0.030 to -0.015) | **<0.001** | -0.025 (-0.033 to -0.017) | **<0.001** |
| eGFR, ml/min/1.73m$^2$ | -0.011 (-0.014 to -0.008) | **<0.001** | -0.013 (-0.015 to -0.010) | **<0.001** | 0.001 (-0.002 to 0.004) | 0.47 | -0.011 (-0.014 to -0.008) | **<0.001** | -0.015 (-0.018 to -0.012) | **<0.001** |

Estimates represent differences in baseline biomarker levels comparing mean vs mean + 1 unit of the predictor (or differences between categories), while holding other covariates constant at mean (adjusted models; all other characteristics included as covariates). Estimates were obtained from linear regression models, using marginal prediction methods to derive interpretable effect estimates. Biomarkers were analyzed on a transformed scale; estimates are presented on the original scale (Lumipulse p-tau217 unit pg/ml, NULISAseq™ CNS Disease Panel 120 unit NPQ, NULISA Protein Quantification Unit, log2-scale values reflecting relative protein abundances). eGFR: indicator of renal function, with higher values indicating better function.

Aβ=beta-amyloid; *APOE*=apolipoprotein E; BMI=body mass index; eGFR=estimated glomerular filtration rate; GFAP=glial fibrillary acidic protein; NFL=neurofilament light chain.

**Table 4.** Risk of cognitive decline according to baseline p-tau217 status (Lumipulse assay)

| | Odds ratio (95% CI), p-value | | |
|---|---|---|---|
| | Normal p-tau217 (n=936) | Intermediate p-tau217 (n=82) | Pathological p-tau217 (n=70) |
| Global cognition | 1 (reference) | 1.14 (0.67 to 1.92), p=0.64 | 1.69 (0.96 to 2.96), p=0.07 |
| Memory | 1 (reference) | 1.45 (0.86 to 2.44), p=0.16 | **2.02 (1.15 to 3.54), p=0.01** |
| Abbreviated memory | 1 (reference) | 1.02 (0.60 to 1.71), p=0.95 | 1.59 (0.93 to 2.73), p=0.09 |
| Executive functioning | 1 (reference) | 1.32 (0.81 to 2.17), p=0.26 | 1.39 (0.81 to 2.39), p=0.23 |
| Processing speed | 1 (reference) | 1.13 (0.69 to 1.84), p=0.62 | 1.40 (0.83 to 2.36), p=0.21 |

Risk of cognitive decline was defined as a negative change in the Neuropsychological Test Battery (NTB) z-score from baseline to two years. Decline in each domain was analyzed separately (global cognition, memory, abbreviated memory, executive functioning, processing speed). Cut-offs for Lumipulse p-tau217 status: normal <0.22 pg/ml, intermediate 0.22–0.34 pg/ml, pathological >0.34 pg/ml. Logistic regression with odds ratios (ORs) and 95% confidence intervals (Cis) are shown relative to the normal group. Analyses were adjusted for the baseline cognitive score (z-score for global cognition, memory, abbreviated memory, executive functioning, or processing speed), baseline age, sex, education, study site, *APOE* genotype, body mass index (BMI), and estimated glomerular filtration rate (eGFR).

**Table 5.** Baseline biomarker levels and intervention effects on cognition (global cognition)

| | Baseline level of biomarker | Mean change (SE) in NTB Global Cognition | | Difference in mean change between Intervention and Control, per year | | Difference in intervention effect by biomarker level (randomization*BM*time), stratified analysis | | Difference in intervention effect by biomarker level (randomization*BM*time), main analysis | |
|---|---|---|---|---|---|---|---|---|---|
| | | Intervention | Control | Estimate (95% CI) | p | Estimate (95% CI) | p | Estimate (95% CI) | p |
| p-tau217 (Lumipulse) | Normal | 0.110 (0.008) | 0.087 (0.008) | 0.023 (0.001 to 0.045) | **0.04** | 0.006 (-0.053 to 0.064) | 0.85 | -0.003 (-0.024 to 0.017) | 0.77 |
| | Intermediate or pathological | 0.056 (0.019) | 0.028 (0.020) | 0.029 (-0.026 to 0.083) | 0.30 | | | | |
| p-tau217 (Lumipulse) | ≤ median | 0.123 (0.011) | 0.094 (0.010) | 0.029 (0.000 to 0.058) | 0.05 | -0.011 (-0.052 to 0.031) | 0.62 | -0.003 (-0.024 to 0.017) | 0.77 |
| | > median | 0.080 (0.010) | 0.062 (0.011) | 0.019 (-0.011 to 0.048) | 0.22 | | | | |
| p-tau181 | ≤ median | 0.120 (0.011) | 0.084 (0.010) | 0.036 (0.007 to 0.065) | **0.02** | -0.024 (-0.065 to 0.017) | 0.25 | -0.003 (-0.024 to 0.018) | 0.79 |
| | > median | 0.083 (0.010) | 0.071 (0.011) | 0.012 (-0.017 to 0.041) | 0.42 | | | | |
| p-tau217 (NULISA) | ≤ median | 0.119 (0.010) | 0.098 (0.011) | 0.021 (-0.008 to 0.050) | 0.15 | 0.002 (-0.039 to 0.044) | 0.91 | 0.005 (-0.016 to 0.026) | 0.63 |
| | > median | 0.083 (0.011) | 0.059 (0.010) | 0.024 (-0.005 to 0.053) | 0.11 | | | | |
| p-tau231 | ≤ median | 0.126 (0.011) | 0.093 (0.010) | 0.033 (0.004 to 0.062) | **0.03** | -0.016 (-0.057 to 0.025) | 0.43 | -0.003 (-0.024 to 0.018) | 0.79 |
| | > median | 0.078 (0.010) | 0.062 (0.011) | 0.016 (-0.013 to 0.045) | 0.28 | | | | |
| Aβ42 | ≤ median | 0.104 (0.011) | 0.076 (0.010) | 0.028 (-0.002 to 0.057) | 0.07 | -0.008 (-0.049 to 0.034) | 0.71 | 0.005 (-0.016 to 0.026) | 0.64 |
| | > median | 0.100 (0.010) | 0.080 (0.011) | 0.020 (-0.009 to 0.049) | 0.19 | | | | |
| Aβ40 | ≤ median | 0.111 (0.011) | 0.098 (0.010) | 0.013 (-0.016 to 0.042) | 0.40 | 0.023 (-0.018 to 0.065) | 0.27 | 0.017 (-0.004 to 0.038) | 0.12 |
| | > median | 0.092 (0.010) | 0.056 (0.011) | 0.036 (0.007 to 0.065) | **0.02** | | | | |
| Aβ42/40 | ≤ median | 0.093 (0.011) | 0.053 (0.010) | 0.040 (0.011 to 0.069) | **0.01** | -0.034 (-0.075 to 0.007) | 0.11 | -0.008 (-0.028 to 0.013) | 0.47 |
| | > median | 0.110 (0.010) | 0.103 (0.011) | 0.006 (-0.023 to 0.035) | 0.67 | | | | |
| GFAP | ≤ median | 0.122 (0.011) | 0.100 (0.010) | 0.022 (-0.007 to 0.050) | 0.14 | 0.006 (-0.035 to 0.047) | 0.76 | 0.002 (-0.019 to 0.023) | 0.86 |
| | > median | 0.082 (0.010) | 0.054 (0.011) | 0.028 (-0.001 to 0.057) | 0.06 | | | | |
| NFL | ≤ median | 0.126 (0.010) | 0.102 (0.010) | 0.024 (-0.005 to 0.053) | 0.10 | 0.001 (-0.040 to 0.042) | 0.95 | 0.004 (-0.017 to 0.024) | 0.71 |
| | > median | 0.077 (0.010) | 0.052 (0.011) | 0.026 (-0.004 to 0.055) | 0.09 | | | | |

Estimates were obtained from linear mixed-effects models, using marginal prediction methods to derive interpretable effect estimates. Analyses were adjusted for baseline age, sex, education, study site, *APOE* genotype, body mass index (BMI), and estimated glomerular filtration rate (eGFR). Biomarker (BM) × randomization group × time interactions derived from the model where biomarkers were entered as continuous variables are considered the main results; estimates for intervention effects are additionally presented within biomarker strata (median split and predefined Lumipulse p-tau217 categories, normal <0.22 pg/ml vs. intermediate 0.22–0.34 pg/ml or pathological >0.34 pg/ml). Biomarker × randomization group × time interactions p>0.05 indicate that the intervention effects on cognition did not significantly differ by baseline biomarker levels.

Aβ=beta-amyloid; BM=biomarker; GFAP=glial fibrillary acidic protein; NFL=neurofilament light chain; NTB=neuropsychological test battery; p-tau=phosphorylated tau.

**Table 6.** Baseline biomarker levels and intervention effects on cognition (memory)

| | Baseline level of biomarker | Mean change (SE) in NTB Memory | | Difference in mean change between Intervention and Control, per year | | Difference in intervention effect by biomarker level (randomization*BM*time), stratified analysis | | Difference in intervention effect by biomarker level (randomization*BM*time), main analysis | |
|---|---|---|---|---|---|---|---|---|---|
| | | **Intervention** | **Control** | **Estimate (95% CI)** | **p** | **Estimate (95% CI)** | **p** | **Estimate (95% CI)** | **p** |
| p-tau217 (Lumipulse) | Normal | 0.185 (0.013) | 0.168 (0.013) | 0.018 (-0.018 to 0.054) | 0.34 | 0.005 (-0.091 to 0.101) | 0.92 | 0.000 (-0.034 to 0.034) | 1.00 |
| | Intermediate or pathological | 0.103 (0.032) | 0.081 (0.032) | 0.023 (-0.066 to 0.111) | 0.62 | | | | |
| p-tau217 (Lumipulse) | ≤ median | 0.193 (0.017) | 0.170 (0.017) | 0.023 (-0.025 to 0.070) | 0.35 | -0.008 (-0.076 to 0.059) | 0.81 | 0.000 (-0.034 to 0.034) | 1.00 |
| | > median | 0.154 (0.017) | 0.140 (0.017) | 0.014 (-0.034 to 0.062) | 0.56 | | | | |
| p-tau181 | ≤ median | 0.188 (0.017) | 0.165 (0.017) | 0.024 (-0.024 to 0.072) | 0.33 | -0.013 (-0.080 to 0.055) | 0.72 | 0.005 (-0.029 to 0.040) | 0.76 |
| | > median | 0.159 (0.017) | 0.147 (0.017) | 0.011 (-0.036 to 0.059) | 0.64 | | | | |
| p-tau217 (NULISA) | ≤ median | 0.194 (0.017) | 0.181 (0.017) | 0.013 (-0.034 to 0.061) | 0.58 | 0.005 (-0.062 to 0.072) | 0.89 | 0.013 (-0.021 to 0.047) | 0.45 |
| | > median | 0.151 (0.017) | 0.133 (0.017) | 0.018 (-0.029 to 0.066) | 0.46 | | | | |
| p-tau231 | ≤ median | 0.198 (0.017) | 0.176 (0.017) | 0.022 (-0.026 to 0.069) | 0.37 | -0.007 (-0.074 to 0.060) | 0.84 | 0.008 (-0.026 to 0.042) | 0.64 |
| | > median | 0.150 (0.017) | 0.135 (0.017) | 0.014 (-0.033 to 0.062) | 0.55 | | | | |
| Aβ42 | ≤ median | 0.168 (0.018) | 0.151 (0.017) | 0.017 (-0.031 to 0.065) | 0.49 | -0.001 (-0.068 to 0.067) | 0.99 | 0.023 (-0.011 to 0.058) | 0.18 |
| | > median | 0.178 (0.017) | 0.161 (0.018) | 0.016 (-0.031 to 0.064) | 0.50 | | | | |
| Aβ40 | ≤ median | 0.190 (0.017) | 0.188 (0.017) | 0.002 (-0.045 to 0.050) | 0.93 | 0.032 (-0.035 to 0.099) | 0.35 | 0.032 (-0.003 to 0.066) | 0.07 |
| | > median | 0.157 (0.017) | 0.123 (0.017) | 0.034 (-0.013 to 0.082) | 0.16 | | | | |
| Aβ42/40 | ≤ median | 0.160 (0.017) | 0.114 (0.017) | 0.046 (-0.002 to 0.094) | 0.06 | -0.059 (-0.126 to 0.007) | 0.08 | -0.004 (-0.037 to 0.029) | 0.81 |
| | > median | 0.186 (0.017) | 0.199 (0.017) | -0.013 (-0.061 to 0.034) | 0.57 | | | | |
| GFAP | ≤ median | 0.186 (0.017) | 0.197 (0.017) | -0.010 (-0.057 to 0.037) | 0.67 | 0.059 (-0.008 to 0.125) | 0.09 | 0.022 (-0.012 to 0.056) | 0.21 |
| | > median | 0.160 (0.017) | 0.112 (0.017) | 0.048 (0.001 to 0.096) | 0.05 | | | | |
| NFL | ≤ median | 0.200 (0.017) | 0.188 (0.017) | 0.012 (-0.035 to 0.059) | 0.63 | 0.014 (-0.053 to 0.081) | 0.68 | 0.022 (-0.011 to 0.056) | 0.19 |
| | > median | 0.148 (0.017) | 0.122 (0.017) | 0.026 (-0.022 to 0.073) | 0.29 | | | | |

Estimates were obtained from linear mixed-effects models, using marginal prediction methods to derive interpretable effect estimates. Analyses were adjusted for baseline age, sex, education, study site, *APOE* genotype, body mass index (BMI), and estimated glomerular filtration rate (eGFR). Biomarker (BM) × randomization group × time interactions derived from the model where biomarkers were entered as continuous variables are considered the main results; estimates for intervention effects are additionally presented within biomarker strata (median split and predefined Lumipulse p-tau217 categories, normal <0.22 pg/ml vs. intermediate 0.22–0.34 pg/ml or pathological >0.34 pg/ml). Biomarker × randomization group × time interactions p>0.05 indicate that the intervention effects on cognition did not significantly differ by baseline biomarker levels.

Aβ=beta-amyloid; BM=biomarker; GFAP=glial fibrillary acidic protein; NFL=neurofilament light chain; NTB=neuropsychological test battery; p-tau=phosphorylated tau.

**Table 7**. Baseline biomarker levels and intervention effects on cognition (abbreviated memory)

| | Baseline level of biomarker | Mean change (SE) in NTB Abbreviated memory | | Difference in mean change between Intervention and Control, per year | | Difference in intervention effect by biomarker level (randomization*BM*time), stratified analysis | | Difference in intervention effect by biomarker level (randomization*BM*time), main analysis | |
|---|---|---|---|---|---|---|---|---|---|
| | | Intervention | Control | Estimate (95% CI) | p | Estimate (95% CI) | p | Estimate (95% CI) | p |
| p-tau217 (Lumipulse) | Normal | 0.130 (0.014) | 0.085 (0.014) | 0.045 (0.006 to 0.084) | **0.02** | -0.035 (-0.138 to 0.067) | 0.50 | 0.001 (-0.035 to 0.038) | 0.94 |
| | Intermediate or pathological | 0.062 (0.034) | 0.052 (0.035) | 0.010 (-0.085 to 0.105) | 0.84 | | | | |
| p-tau217 (Lumipulse) | ≤ median | 0.129 (0.018) | 0.090 (0.018) | 0.040 (-0.011 to 0.090) | 0.13 | 0.001 (-0.071 to 0.074) | 0.97 | 0.001 (-0.035 to 0.038) | 0.94 |
| | > median | 0.111 (0.019) | 0.070 (0.019) | 0.041 (-0.011 to 0.093) | 0.12 | | | | |
| p-tau181 | ≤ median | 0.125 (0.019) | 0.092 (0.018) | 0.034 (-0.018 to 0.085) | 0.20 | 0.009 (-0.064 to 0.081) | 0.82 | -0.006 (-0.043 to 0.031) | 0.76 |
| | > median | 0.116 (0.018) | 0.074 (0.019) | 0.042 (-0.009 to 0.093) | 0.11 | | | | |
| p-tau217 (NULISA) | ≤ median | 0.138 (0.018) | 0.104 (0.019) | 0.035 (-0.016 to 0.086) | 0.18 | 0.003 (-0.069 to 0.075) | 0.94 | 0.006 (-0.031 to 0.043) | 0.74 |
| | > median | 0.101 (0.019) | 0.063 (0.018) | 0.038 (-0.014 to 0.089) | 0.15 | | | | |
| p-tau231 | ≤ median | 0.128 (0.018) | 0.103 (0.018) | 0.025 (-0.025 to 0.076) | 0.33 | 0.025 (-0.047 to 0.098) | 0.49 | 0.003 (-0.034 to 0.040) | 0.89 |
| | > median | 0.113 (0.018) | 0.062 (0.019) | 0.051 (-0.001 to 0.102) | 0.05 | | | | |
| Aβ42 | ≤ median | 0.113 (0.019) | 0.073 (0.018) | 0.040 (-0.011 to 0.092) | 0.12 | -0.007 (-0.080 to 0.065) | 0.84 | 0.020 (-0.017 to 0.057) | 0.29 |
| | > median | 0.127 (0.018) | 0.094 (0.019) | 0.033 (-0.018 to 0.084) | 0.21 | | | | |
| Aβ40 | ≤ median | 0.126 (0.019) | 0.087 (0.018) | 0.039 (-0.012 to 0.090) | 0.14 | -0.002 (-0.074 to 0.071) | 0.96 | 0.007 (-0.030 to 0.044) | 0.71 |
| | > median | 0.115 (0.018) | 0.079 (0.019) | 0.037 (-0.015 to 0.088) | 0.16 | | | | |
| Aβ42/40 | ≤ median | 0.110 (0.019) | 0.067 (0.018) | 0.043 (-0.009 to 0.094) | 0.10 | -0.011 (-0.084 to 0.061) | 0.76 | 0.011 (-0.025 to 0.047) | 0.55 |
| | > median | 0.130 (0.018) | 0.099 (0.019) | 0.031 (-0.019 to 0.082) | 0.23 | | | | |
| GFAP | ≤ median | 0.138 (0.018) | 0.102 (0.018) | 0.036 (-0.015 to 0.087) | 0.16 | 0.006 (-0.066 to 0.078) | 0.87 | 0.003 (-0.033 to 0.040) | 0.86 |
| | > median | 0.103 (0.018) | 0.061 (0.019) | 0.042 ( -0.010 to 0.094) | 0.11 | | | | |
| NFL | ≤ median | 0.136 (0.018) | 0.091 (0.018) | 0.045 (-0.005 to 0.096) | 0.08 | -0.015 (-0.087 to 0.058) | 0.69 | 0.018 (-0.018 to 0.054) | 0.33 |
| | > median | 0.105 (0.018) | 0.074 (0.019) | 0.031 (-0.021 to 0.082) | 0.25 | | | | |

Estimates were obtained from linear mixed-effects models, using marginal prediction methods to derive interpretable effect estimates. Analyses were adjusted for baseline age, sex, education, study site, *APOE* genotype, body mass index (BMI), and estimated glomerular filtration rate (eGFR). Biomarker (BM) × randomization group × time interactions derived from the model where biomarkers were entered as continuous variables are considered the main results; estimates for intervention effects are additionally presented within biomarker strata (median split and predefined Lumipulse p-tau217 categories, normal <0.22 pg/ml vs. intermediate 0.22–0.34 pg/ml or pathological >0.34 pg/ml). Biomarker × randomization group × time interactions p>0.05 indicate that the intervention effects on cognition did not significantly differ by baseline biomarker levels.

Aβ=beta-amyloid; BM=biomarker; GFAP=glial fibrillary acidic protein; NFL=neurofilament light chain; NTB=neuropsychological test battery; p-tau=phosphorylated tau.

**Table 8**. Baseline biomarker levels and intervention effects on cognition (executive functioning)

| | Baseline level of biomarker | Mean change (SE) in NTB Executive functioning | | Difference in mean change between Intervention and Control, per year | | Difference in intervention effect by biomarker level (randomization*BM*time), stratified analysis | | Difference in intervention effect by biomarker level (randomization*BM*time), main analysis | |
|---|---|---|---|---|---|---|---|---|---|
| | | Intervention | Control | Estimate (95% CI) | p | Estimate (95% CI) | p | Estimate (95% CI) | p |
| p-tau217 (Lumipulse) | Normal | 0.054 (0.010) | 0.028 (0.010) | 0.026 (-0.003 to 0.055) | 0.08 | 0.001 (-0.076 to 0.078) | 0.98 | -0.008 (-0.035 to 0.019) | 0.56 |
| | Intermediate or pathological | 0.017 (0.025) | -0.010 (0.026) | 0.027 (-0.044 to 0.098) | 0.45 | | | | |
| p-tau217 (Lumipulse) | ≤ median | 0.074 (0.014) | 0.034 (0.013) | 0.040 (0.002 to 0.077) | **0.04** | -0.026 ( -0.080 to 0.027) | 0.34 | -0.008 (-0.035 to 0.019) | 0.56 |
| | > median | 0.024 (0.014) | 0.010 (0.014) | 0.013 (-0.025 to 0.051) | 0.49 | | | | |
| p-tau181 | ≤ median | 0.073 (0.014) | 0.028 (0.014) | 0.045 (0.007 to 0.083) | **0.02** | -0.035 (-0.089 to 0.019) | 0.20 | -0.007 (-0.035 to 0.020) | 0.60 |
| | > median | 0.025 (0.013) | 0.015 (0.014) | 0.010 (-0.028 to 0.048) | 0.60 | | | | |
| p-tau217 (NULISA) | ≤ median | 0.060 (0.013) | 0.041 (0.014) | 0.019 (-0.019 to 0.057) | 0.33 | 0.014 (-0.040 to 0.068) | 0.61 | 0.007 (-0.021 to 0.034) | 0.63 |
| | > median | 0.036 (0.014) | 0.003 (0.014) | 0.033 (-0.005 to 0.071) | 0.09 | | | | |
| p-tau231 | ≤ median | 0.074 (0.014) | 0.034 (0.014) | 0.040 (0.002 to 0.077) | **0.04** | -0.024 (-0.077 to 0.030) | 0.38 | -0.010 (-0.038 to 0.017) | 0.46 |
| | > median | 0.024 (0.013) | 0.008 (0.014) | 0.016 (-0.022 to 0.054) | 0.41 | | | | |
| Aβ42 | ≤ median | 0.063 (0.014) | 0.017 (0.013) | 0.047 (0.009 to 0.085) | **0.02** | -0.038 (-0.092 to 0.016) | 0.16 | -0.021 (-0.049 to 0.006) | 0.13 |
| | > median | 0.035 (0.013) | 0.027 (0.014) | 0.008 (-0.029 to 0.046) | 0.66 | | | | |
| Aβ40 | ≤ median | 0.052 (0.014) | 0.025 (0.014) | 0.027 (-0.011 to 0.065) | 0.16 | 0.000 (-0.054 to 0.053) | 0.99 | 0.000 (-0.027 to 0.027) | 1.00 |
| | > median | 0.045 (0.014) | 0.018 (0.014) | 0.027 (-0.011 to 0.065) | 0.17 | | | | |
| Aβ42/40 | ≤ median | 0.054 (0.014) | 0.016 (0.014) | 0.038 (0.000 to 0.076) | 0.05 | -0.022 (-0.075 to 0.032) | 0.43 | -0.016 (-0.042 to 0.011) | 0.25 |
| | > median | 0.043 (0.013) | 0.027 (0.014) | 0.016 (-0.021 to 0.054) | 0.40 | | | | |
| GFAP | ≤ median | 0.075 (0.014) | 0.027 (0.013) | 0.047 (0.010 to 0.085) | **0.01** | -0.040 (-0.094 to 0.013) | 0.14 | -0.017 (-0.044 to 0.011) | 0.23 |
| | > median | 0.022 (0.014) | 0.015 (0.014) | 0.007 (-0.031 to 0.046) | 0.70 | | | | |
| NFL | ≤ median | 0.066 (0.014) | 0.037 (0.013) | 0.029 (-0.009 to 0.067) | 0.13 | -0.003 (-0.056 to 0.051) | 0.92 | -0.010 (-0.037 to 0.017) | 0.46 |
| | > median | 0.031 (0.014) | 0.005 (0.014) | 0.026 (-0.012 to 0.064) | 0.18 | | | | |

Estimates were obtained from linear mixed-effects models, using marginal prediction methods to derive interpretable effect estimates. Analyses were adjusted for baseline age, sex, education, study site, *APOE* genotype, body mass index (BMI), and estimated glomerular filtration rate (eGFR). Biomarker (BM) × randomization group × time interactions derived from the model where biomarkers were entered as continuous variables are considered the main results; estimates for intervention effects are additionally presented within biomarker strata (median split and predefined Lumipulse p-tau217 categories, normal <0.22 pg/ml vs. intermediate 0.22–0.34 pg/ml or pathological >0.34 pg/ml). Biomarker × randomization group × time interactions p>0.05 indicate that the intervention effects on cognition did not significantly differ by baseline biomarker levels.

Aβ=beta-amyloid; BM=biomarker; GFAP=glial fibrillary acidic protein; NFL=neurofilament light chain; NTB=neuropsychological test battery; p-tau=phosphorylated tau.

**Table 9**. Baseline biomarker levels and intervention effects on cognition (processing speed)

| | Baseline level of biomarker | Mean change (SE) in NTB Processing speed | | Difference in mean change between Intervention and Control, per year | | Difference in intervention effect by biomarker level (randomization*BM*time), stratified analysis | | Difference in intervention effect by biomarker level (randomization*BM*time), main analysis | |
|---|---|---|---|---|---|---|---|---|---|
| | | **Intervention** | **Control** | **Estimate (95% CI)** | **p** | **Estimate (95% CI)** | **p** | **Estimate (95% CI)** | **p** |
| p-tau217 (Lumipulse) | Normal | 0.053 (0.011) | 0.026 (0.011) | 0.027 (-0.003 to 0.057) | 0.08 | 0.029 (-0.051 to 0.108) | 0.48 | -0.003 (-0.031 to 0.024) | 0.80 |
| | Intermediate or pathological | 0.035 (0.026) | -0.021 (0.027) | 0.056 (-0.018 to 0.129) | 0.14 | | | | |
| p-tau217 (Lumipulse) | ≤ median | 0.076 (0.014) | 0.042 (0.014) | 0.034 (-0.005 to 0.073) | 0.09 | -0.004 (-0.060 to 0.051) | 0.88 | -0.003 (-0.031 to 0.024) | 0.80 |
| | > median | 0.025 (0.014) | -0.005 (0.014) | 0.029 (-0.010 to 0.069) | 0.14 | | | | |
| p-tau181 | ≤ median | 0.070 (0.014) | 0.018 (0.014) | 0.052 (0.012 to 0.091) | **0.01** | -0.038 (-0.094 to 0.017) | 0.18 | -0.005 (-0.034 to 0.023) | 0.72 |
| | > median | 0.031 (0.014) | 0.018 (0.014) | 0.013 (-0.026 to 0.052) | 0.51 | | | | |
| p-tau217 (NULISA) | ≤ median | 0.071 (0.014) | 0.029 (0.014) | 0.043 (0.004 to 0.082) | **0.03** | -0.024 (-0.079 to 0.032) | 0.40 | -0.007 (-0.036 to 0.021) | 0.61 |
| | > median | 0.027 (0.014) | 0.008 (0.014) | 0.019 (-0.020 to 0.058) | 0.35 | | | | |
| p-tau231 | ≤ median | 0.077 (0.014) | 0.025 (0.014) | 0.052 (0.013 to 0.091) | **0.01** | -0.038 (-0.094 to 0.017) | 0.18 | -0.006 (-0.034 to 0.023) | 0.70 |
| | > median | 0.024 (0.014) | 0.011 (0.014) | 0.013 (-0.026 to 0.053) | 0.50 | | | | |
| Aβ42 | ≤ median | 0.044 (0.015) | 0.026 (0.014) | 0.019 (-0.021 to 0.058) | 0.35 | 0.027 (-0.029 to 0.082) | 0.35 | 0.011 (-0.017 to 0.039) | 0.45 |
| | > median | 0.055 (0.014) | 0.010 (0.015) | 0.045 (0.006 to 0.085) | **0.02** | | | | |
| Aβ40 | ≤ median | 0.050 (0.014) | 0.042 (0.014) | 0.008 (-0.031 to 0.047) | 0.69 | 0.049 (-0.007 to 0.104) | 0.08 | 0.015 (-0.014 to 0.043) | 0.31 |
| | > median | 0.050 (0.014) | -0.007 (0.014) | 0.057 (0.018 to 0.096) | **0.01** | | | | |
| Aβ42/40 | ≤ median | 0.032 (0.014) | 0.000 (0.014) | 0.032 (-0.007 to 0.072) | 0.11 | -0.002 (-0.057 to 0.054) | 0.95 | -0.003 (-0.031 to 0.025) | 0.83 |
| | > median | 0.067 (0.014) | 0.037 (0.014) | 0.030 (-0.009 to 0.069) | 0.13 | | | | |
| GFAP | ≤ median | 0.077 (0.014) | 0.033 (0.014) | 0.044 (0.005 to 0.083) | **0.03** | -0.022 (-0.077 to 0.034) | 0.44 | -0.004 (-0.032 to 0.024) | 0.78 |
| | > median | 0.024 (0.014) | 0.002 (0.014) | 0.022 (-0.017 to 0.062) | 0.27 | | | | |
| NFL | ≤ median | 0.083 (0.014) | 0.048 (0.014) | 0.036 (-0.003 to 0.074) | 0.07 | -0.005 ( -0.060 to 0.051) | 0.87 | -0.007 (-0.035 to 0.021) | 0.63 |
| | > median | 0.018 (0.014) | -0.013 (0.014) | 0.031 (-0.008 to 0.070) | 0.12 | | | | |

Estimates were obtained from linear mixed-effects models, using marginal prediction methods to derive interpretable effect estimates. Analyses were adjusted for baseline age, sex, education, study site, *APOE* genotype, body mass index (BMI), and estimated glomerular filtration rate (eGFR). Biomarker (BM) × randomization group × time interactions derived from the model where biomarkers were entered as continuous variables are considered the main results; estimates for intervention effects are additionally presented within biomarker strata (median split and predefined Lumipulse p-tau217 categories, normal <0.22 pg/ml vs. intermediate 0.22–0.34 pg/ml or pathological >0.34 pg/ml). Biomarker × randomization group × time interactions p>0.05 indicate that the intervention effects on cognition did not significantly differ by baseline biomarker levels.

Aβ=beta-amyloid; BM=biomarker; GFAP=glial fibrillary acidic protein; NFL=neurofilament light chain; NTB=neuropsychological test battery; p-tau=phosphorylated tau.

**Table 10.** Transitions in p-tau217 status (Lumipulse) from baseline to two years

| Status: Baseline → 2 years | Pooled, estimated % | Intervention group, estimated % | Control group, estimated % | Intervention-Control difference, estimate (95% CI) | p |
|---|---|---|---|---|---|
| Normal → Normal | 93.9% | 93.3% | 94.5% | -1.2 (-4.1 to 1.7) | 0.42 |
| Normal → Intermediate | 4.5% | 5.1% | 4.0% | 1.1 (-1.4 to 3.7) | 0.38 |
| Normal → Pathological | 1.6% | 1.6% | 1.5% | 0.1 (-1.4 to 1.5) | 0.94 |
| Intermediate → Intermediate | 52.5% | 44.9% | 59.0% | -14.1 (-37.6 to 9.4) | 0.24 |
| Intermediate → Pathological | 23.6% | 33.1% | 16.2% | 16.9 (-2.5 to 36.3) | 0.09 |
| Intermediate → Normal | 23.8% | 22.0% | 24.8% | -2.8 (-24.4 to 18.9) | 0.80 |
| Pathological → Pathological | 76.1% | 78.5% | 73.1% | 5.4 (-15.7 to 26.5) | 0.62 |
| Pathological → Intermediate | 4.5% | 5.6% | 3.6% | 2.0 (-6.2 to 10.3) | 0.63 |
| Pathological → Normal | 19.4% | 15.9% | 23.3% | -7.4 (-27.5 to 12.7) | 0.47 |

Estimated transitions in Lumipulse p-tau217 status from baseline to two years from logistic regression model, including participants with both biomarker measurement points. Analysis was adjusted for baseline age, sex, education, site, *APOE*, baseline body mass index (BMI), baseline estimated glomerular filtration rate (eGFR), 2-year eGFR change and randomization group* eGFR change. Cut-offs for p-tau217 status: normal <0.22 pg/ml, intermediate 0.22–0.34 pg/ml, pathological >0.34 pg/ml.

**Table 11.** Change in biomarker levels in the intervention and control groups (model including BMI change)

| | **Estimated mean change from baseline, Intervention group** | | | **Estimated mean change from baseline, Control group** | | | **Intervention-Control difference in the estimated mean change** | | |
|---|---|---|---|---|---|---|---|---|---|
| | Estimate (95% CI) | SE | p | Estimate (95% CI) | SE | p | Estimate (95% CI) | SE | p |
| **p-tau217 (Lumipulse)** | 0.010 (0.007 to 0.014) | 0.002 | **<0.001** | 0.009 (0.005 to 0.012) | 0.002 | **<0.001** | 0.002 (-0.003 to 0.006) | 0.002 | 0.50 |
| **p-tau181** | 0.072 (0.024 to 0.119) | 0.024 | **0.003** | 0.072 (0.025 to 0.119) | 0.024 | **0.003** | -0.000 (-0.066 to 0.065) | 0.033 | 0.99 |
| **p-tau217 (NULISA)** | 0.079 (0.042 to 0.116) | 0.019 | **<0.001** | 0.049 (0.012 to 0.085) | 0.019 | **0.01** | 0.030 (-0.021 to 0.081) | 0.026 | 0.24 |
| **p-tau231** | 0.066 (0.021 to 0.111) | 0.023 | **0.004** | 0.065 (0.021 to 0.109) | 0.023 | **0.004** | 0.001 (-0.060 to 0.063) | 0.032 | 0.96 |
| **Aβ42** | 0.031 (-0.014 to 0.077) | 0.023 | 0.17 | 0.062 (0.017 to 0.107) | 0.023 | **0.01** | -0.030 (-0.093 to 0.032) | 0.032 | 0.34 |
| **Aβ40** | -0.026 (-0.071 to 0.018) | 0.023 | 0.25 | -0.018 (-0.062 to 0.026) | 0.022 | 0.42 | -0.008 (-0.070 to 0.053) | 0.031 | 0.79 |
| **Aβ42/40** | 0.085 (0.028 to 0.142) | 0.029 | **0.004** | 0.086 (0.030 to 0.143) | 0.029 | **0.003** | -0.002 (-0.080 to 0.077) | 0.040 | 0.97 |
| **GFAP** | 0.119 (0.080 to 0.158) | 0.020 | **<0.001** | 0.125 (0.086 to 0.164) | 0.020 | **<0.001** | -0.006 (-0.060 to 0.048) | 0.028 | 0.82 |
| **NFL** | 0.106 (0.057 to 0.155) | 0.025 | **<0.001** | 0.086 (0.037 to 0.134) | 0.025 | **<0.001** | 0.021 (-0.047 to 0.088) | 0.034 | 0.55 |

Estimates represent biomarker change from baseline to 2 years in the intervention and control groups and the group differences in these changes. Intervention group n=553 (Lumipulse p-tau217) and n=554 (other biomarkers); control group n=548 (Lumipulse p-tau217) and n=550 (other biomarkers). Estimates were obtained from linear mixed-effects models, using marginal prediction methods to derive interpretable effect estimates. Positive estimates indicate increase in biomarker levels. Biomarkers were analyzed on a transformed scale; estimates are presented on the original scale. Analyses were adjusted for baseline age, sex, education, study site, *APOE* genotype, body mass index (BMI), estimated glomerular filtration rate (eGFR), 2-year change in eGFR, randomization group*eGFR change interaction, and 2-year change in BMI.

Aβ=beta-amyloid; GFAP=glial fibrillary acidic protein; NFL=neurofilament light chain; p-tau=phosphorylated tau.

**Table 12.** Changes in biomarker levels by the level of adherence to intervention

| | **Estimated mean change from baseline, Adherent participants (n=231)** | | | **Estimated mean change from baseline, Partially adherent participants (n=246)** | | | **Estimated mean change from baseline, Non-adherent participants (n=77)** | | | **Estimated mean change from baseline, Control group (n=550)** | | |
|---|---|---|---|---|---|---|---|---|---|---|---|---|
| | Estimate (95% CI) | SE | p | Estimate (95% CI) | SE | p | Estimate (95% CI) | SE | p | Estimate (95% CI) | SE | p |
| **p-tau217 (Lumipulse)** | 0.008 (0.003 to 0.014) | 0.003 | **0.002** | 0.016 (0.010 to 0.021) | 0.003 | **<0.001** | 0.009 (0.001 to 0.017) | 0.004 | **0.03** | 0.010 (0.006 to 0.013) | 0.002 | **<0.001** |
| **p-tau181** | 0.092 (0.019 to 0.166) | 0.038 | **0.01** | 0.088 (0.019 to 0.157) | 0.035 | **0.01** | 0.037 (-0.079 to 0.153) | 0.060 | 0.53 | 0.079 (0.033 to 0.126) | 0.024 | **0.001** |
| **p-tau217 (NULISA)** | 0.103 (0.046 to 0.160) | 0.029 | **<0.001** | 0.087 (0.033 to 0.141) | 0.027 | **0.001** | 0.072 (-0.017 to 0.161) | 0.045 | 0.11 | 0.057 (0.021 to 0.093) | 0.018 | **0.002** |
| **p-tau231** | 0.073 (0.003 to 0.142) | 0.036 | **0.04** | 0.100 (0.034 to 0.164) | 0.033 | **0.003** | 0.049 (-0.061 to 0.158) | 0.056 | 0.38 | 0.075 (0.032 to 0.119) | 0.022 | **0.001** |
| **Aβ42** | 0.074 (0.005 to 0.144) | 0.036 | **0.04** | 0.010 (-0.055 to 0.075) | 0.033 | 0.76 | 0.016 (-0.094 to 0.126) | 0.056 | 0.77 | 0.066 (0.022 to 0.110) | 0.022 | **0.003** |
| **Aβ40** | 0.005 (-0.064 to 0.075) | 0.035 | 0.88 | -0.053 (-0.117 to 0.011) | 0.035 | 0.11 | -0.004 (-0.112 to 0.105) | 0.056 | 0.95 | -0.015 (-0.058 to 0.028) | 0.022 | 0.50 |
| **Aβ42/40** | 0.124 (0.036 to 0.211) | 0.045 | **0.006** | 0.076 (-0.006 to 0.158) | 0.042 | 0.07 | 0.019 (-0.119 to 0.157) | 0.071 | 0.79 | 0.088 (0.032 to 0.143) | 0.028 | **0.002** |
| **GFAP** | 0.152 (0.090 to 0.213) | 0.031 | **<0.001** | 0.124 (0.068 to 0.180) | 0.029 | **<0.001** | 0.105 (0.100 to 0.200) | 0.048 | **0.03** | 0.135 (0.096 to 0.173) | 0.020 | **<0.001** |
| **NFL** | 0.154 (0.078 to 0.231) | 0.039 | **<0.001** | 0.105 (0.035 to 0.176) | 0.036 | **0.003** | 0.121 (0.003 to 0.240) | 0.060 | **0.04** | 0.100 (0.053 to 0.148) | 0.024 | **<0.001** |

Estimates represent biomarker change from baseline to 2 years in the intervention group, split by the level of adherence to intervention, and the control group. Estimates were obtained from linear mixed-effects models, using marginal prediction methods to derive interpretable effect estimates. Positive estimates indicate increase in biomarker levels. Biomarkers were analyzed on a transformed scale; estimates are presented on the original scale. Analyses were adjusted for baseline age, sex, education, study site, *APOE* genotype, body mass index (BMI), estimated glomerular filtration rate (eGFR), 2-year change in eGFR, and randomization group*eGFR change interaction. All differences between the groups were statistically non-significant p>0.05.

Adherence reflects participation in scheduled intervention activities (calculated only for the intervention group) and is based on prespecified definition. Completion of the individual and group-based sessions in the different intervention domains (diet, exercise, cognitive training, vascular intervention) were summarized to an overall adherence score (range 0-8). Non-adherence is defined as 0-5 points, partial adherence as 6-7 points, and adherence as 8 points. Details on the calculations are provided in Ngandu T, Lehtisalo J, Korkki S, et al. The effect of adherence on cognition in a multidomain lifestyle intervention (FINGER). *Alzheimer's Dement*. 2022;18:1325–1334.

Aβ=beta-amyloid; GFAP=glial fibrillary acidic protein; NFL=neurofilament light chain; p-tau=phosphorylated tau.

**Table 13.** Intervention effect on biomarker levels, by *APOE*4 status

| | Intervention-Control difference in the estimated mean change in biomarker levels *APOE*4 carriers | | Intervention-Control difference in the estimated mean change in biomarker levels *APOE*4 non-carriers | | Difference in the intervention effect between *APOE*4 carriers and non-carriers | |
|---|---|---|---|---|---|---|
| | Estimate (95% CI) | p | Estimate (95% CI) | p | Estimate (95% CI) | p |
| **p-tau217 (Lumipulse)** | 0.004 (-0.009 to 0.016) | 0.58 | 0.003 (-0.003 to 0.008) | 0.33 | 0.001 (-0.012 to 0.014) | 0.89 |
| **p-tau181** | -0.019 (-0.136 to 0.098) | 0.75 | 0.013 (-0.070 to 0.096) | 0.76 | -0.032 (-0.173 to 0.109) | 0.66 |
| **p-tau217 (NULISA)** | 0.017 (-0.078 to 0.111) | 0.73 | 0.040 (-0.022 to 0.103) | 0.21 | -0.023 (-0.136 to 0.089) | 0.68 |
| **p-tau231** | -0.011 (-0.121 to 0.099) | 0.84 | 0.018 (-0.061 to 0.096) | 0.66 | -0.029 (-0.162 to 0.104) | 0.67 |
| **Aβ42** | -0.017 (-0.128 to 0.093) | 0.76 | -0.022 (-0.100 to 0.056) | 0.58 | 0.005 (-0.129 to 0.138) | 0.95 |
| **Aβ40** | 0.069 (-0.038 to 0.176) | 0.21 | -0.038 (-0.115 to 0.039) | 0.34 | 0.107 (-0.239 to 0.237) | 0.11 |
| **Aβ42/40** | -0.087 (-0.227 to 0.053) | 0.22 | 0.047 (-0.053 to 0.146) | 0.36 | -0.134 (-0.303 to 0.036) | 0.12 |
| **GFAP** | 0.040 (-0.053 to 0.133) | 0.40 | -0.019 (-0.088 to 0.050) | 0.60 | 0.059 (-0.055 to 0.173) | 0.31 |
| **NFL** | -0.003 (-0.120 to 0.115) | 0.97 | 0.054 (-0.030 to 0.138) | 0.21 | -0.057 (-0.199 to 0.086) | 0.44 |

Estimates were obtained from linear mixed-effects models, using marginal prediction methods to derive interpretable effect estimates. Model included 3-way *APOE**randomization group*time interaction to study the impact of *APOE*4 status on intervention effects on biomarker changes from baseline to 2 years. *APOE4* carriers n=340, non-carriers n=694 (Lumipulse p-tau217) or n=696 (other biomarkers). Biomarkers were analyzed on a transformed scale; estimates are presented on the original scale. Analyses were adjusted for baseline age, sex, education, study site, body mass index (BMI), estimated glomerular filtration rate (eGFR), 2-year change in eGFR, and randomization group*eGFR change interaction.

Aβ=beta-amyloid; GFAP=glial fibrillary acidic protein; NFL=neurofilament light chain; p-tau=phosphorylated tau.

**Figure 1.** Predictive performance for Aβ-PET positivity (models including age)

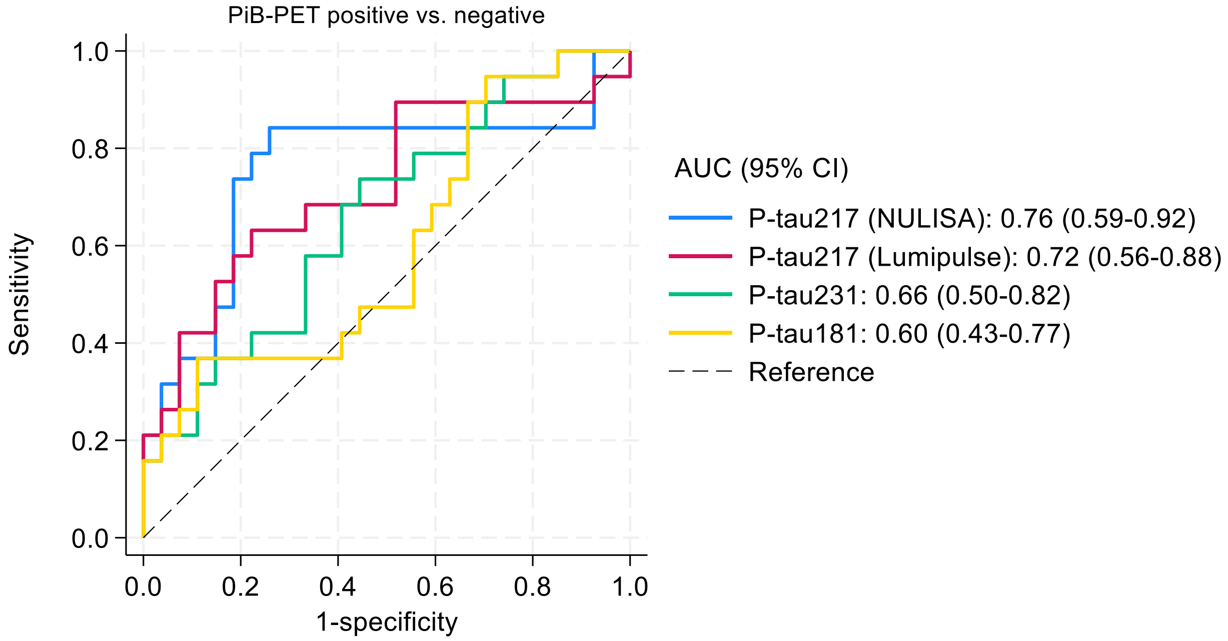

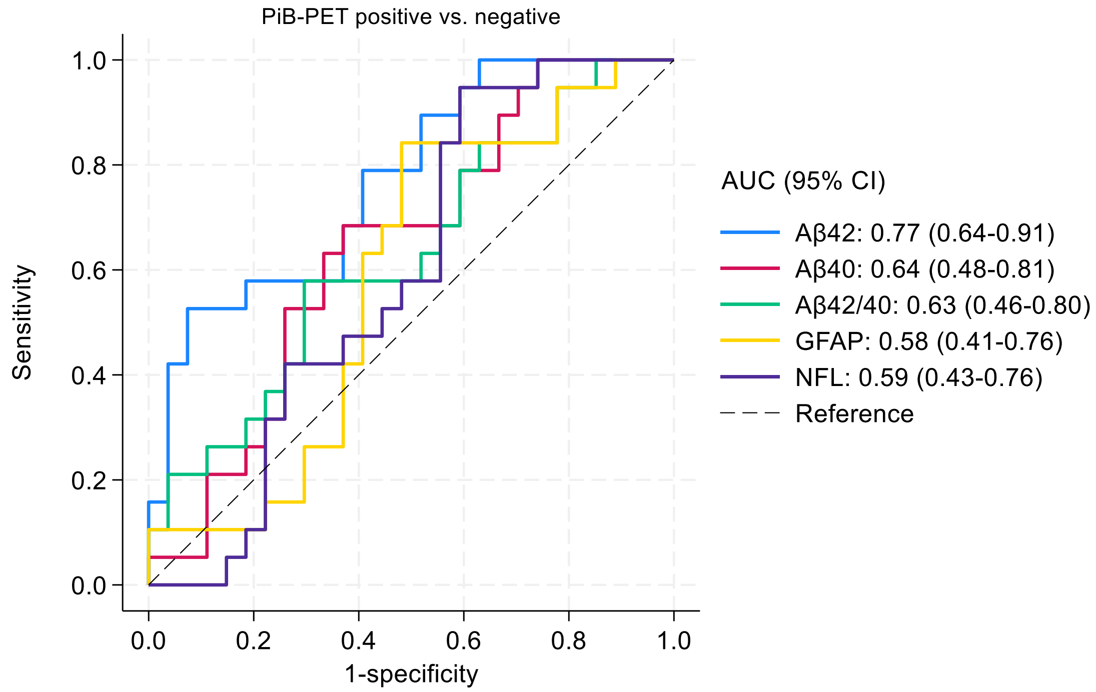

Logistic regression models including biomarkers (continuous variables) and participant age; discrimination quantified by AUCs (95% CI). PiB-PET scan and visual read of Aβ positivity available for 46 participants at baseline. P-tau217 measured with both Lumipulse and NULISAseq™ CNS Disease Panel 120, all other markers with NULISAseq™ CNS Disease Panel 120.

Aβ=beta-amyloid; AUC=area under the curve; GFAP=glial fibrillary acidic protein; NFL=neurofilament light chain; PiB-PET=Pittsburgh compound B positron emission tomography; p-tau=phosphorylated tau.

**Figure 2.** Associations between baseline biomarker levels and cognition (memory)

**A. Baseline cognition**

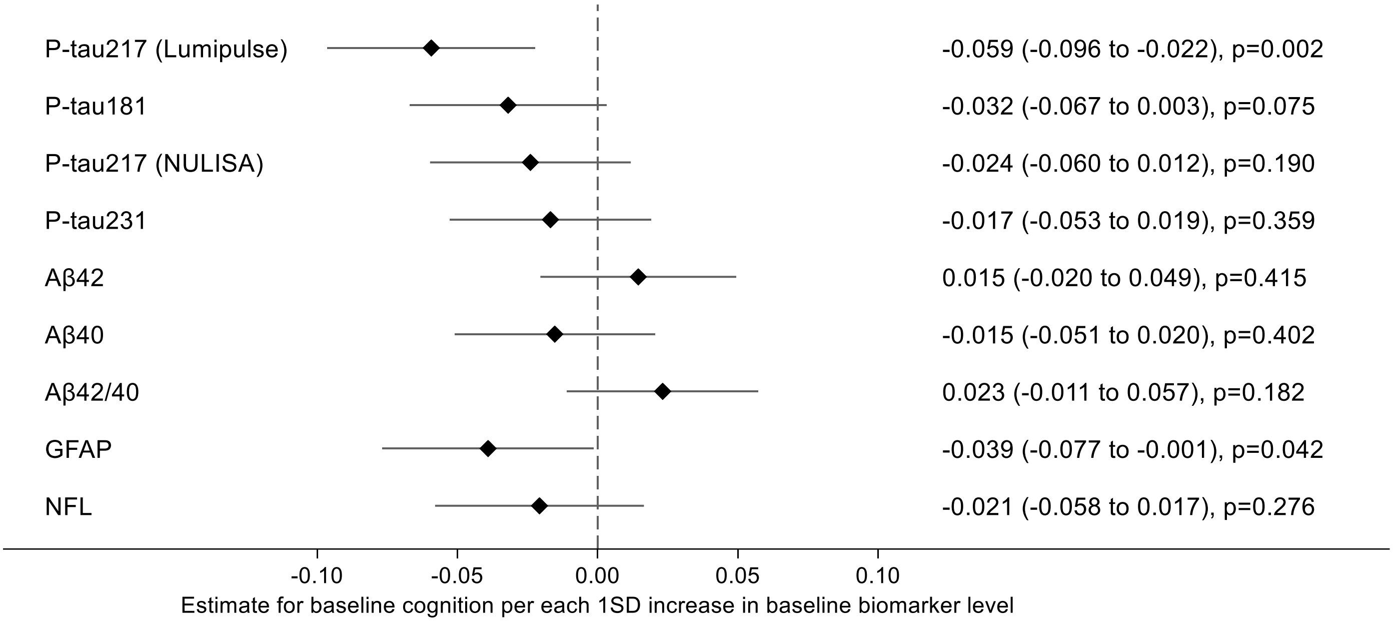

**B. Cognitive change over two years**

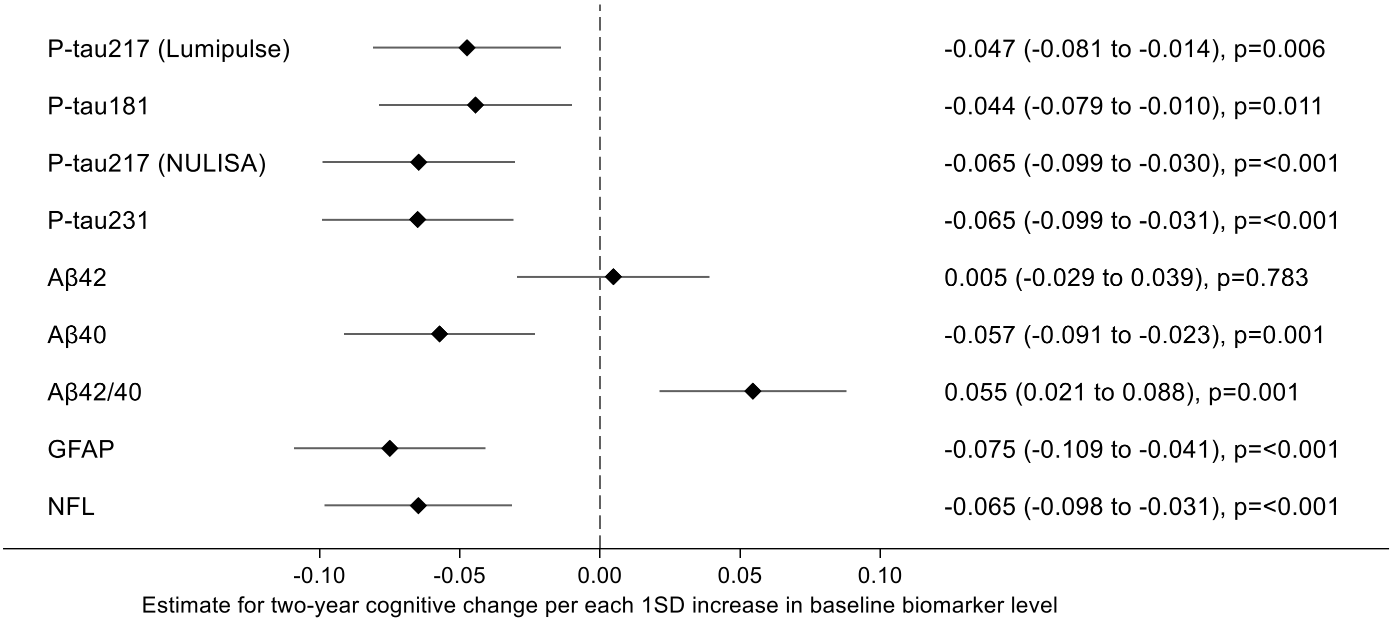

Outcome NTB Memory. Estimates (95% CI) were obtained from linear regression models (for baseline cognition, A) and mixed-effects models (for cognitive change, B), using marginal prediction methods to derive interpretable effect estimates. Results are interpreted as differences in baseline cognitive performance (A) or cognitive change in two years (B), per 1 standard deviation (SD) higher baseline biomarker level, in the whole cohort (irrespective of randomization). Negative estimates indicate worse cognitive performance or less favorable cognitive trajectory (less improvement) with increasing baseline biomarker levels. Analyses were adjusted for baseline age, sex, education, study site, *APOE* genotype, BMI, and eGFR. Baseline analysis n=1220 (Lumipulse p-tau217) and n=1223 (NULISA markers); longitudinal analysis n=1154 (Lumipulse) and n=1157 (NULISA).

Aβ=beta-amyloid; GFAP=glial fibrillary acidic protein; NFL=neurofilament light chain; NTB=neuropsychological test battery; p-tau=phosphorylated tau.

**Figure 3.** Associations between baseline biomarker levels and cognition (abbreviated memory)

**A. Baseline cognition**

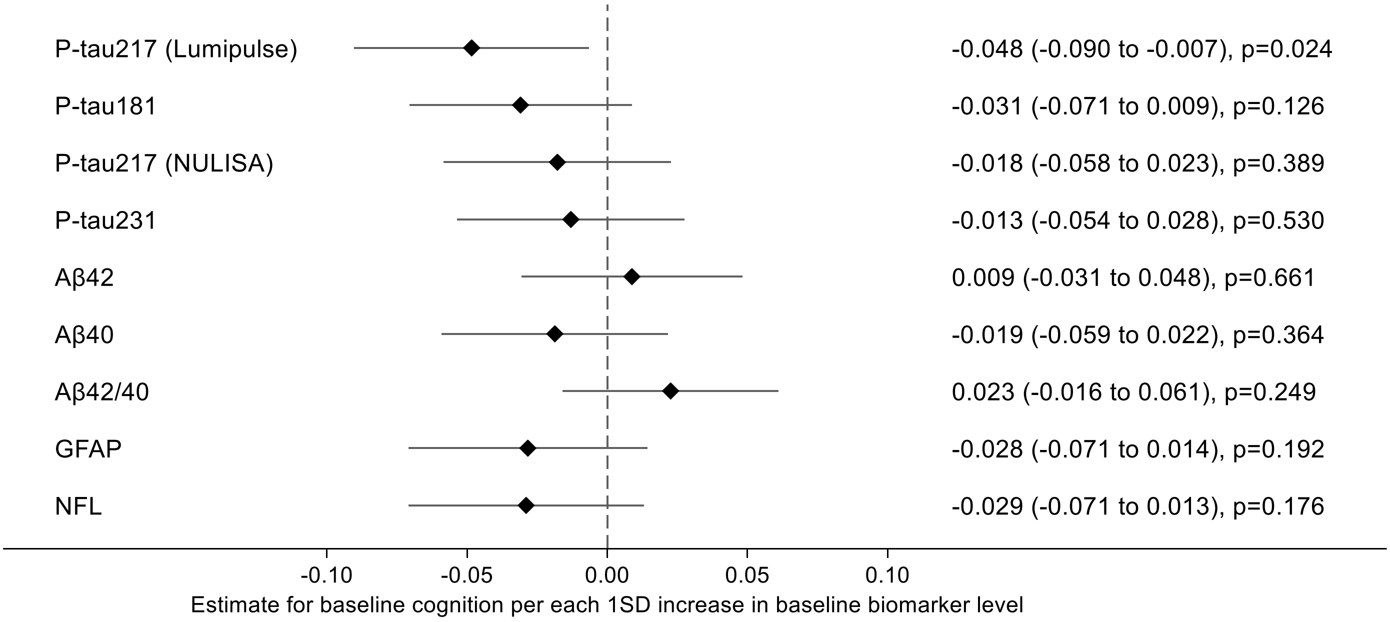

**B. Cognitive change over two years**

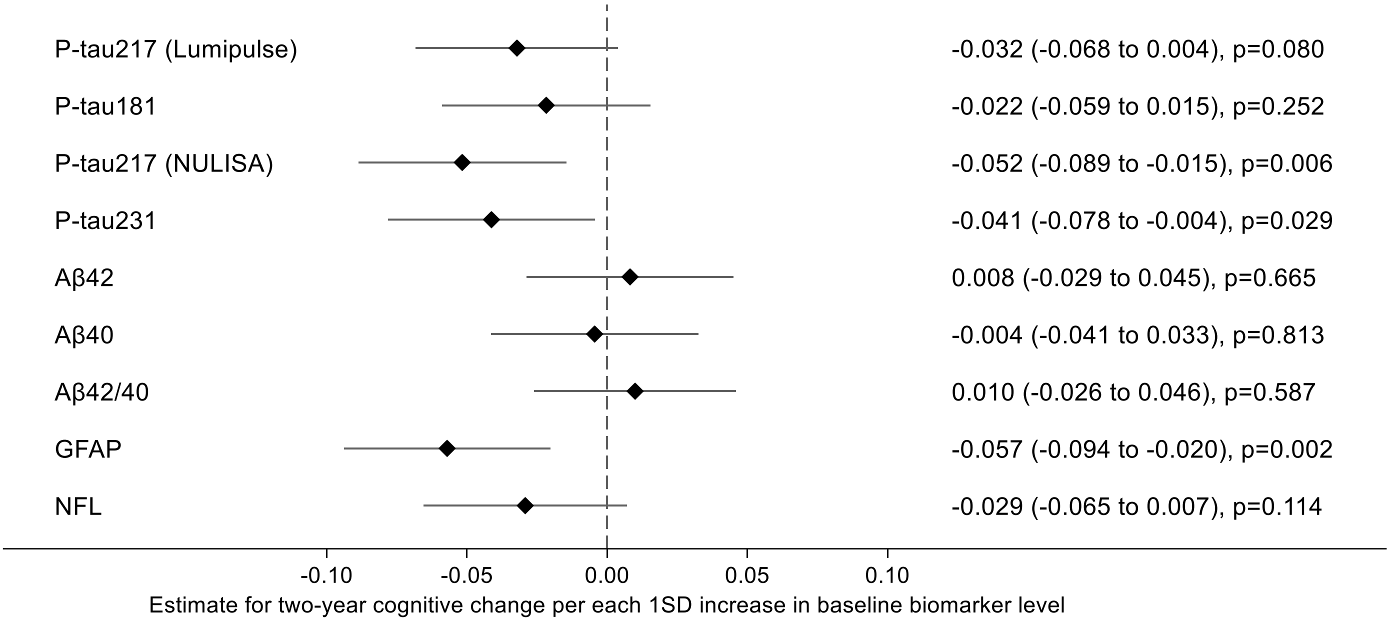

Outcome NTB Abbreviated memory. Estimates (95% CI) were obtained from linear regression models (for baseline cognition, A) and mixed-effects models (for cognitive change, B), using marginal prediction methods to derive interpretable effect estimates. Results are interpreted as differences in baseline cognitive performance (A) or cognitive change in two years (B), per 1 standard deviation (SD) higher baseline biomarker level, in the whole cohort (irrespective of randomization). Negative estimates indicate worse cognitive performance or less favorable cognitive trajectory (less improvement) with increasing baseline biomarker levels. Analyses were adjusted for baseline age, sex, education, study site, *APOE* genotype, BMI, and eGFR. Baseline analysis n=1199 (Lumipulse p-tau217) and n=1202 (NULISA markers); longitudinal analysis n=1153 (Lumipulse) and n=1156 (NULISA).

Aβ=beta-amyloid; GFAP=glial fibrillary acidic protein; NFL=neurofilament light chain; NTB=neuropsychological test battery; p-tau=phosphorylated tau.

**Figure 4.** Associations between baseline biomarker levels and cognition (executive functioning)

**A. Baseline cognition**

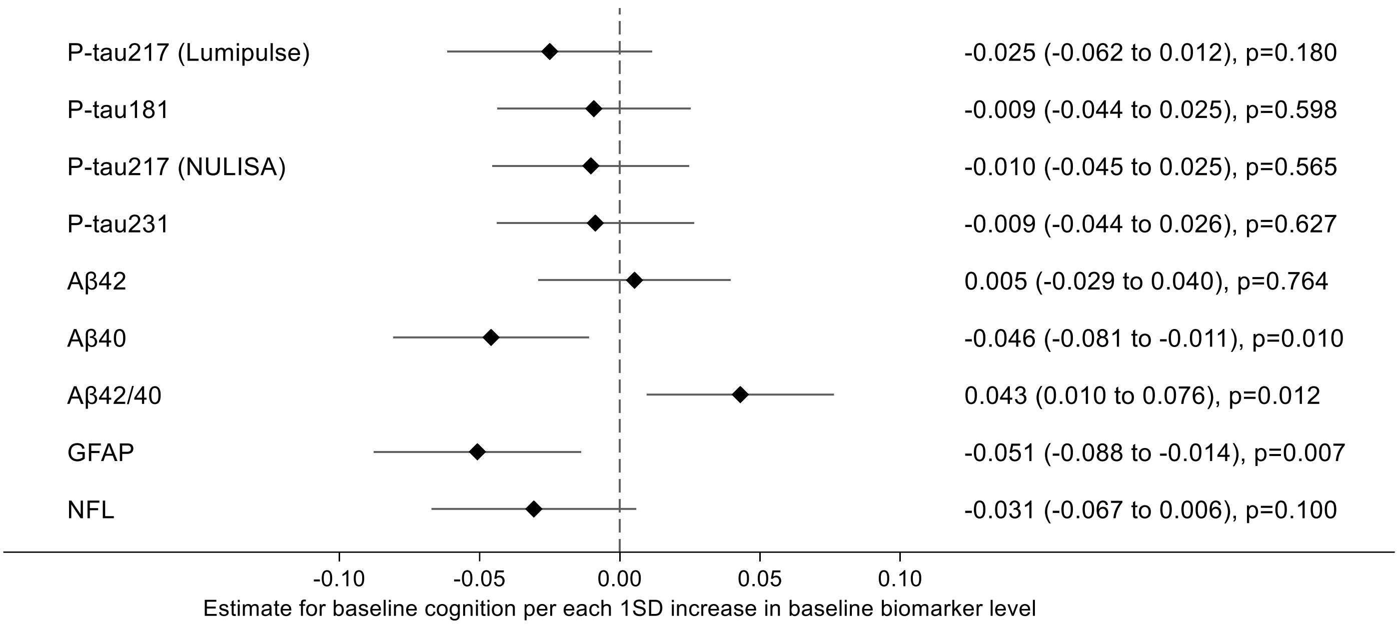

**B. Cognitive change over two years**

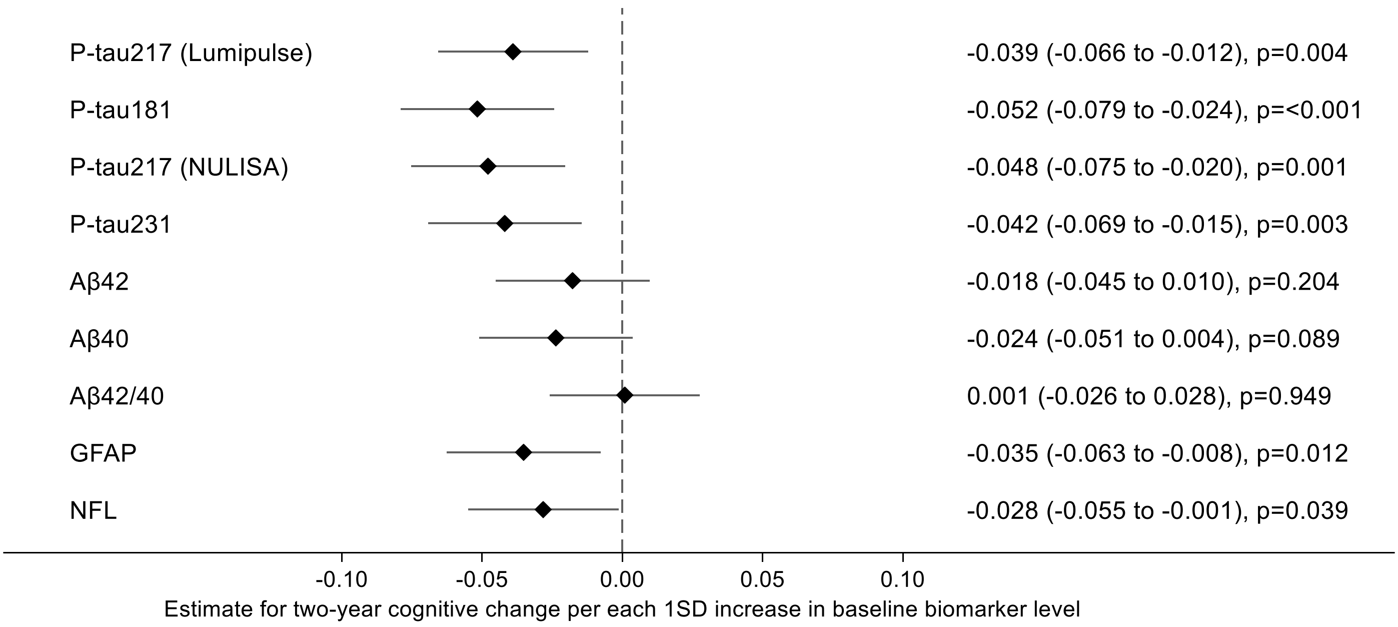

Outcome NTB Executive functioning. Estimates (95% CI) were obtained from linear regression models (for baseline cognition, A) and mixed-effects models (for cognitive change, B), using marginal prediction methods to derive interpretable effect estimates. Results are interpreted as differences in baseline cognitive performance (A) or cognitive change in two years (B), per 1 standard deviation (SD) higher baseline biomarker level, in the whole cohort (irrespective of randomization). Negative estimates indicate worse cognitive performance or less favorable cognitive trajectory (less improvement) with increasing baseline biomarker levels. Analyses were adjusted for baseline age, sex, education, study site, *APOE* genotype, BMI, and eGFR. Baseline analysis n=1219 (Lumipulse p-tau217) and n=1222 (NULISA markers); longitudinal analysis n=1154 (Lumipulse) and n=1157 (NULISA).

Aβ=beta-amyloid; GFAP=glial fibrillary acidic protein; NFL=neurofilament light chain; NTB=neuropsychological test battery; p-tau=phosphorylated tau.

**Figure 5.** Associations between baseline biomarker levels and cognition (processing speed)

**A. Baseline cognition**

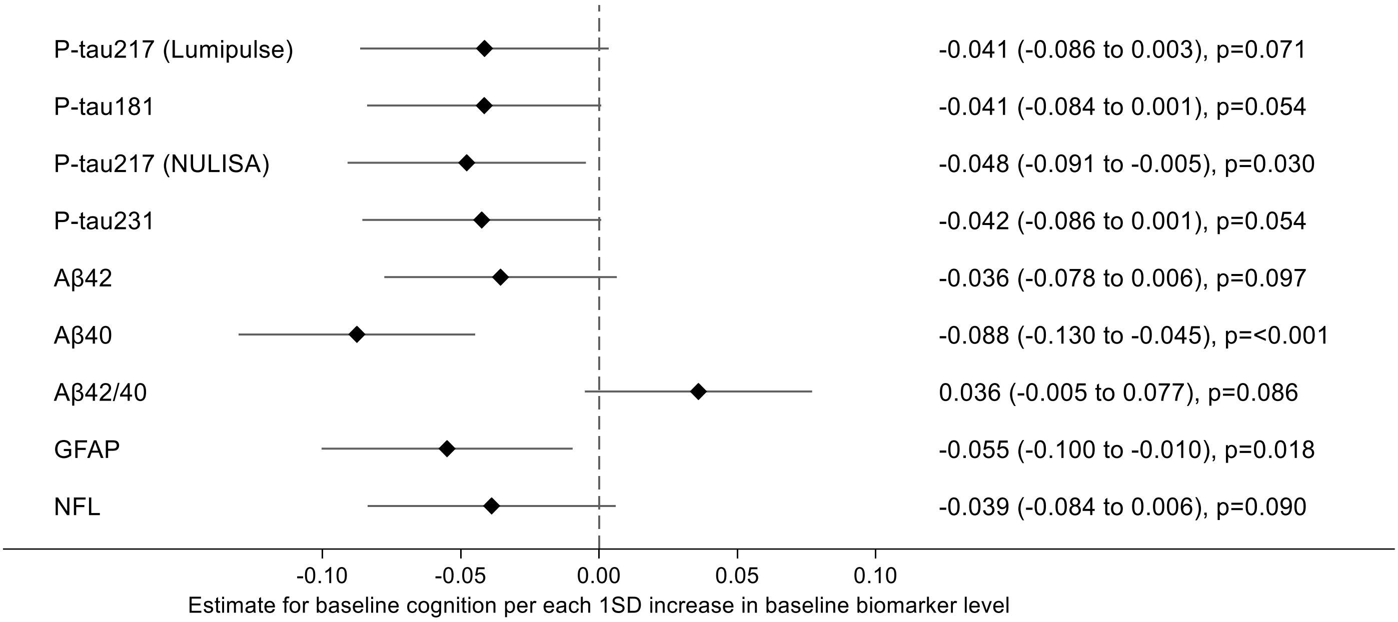

**B. Cognitive change over two years**

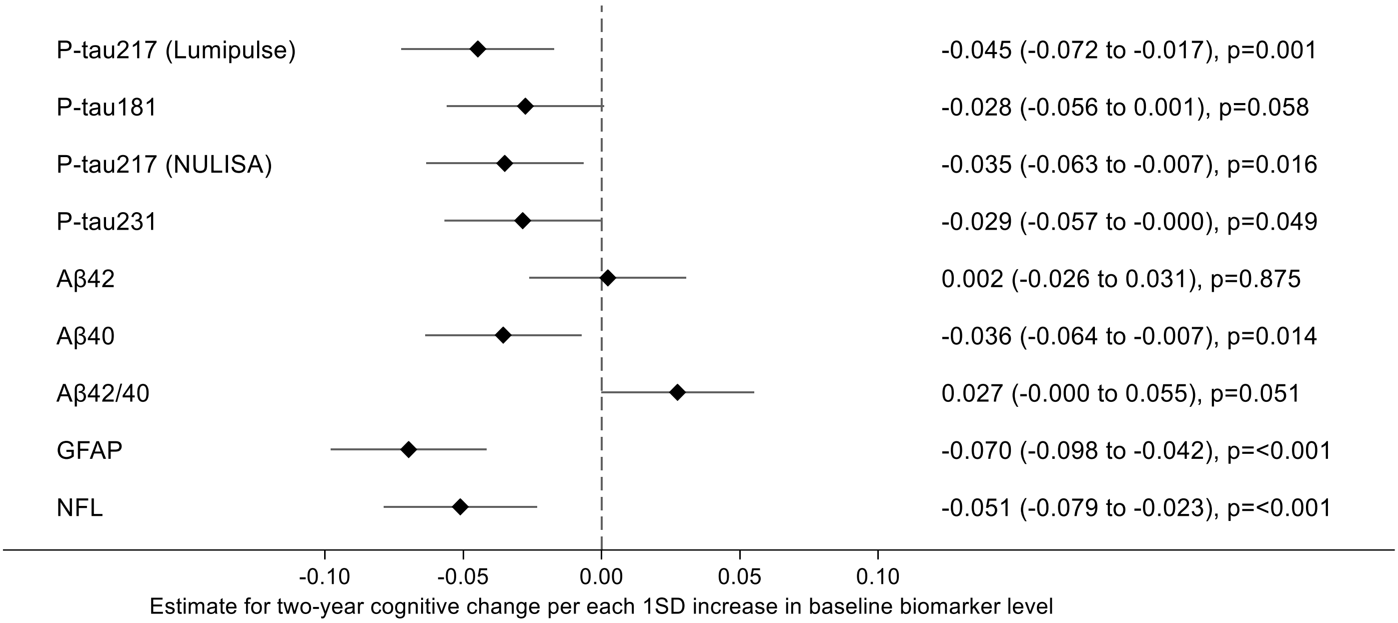

Outcome NTB Processing speed. Estimates (95% CI) were obtained from linear regression models (for baseline cognition, A) and mixed-effects models (for cognitive change, B), using marginal prediction methods to derive interpretable effect estimates. Results are interpreted as differences in baseline cognitive performance (A) or cognitive change in two years (B), per 1 standard deviation (SD) higher baseline biomarker level, in the whole cohort (irrespective of randomization). Negative estimates indicate worse cognitive performance or less favorable cognitive trajectory (less improvement) with increasing baseline biomarker levels. Analyses were adjusted for baseline age, sex, education, study site, *APOE* genotype, BMI, and eGFR. Baseline analysis n=1220 (Lumipulse p-tau217) and n=1223 (NULISA markers); longitudinal analysis n=1154 (Lumipulse) and n=1157 (NULISA).

Aβ=beta-amyloid; GFAP=glial fibrillary acidic protein; NFL=neurofilament light chain; NTB=neuropsychological test battery; p-tau=phosphorylated tau

**Figure 6.** Baseline associations between biomarker levels and cognition, by *APOE4* status

#### A. Global cognition

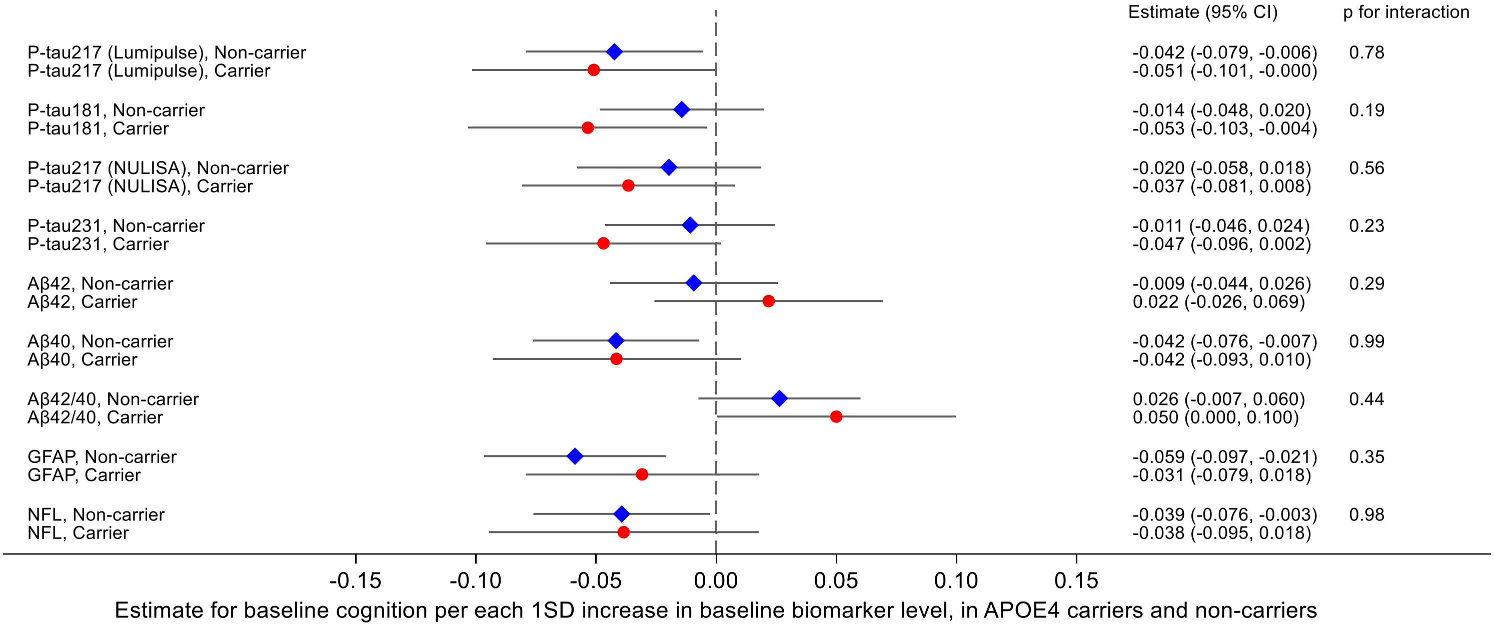

#### B. Memory

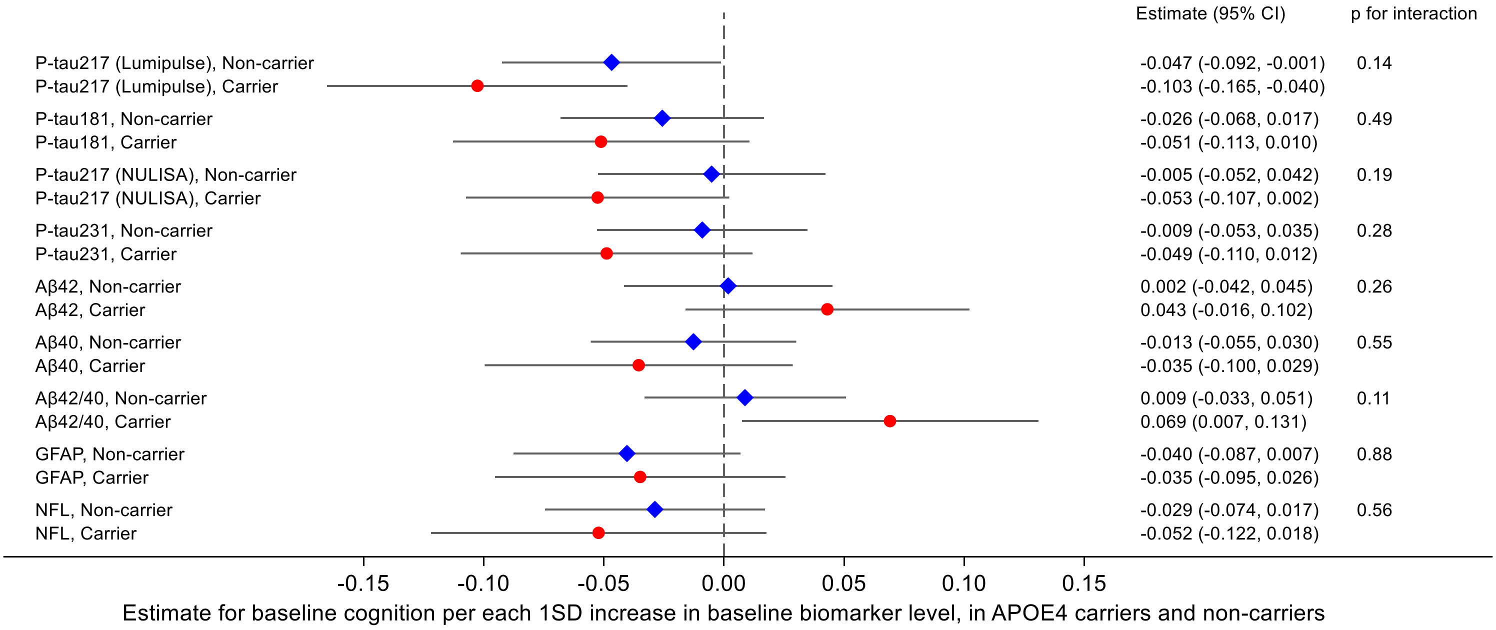

### C. Abbreviated memory

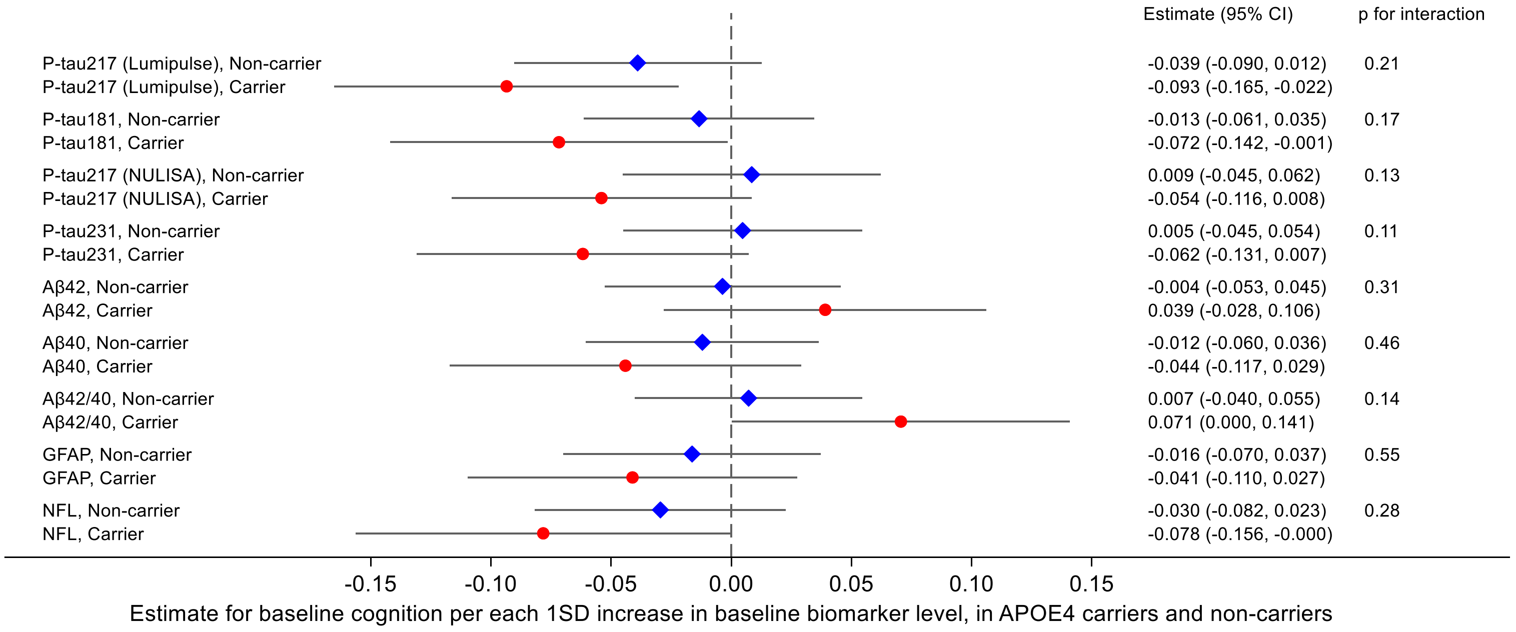

### D. Executive functioning

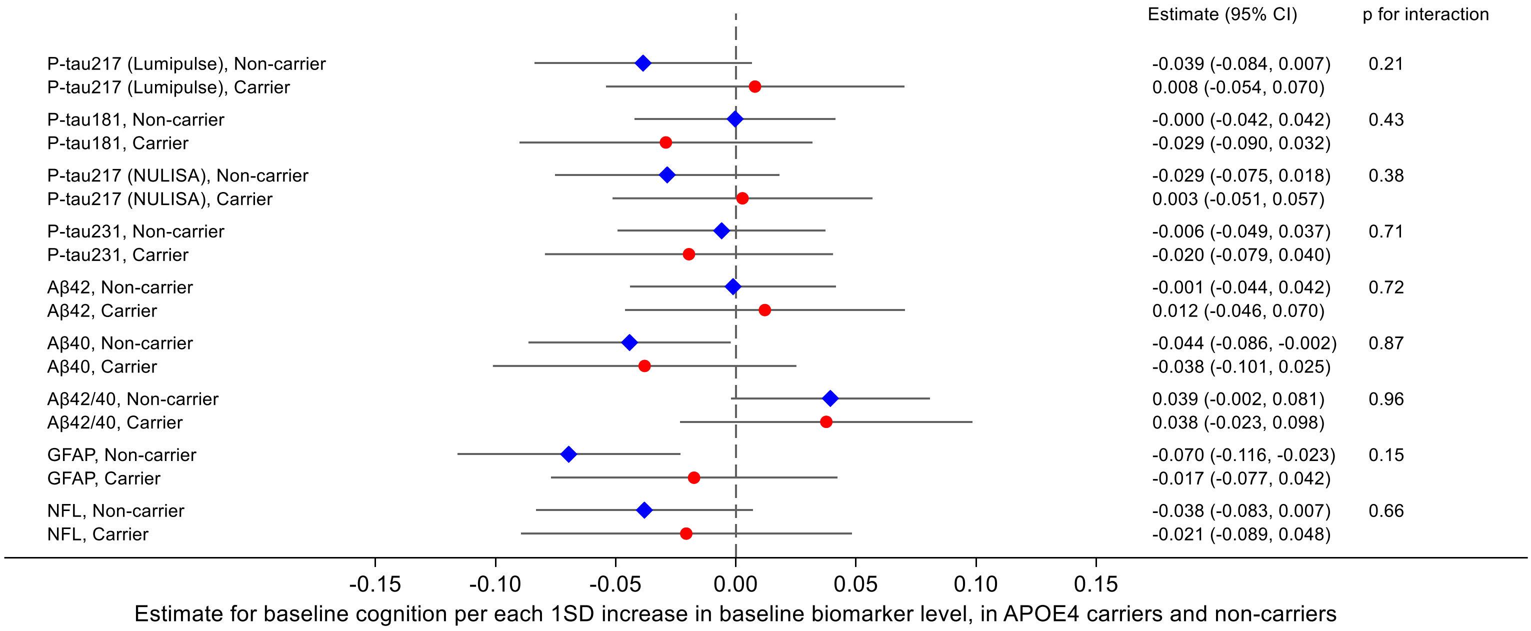

**E. Processing speed**

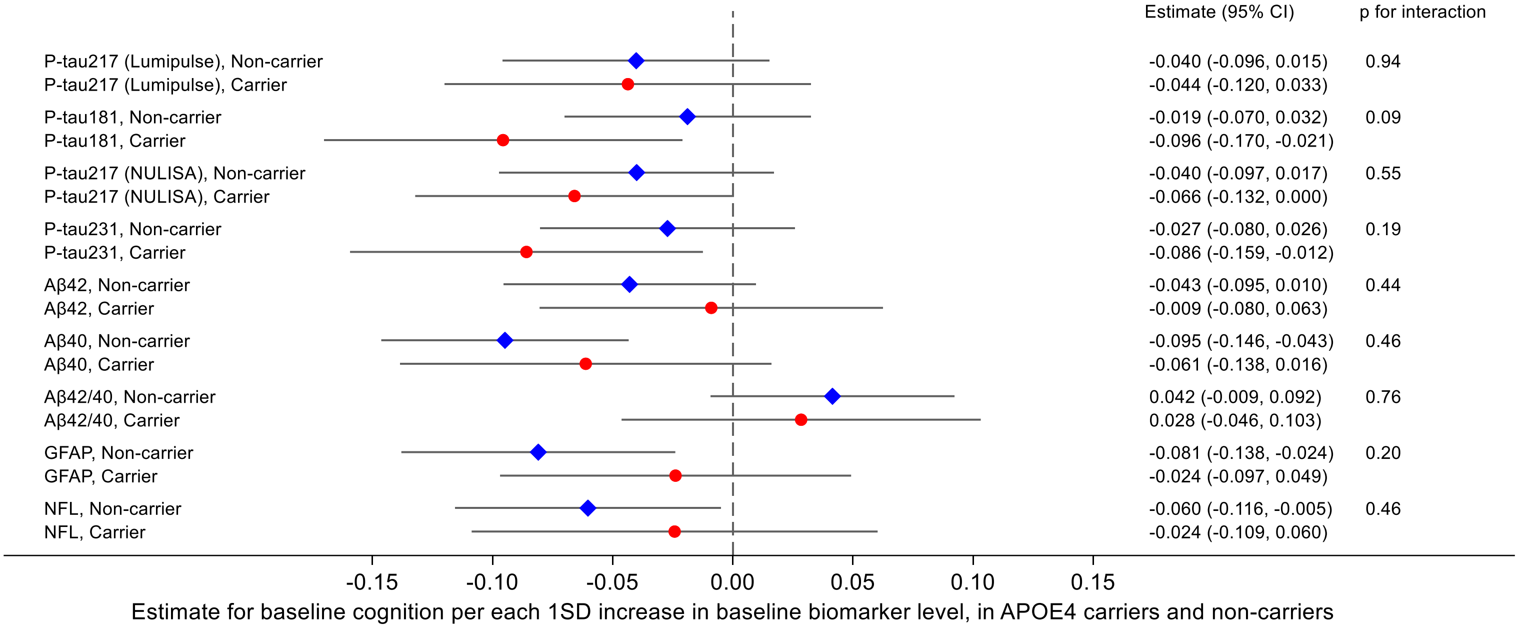

Estimates (95% CI) were obtained from linear regression models including the biomarker**APOE* interaction term, using marginal prediction methods to derive interpretable effect estimates. Results are interpreted as difference in baseline cognitive performance (NTB z-score for global cognition, memory, executive functioning, or processing speed) per 1 SD higher baseline biomarker level, in *APOE*4 carriers (red circle) and non-carriers (blue diamond). Negative estimates indicate worse cognitive performance with increasing baseline biomarker levels. Analyses were adjusted for baseline age, sex, education, study site, BMI, and eGFR. P-values >0.05 indicate that APOE genotype did not significantly modify the associations between baseline biomarker levels and cognitive performance.

Aβ=beta-amyloid; GFAP=glial fibrillary acidic protein; NFL=neurofilament light chain; NTB=neuropsychological test battery; p-tau=phosphorylated tau.

**Figure 7.** Baseline associations between biomarker levels and cognition, by age

**A. P-tau217 (Lumipulse)**

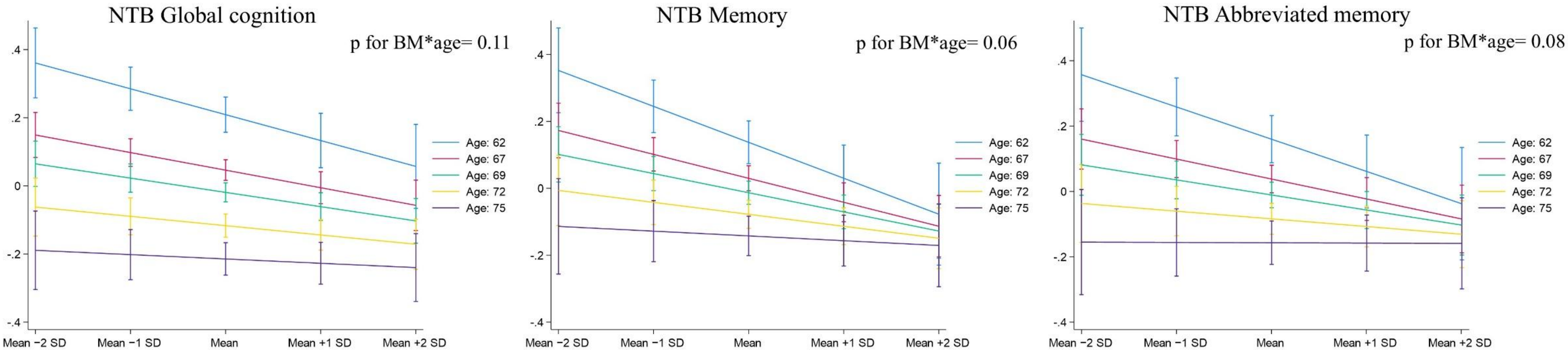

**B. P-tau181**

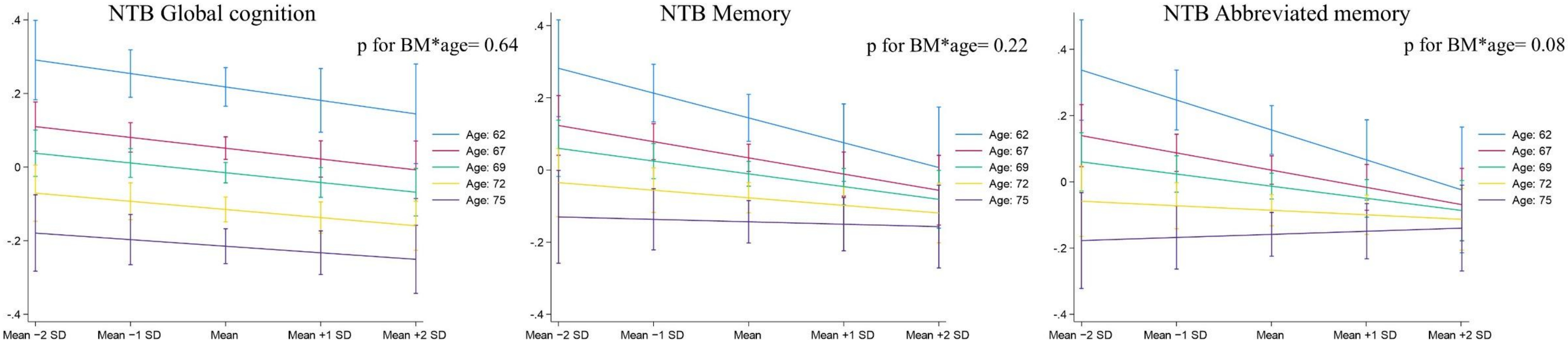

**C. P-tau217 (NULISA)**

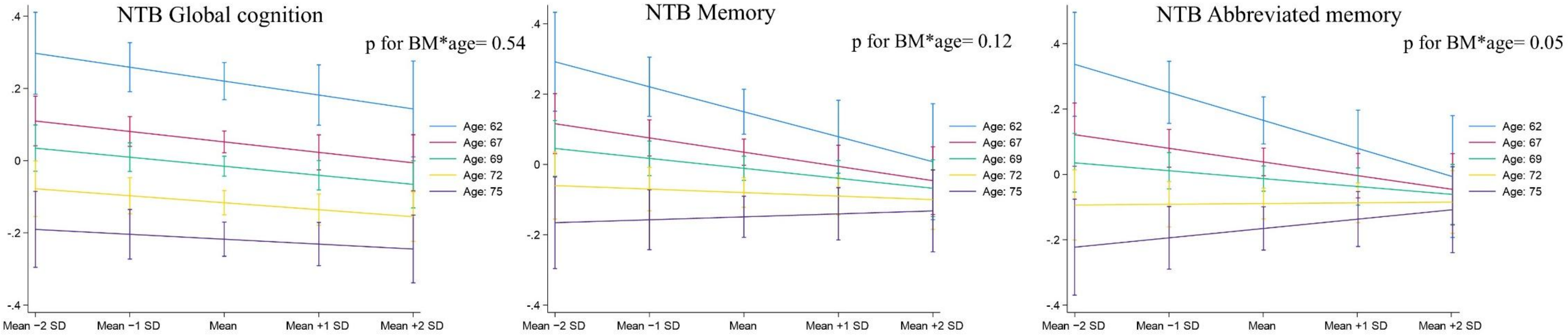

**D. P-tau231**

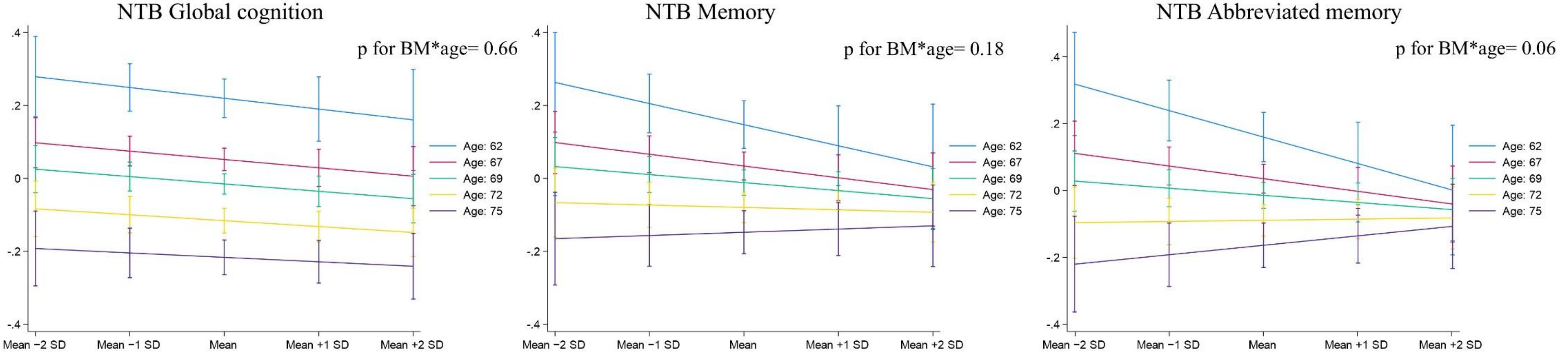

**E. Aβ42**

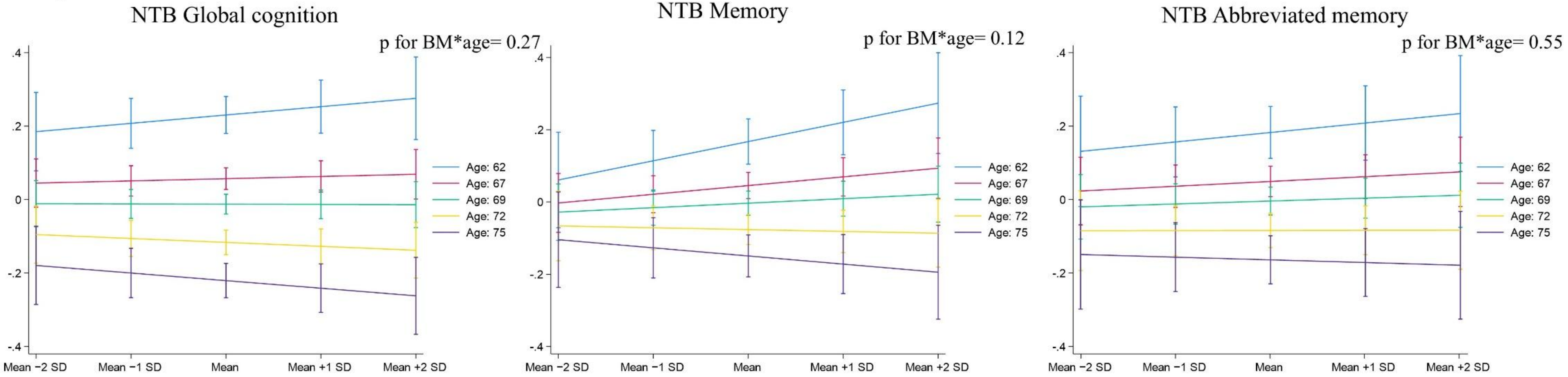
NTB Global cognition
p for BM*age= 0.27
NTB Memory
p for BM*age= 0.12
NTB Abbreviated memory
p for BM*age= 0.55
Age: 62
Age: 67
Age: 69
Age: 72
Age: 75
Mean −2 SD
Mean −1 SD
Mean
Mean +1 SD
Mean +2 SD

**F. Aβ40**

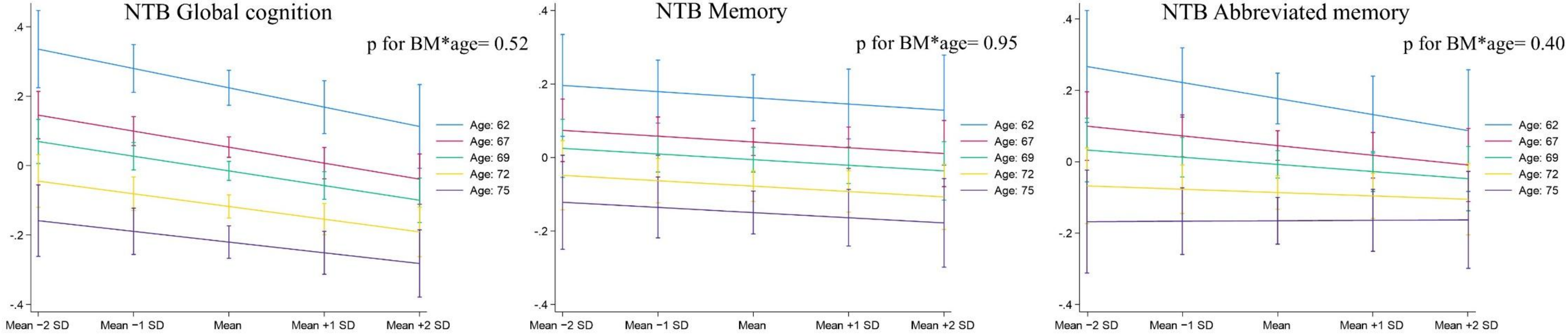
NTB Global cognition
p for BM*age= 0.52
NTB Memory
p for BM*age= 0.95
NTB Abbreviated memory
p for BM*age= 0.40
Age: 62
Age: 67
Age: 69
Age: 72
Age: 75
Mean −2 SD
Mean −1 SD
Mean
Mean +1 SD
Mean +2 SD

**G. Aβ42/40**

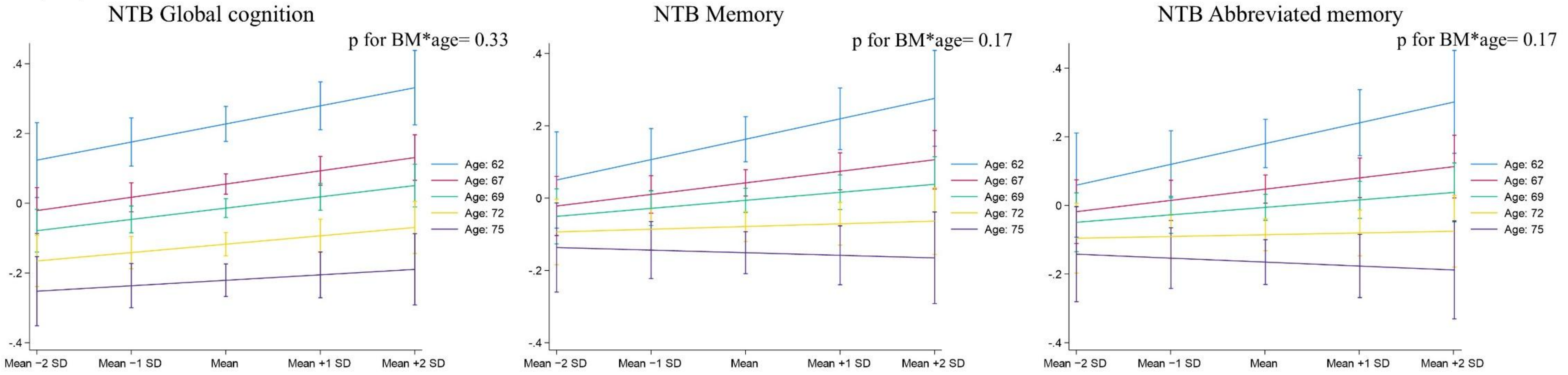

**H. GFAP**

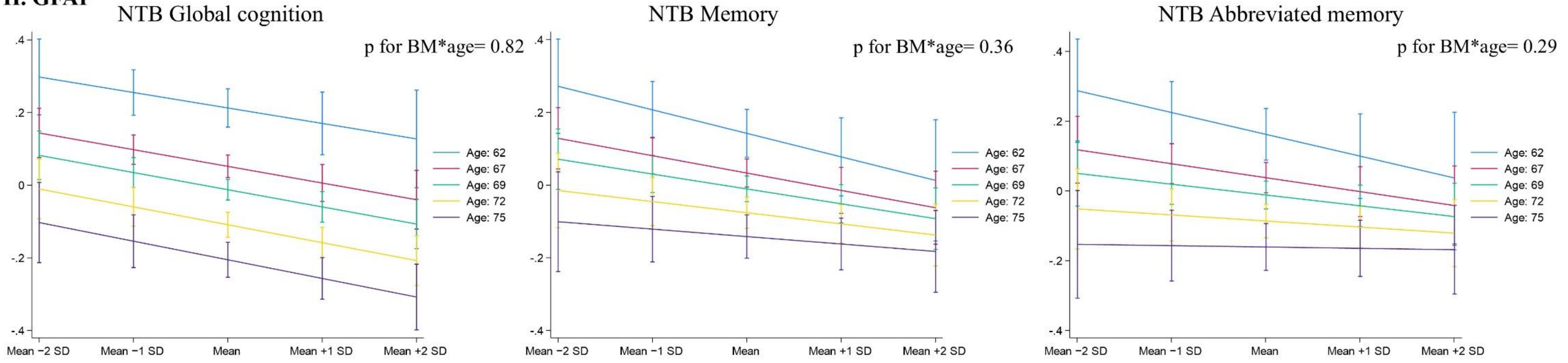

**I. NFL**

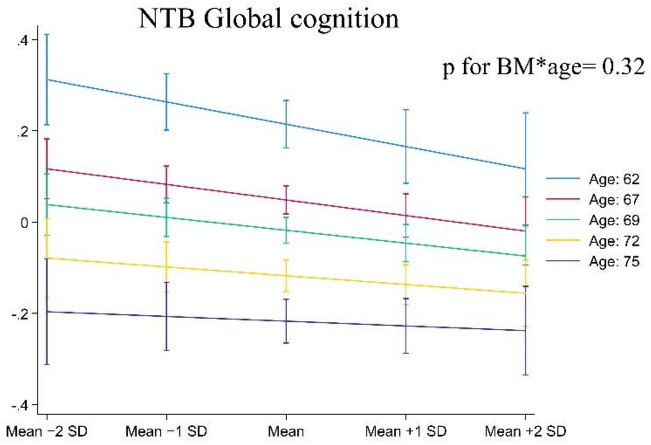

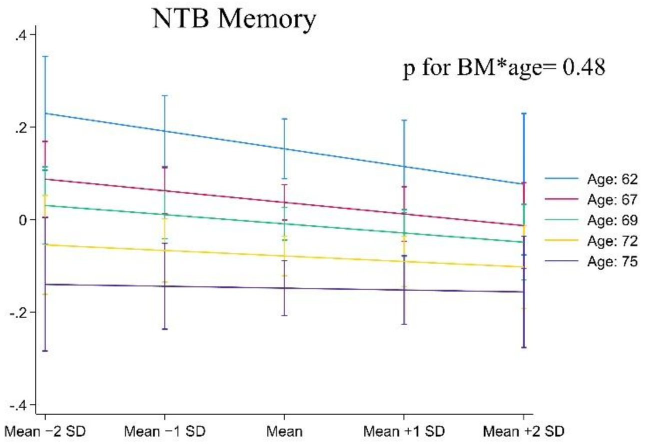

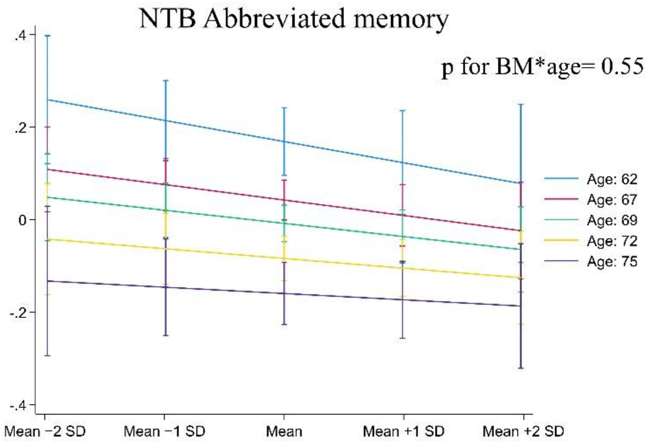

Estimates (95% CI) were obtained from linear regression models including the interaction term biomarker*age, using marginal prediction methods to derive interpretable effect estimates. For visualization, estimated cognitive z-scores (y-axis) are shown for five age groups, corresponding to quintile mean ages, at five baseline biomarker levels, ranging from mean -2SD (lowest) to mean +2SD (highest). A negative slope indicates that increasing baseline biomarker levels were associated with lower cognitive performance at baseline. Analyses were adjusted for sex, education, *APOE*, study site, BMI, and eGFR. P-values >0.05 indicate that age did not significantly modify the associations between baseline biomarker levels and cognitive performance. Results are not shown for executive functioning and processing speed, as no signal was observed for age as an effect modifier (all p-values for interaction >0.2).

Aβ=beta-amyloid; BM=biomarker; GFAP=glial fibrillary acidic protein; NFL=neurofilament light chain; NTB=neuropsychological test battery; p-tau=phosphorylated tau.

**Figure 8.** Baseline associations between biomarker levels and cognition, by level of vascular risk factor burden

**A. P-tau217 (Lumipulse)**

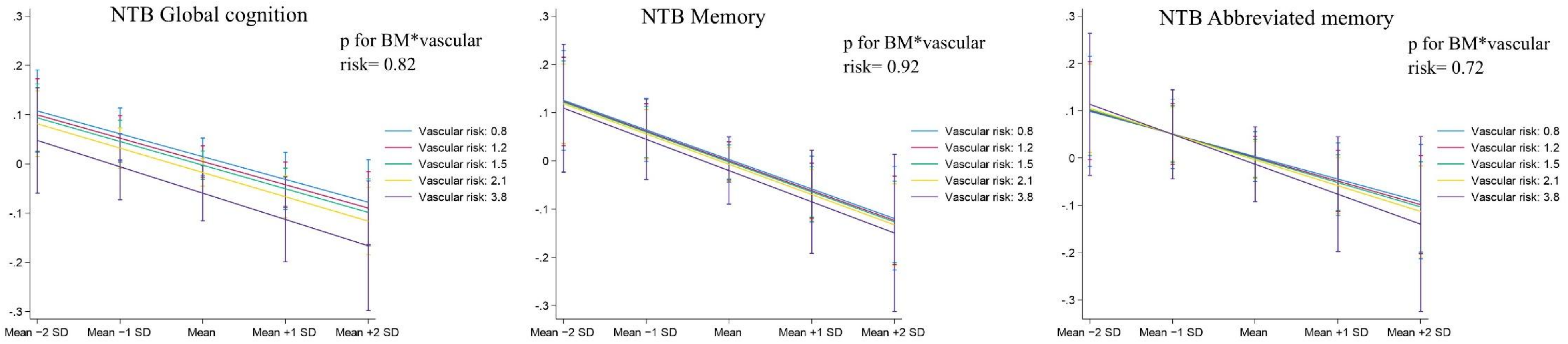

**B. P-tau181**

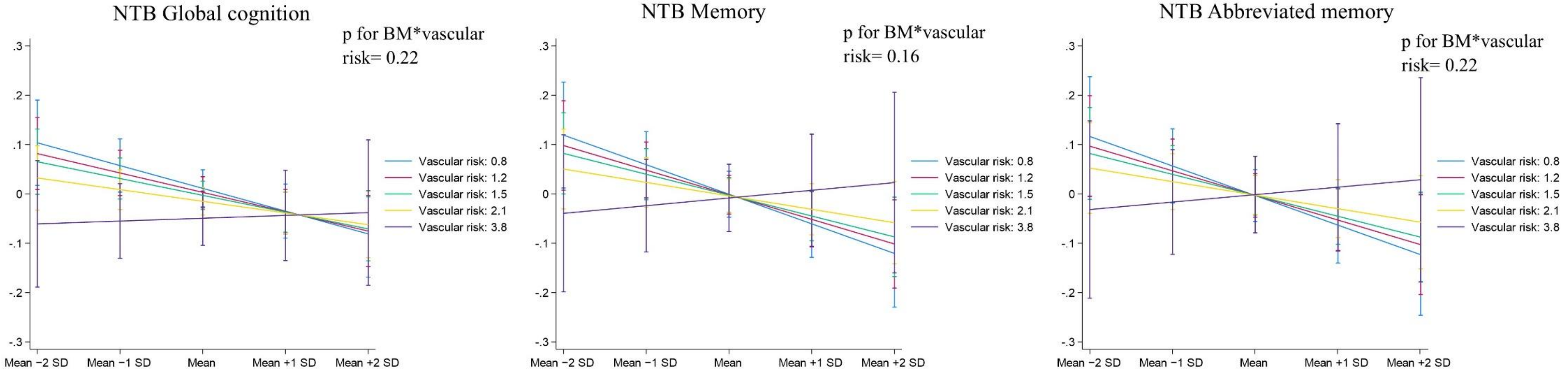

**C. P-tau217 (NULISA)**

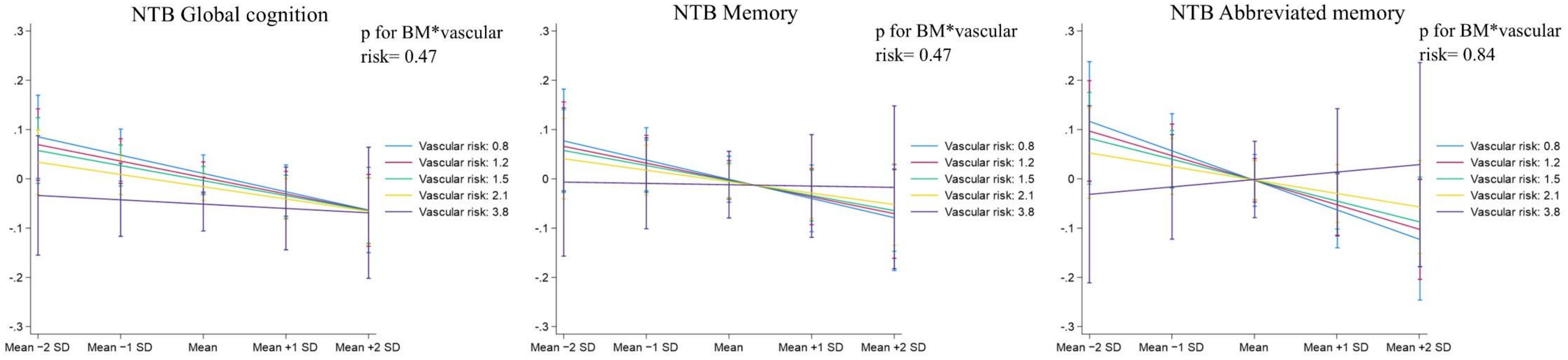

**D. P-tau231**

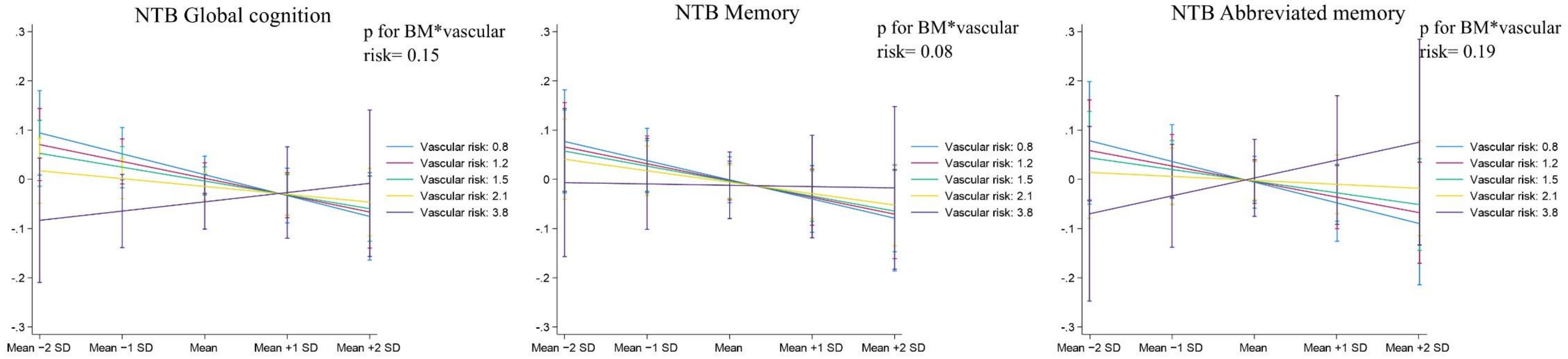

**E. Aβ42**

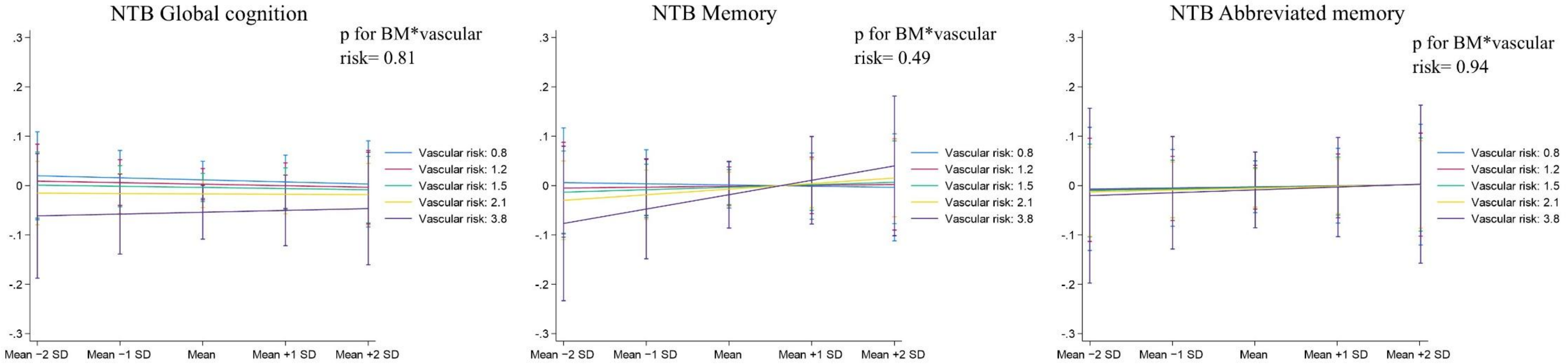
NTB Global cognition
p for BM*vascular risk= 0.81
NTB Memory
p for BM*vascular risk= 0.49
NTB Abbreviated memory
p for BM*vascular risk= 0.94
Vascular risk: 0.8
Vascular risk: 1.2
Vascular risk: 1.5
Vascular risk: 2.1
Vascular risk: 3.8
Mean −2 SD
Mean −1 SD
Mean
Mean +1 SD
Mean +2 SD

**F. Aβ40**

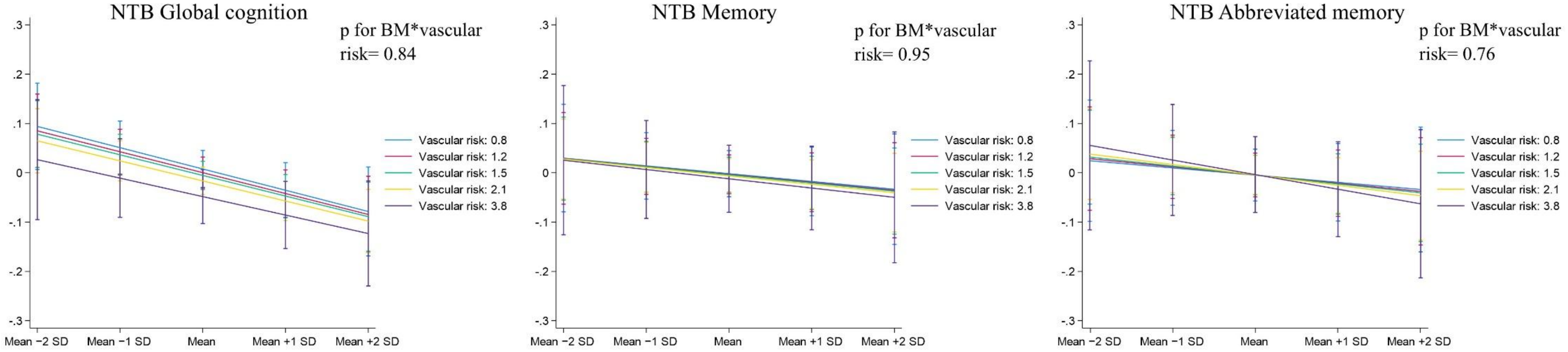
NTB Global cognition
p for BM*vascular risk= 0.84
NTB Memory
p for BM*vascular risk= 0.95
NTB Abbreviated memory
p for BM*vascular risk= 0.76
Vascular risk: 0.8
Vascular risk: 1.2
Vascular risk: 1.5
Vascular risk: 2.1
Vascular risk: 3.8
Mean −2 SD
Mean −1 SD
Mean
Mean +1 SD
Mean +2 SD

**G. Aβ42/40**

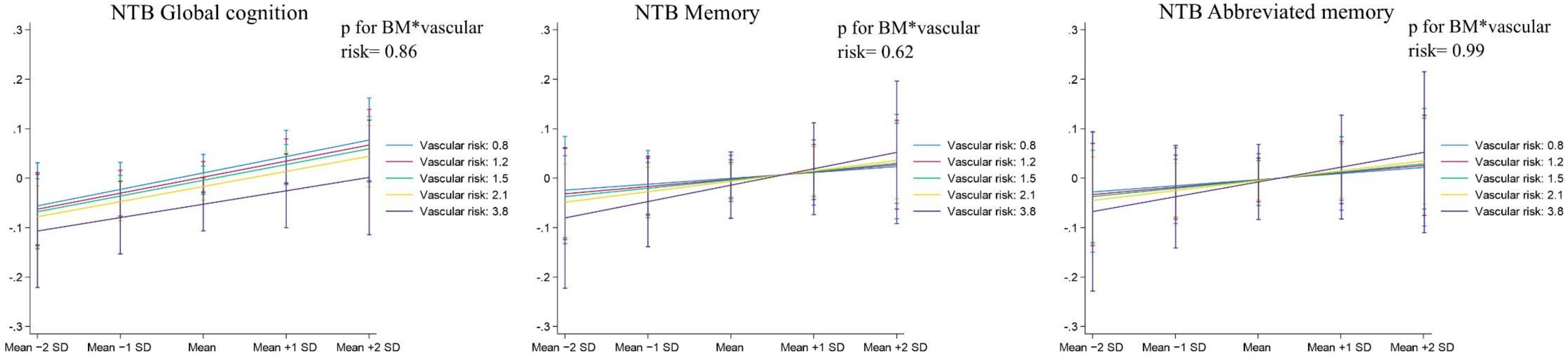

**H. GFAP**

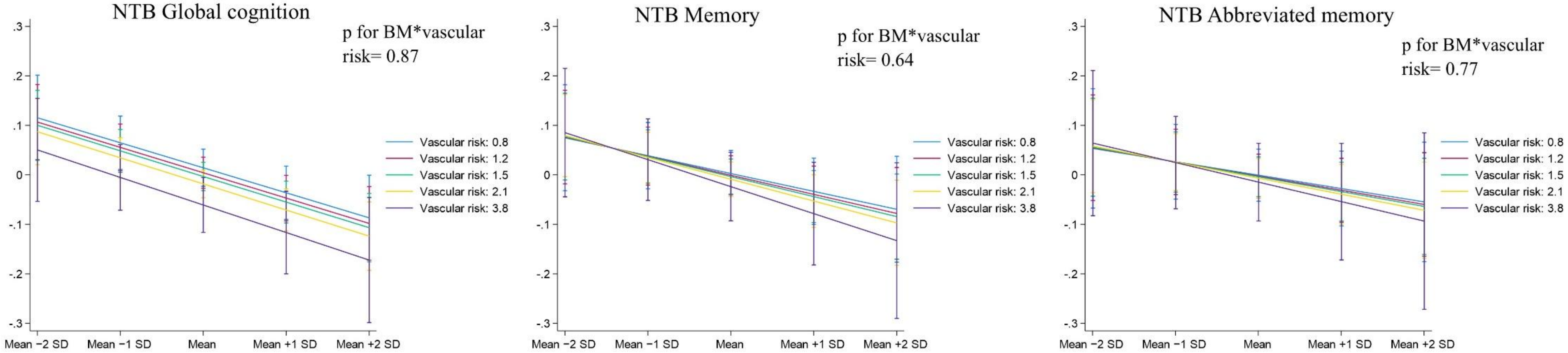

**I. NFL**

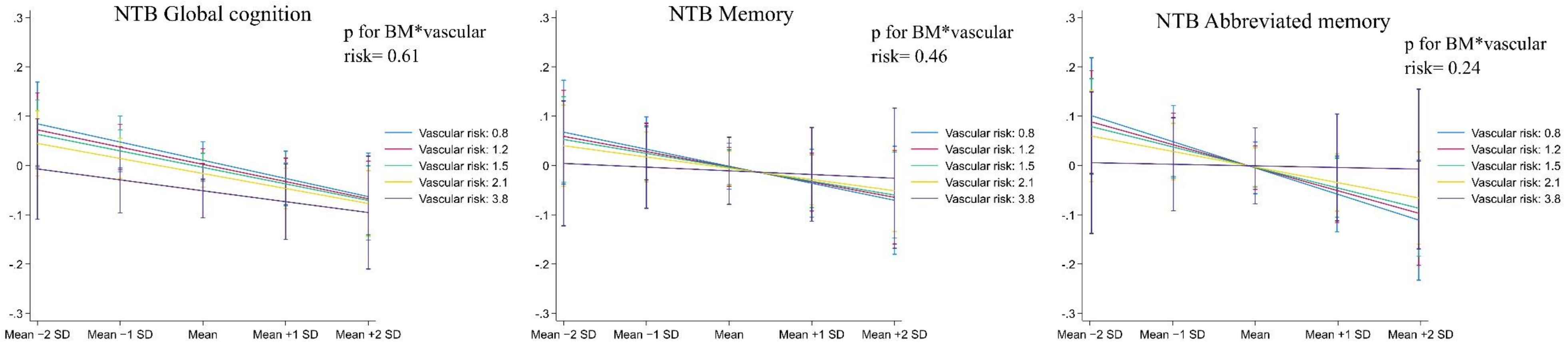

Estimates (95% CI) were obtained from linear regression models including the interaction term biomarker*vascular risk factor burden, using marginal prediction methods to derive interpretable effect estimates. Vascular risk factor burden was assessed using the validated Finnish FINRISK cardiovascular risk score which was divided by the score of age- and sex-matched person without vascular risk factors as done in our previous study (Rosenberg A, Ngandu T, Rusanen M, et al. *Alzheimers Dement*. 2018;14(3):263-270). Higher score indicates higher vascular risk factor burden. For this visualization, estimated cognitive z-scores (y-axis) are shown for five vascular risk groups, corresponding to quintile mean vascular risk scores, at five baseline biomarker levels, ranging from mean -2SD (lowest) to mean +2SD (highest). A negative slope indicates that higher baseline biomarker levels were associated with lower cognitive performance at baseline. Analyses were adjusted for age, sex, education, *APOE*, study site, BMI, and eGFR. P-values >0.05 indicate that vascular risk did not significantly modify the associations between baseline biomarker levels and cognitive performance. Results are not shown for executive functioning and processing speed, as no signal was observed for vascular risk as an effect modifier in these cognitive domains (all p-values for interaction >0.3).

Aβ=beta-amyloid; BM=biomarker; GFAP=glial fibrillary acidic protein; NFL=neurofilament light chain; NTB=neuropsychological test battery; p-tau=phosphorylated tau.

**Figure 9.** Baseline associations between biomarker levels and cognition, by sex

### A. Global cognition

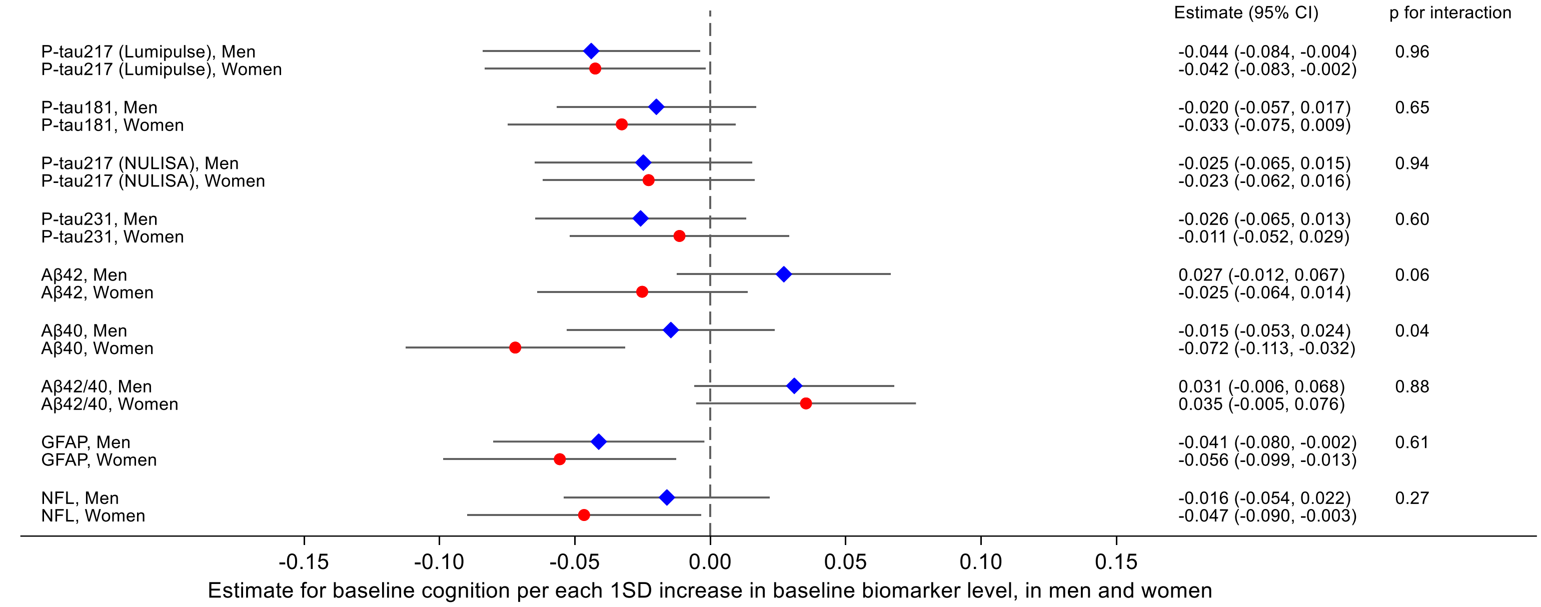

### B. Memory

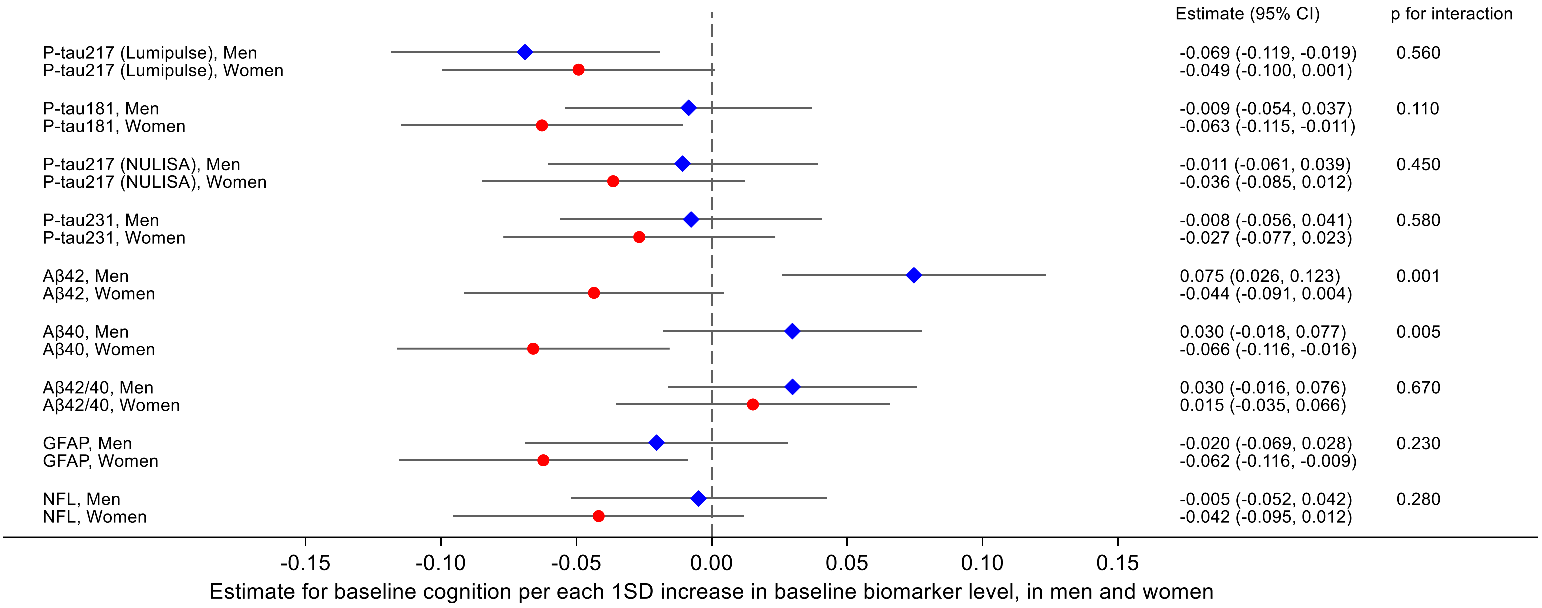

### C. Abbreviated memory

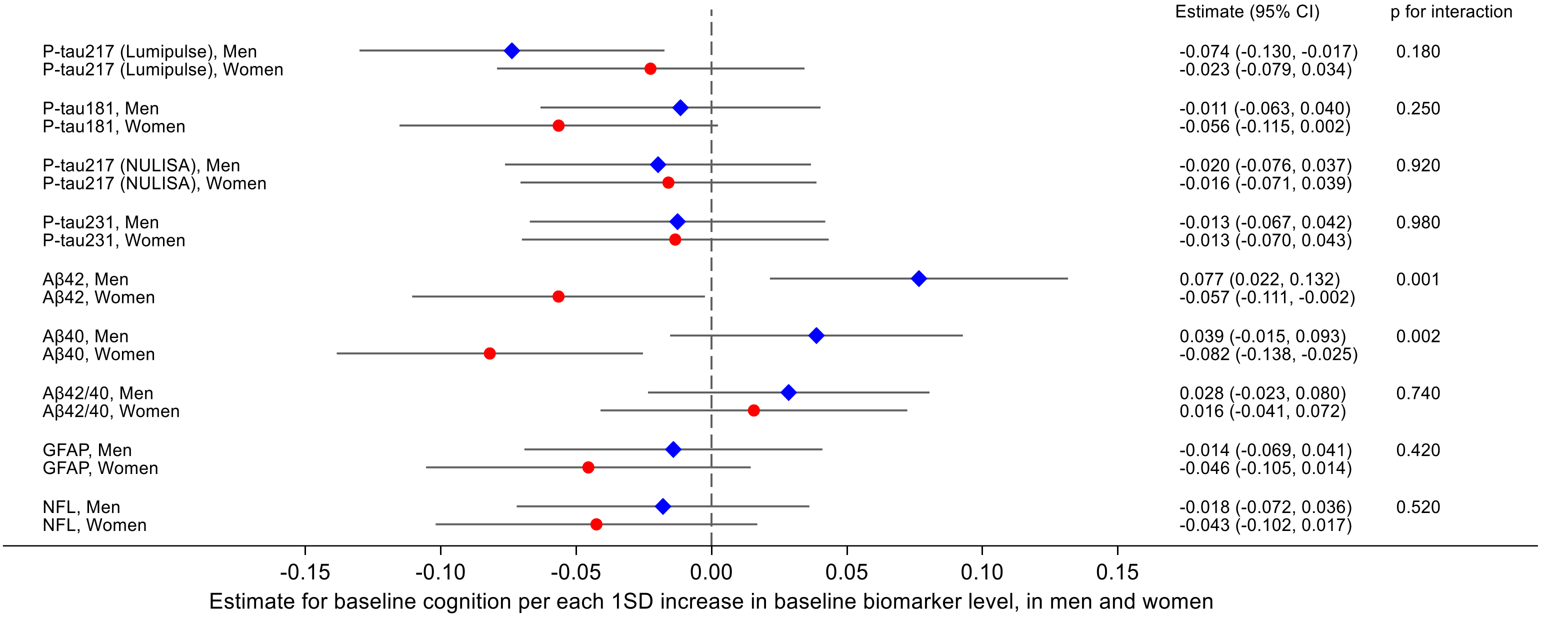

### D. Executive functioning

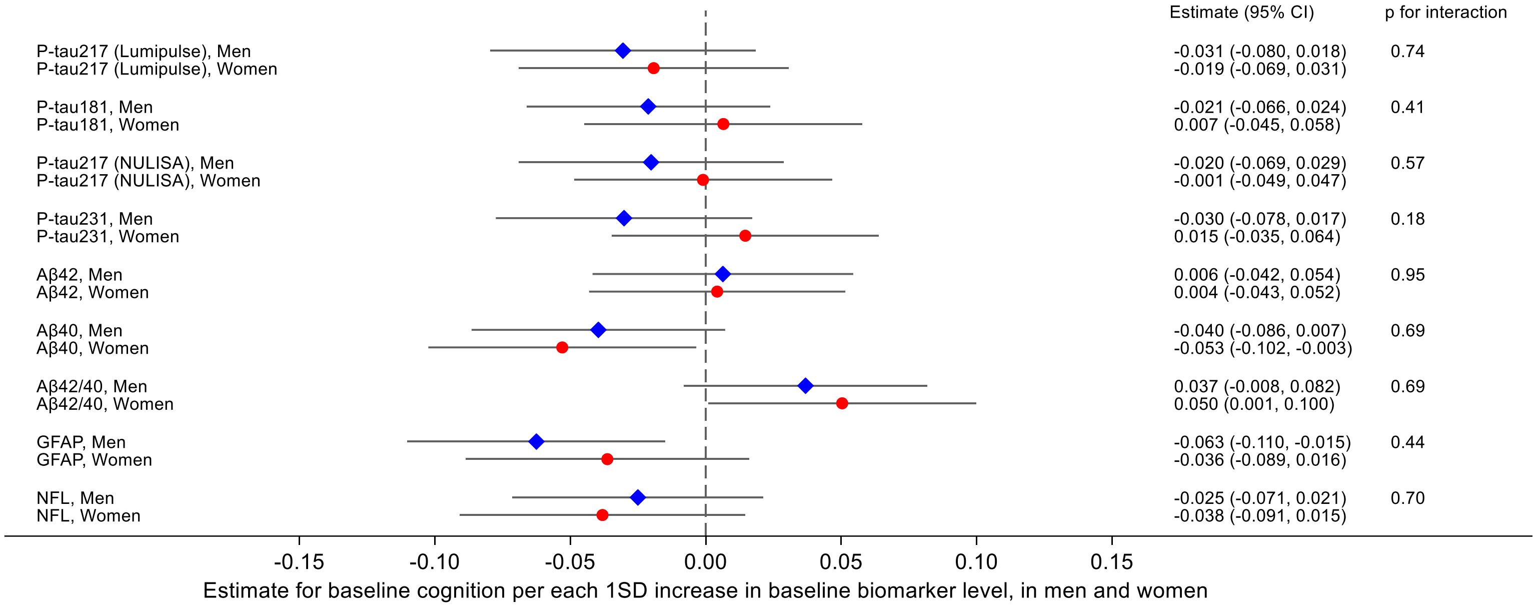

**E. Processing speed**

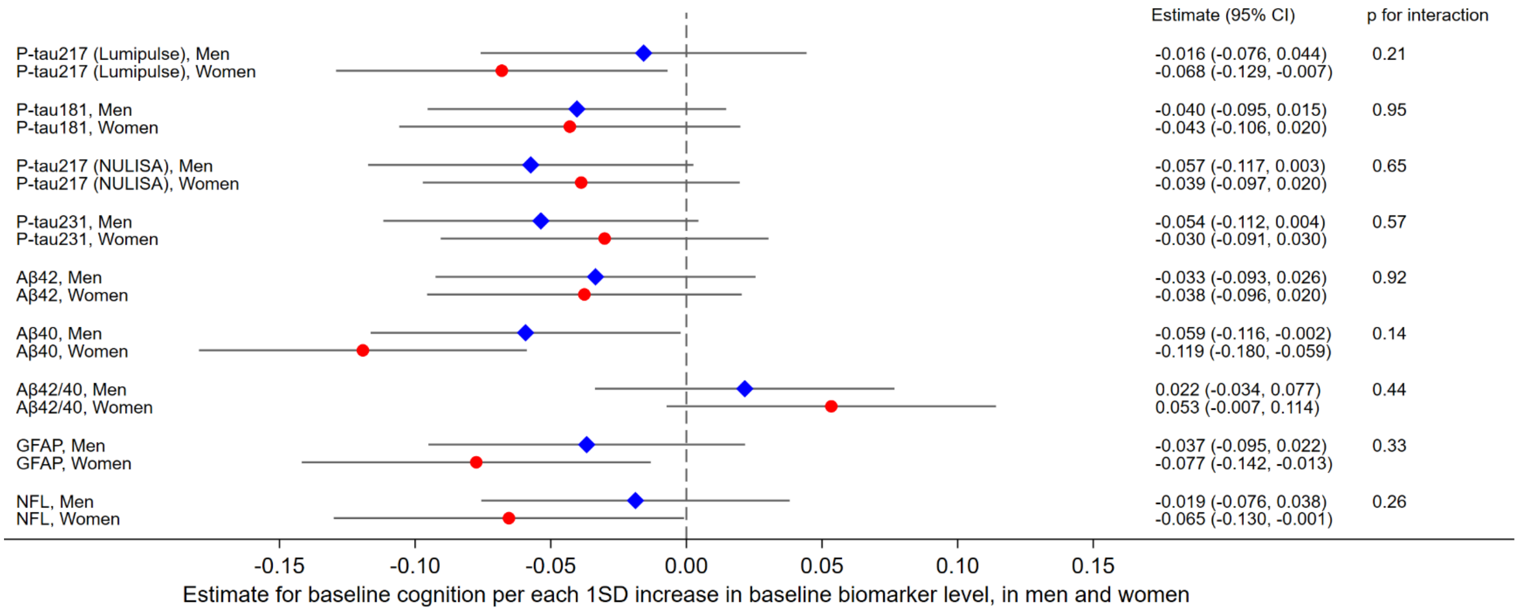

Estimates (95% CI) were obtained from linear regression models including the biomarker*sex interaction term, using marginal prediction methods to derive interpretable effect estimates. Results are interpreted as differences in baseline cognitive performance (NTB z-score for global cognition, memory, executive functioning, or processing speed) per 1 SD higher baseline biomarker level, in women (red circle) and men (blue diamond). Negative estimates indicate worse cognitive performance with increasing baseline biomarker levels. Analyses were adjusted for baseline age, *APOE*, education, study site, BMI, and eGFR. P-values <0.05 indicate that sex modified the association between baseline biomarker levels and cognitive performance.

Aβ=beta-amyloid; GFAP=glial fibrillary acidic protein; NFL=neurofilament light chain; NTB=neuropsychological test battery; p-tau=phosphorylated tau.

**Figure 10.** Baseline biomarker levels and risk of cognitive decline

**A. Global cognition**

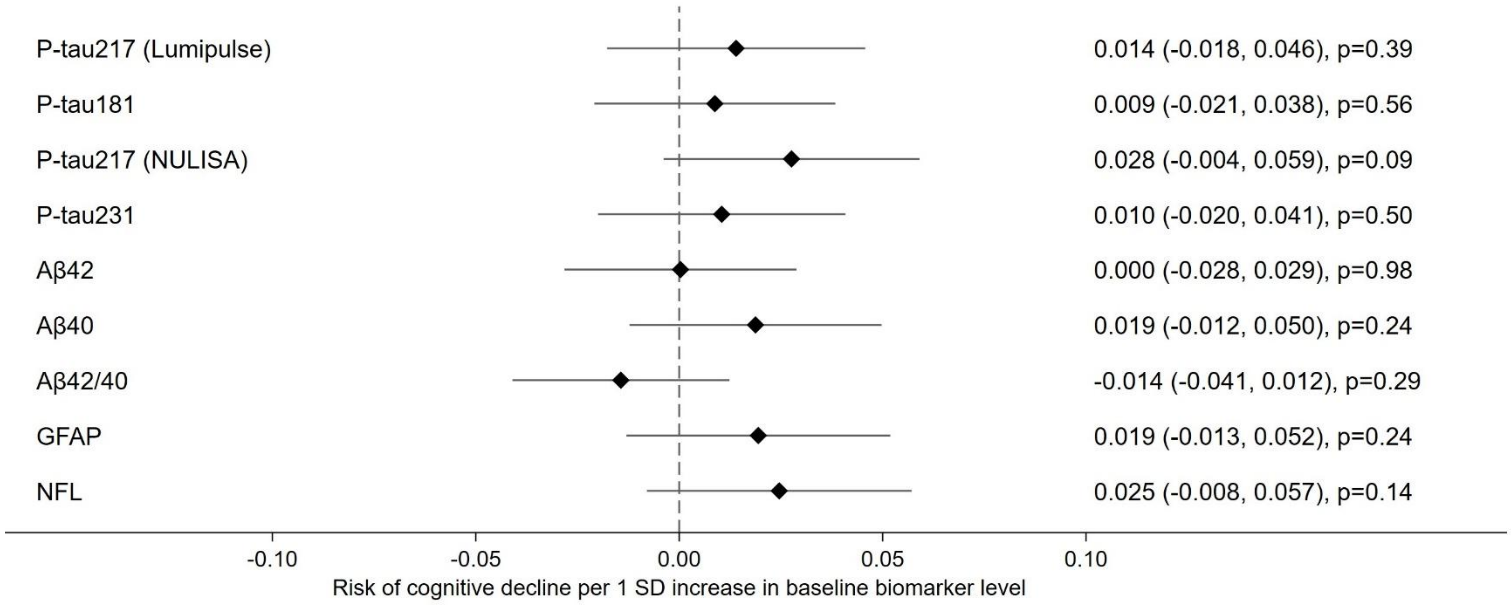

**B. Memory**

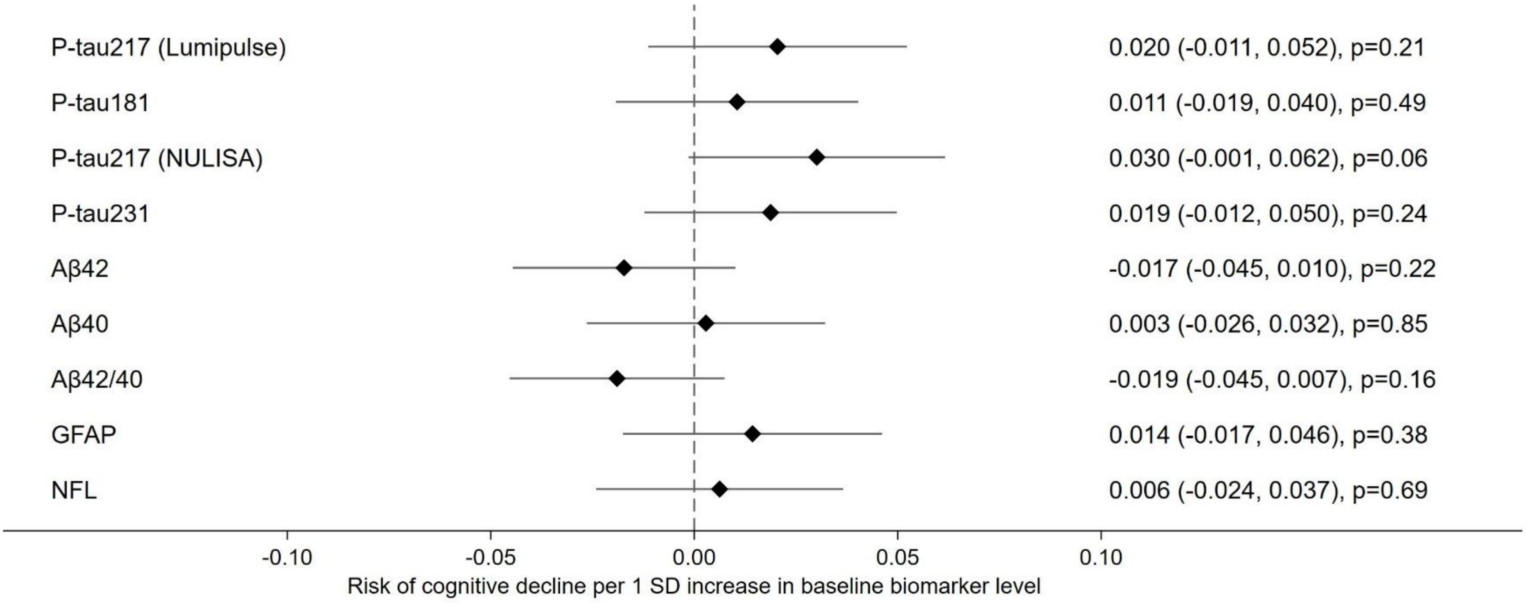

**C. Abbreviated memory**

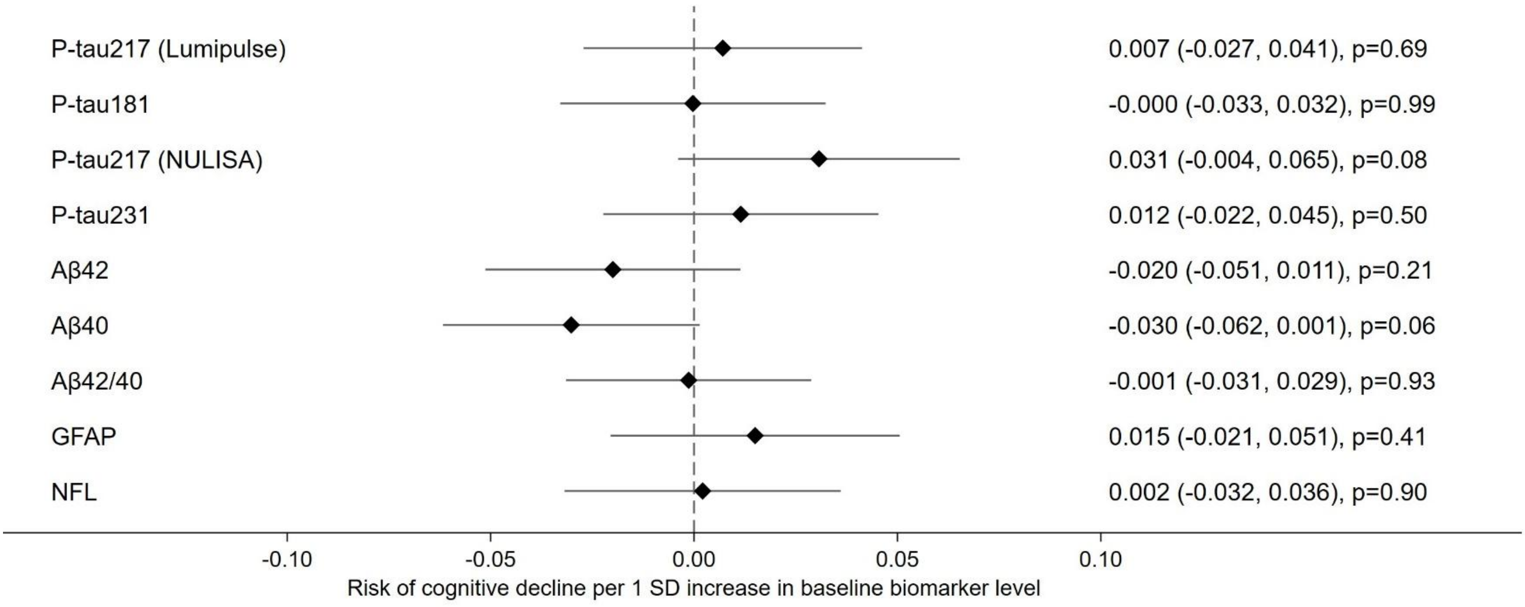

**D. Executive functioning**

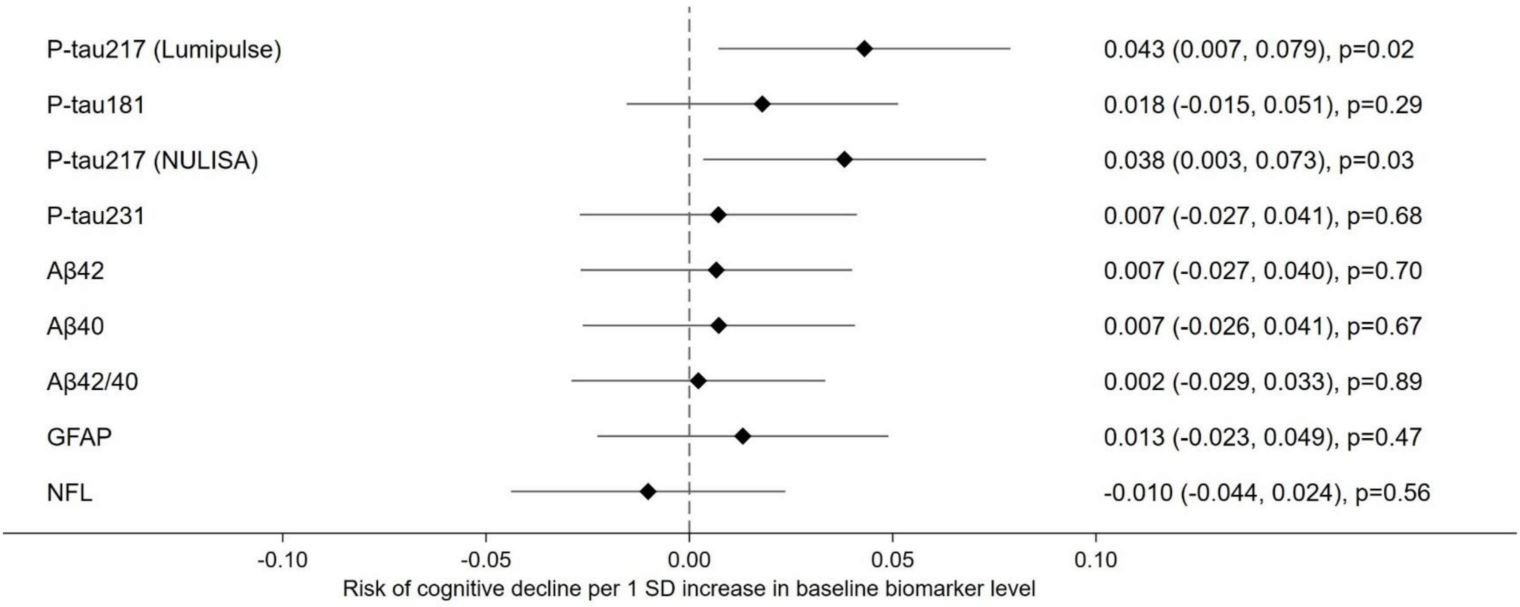

**E. Processing speed**

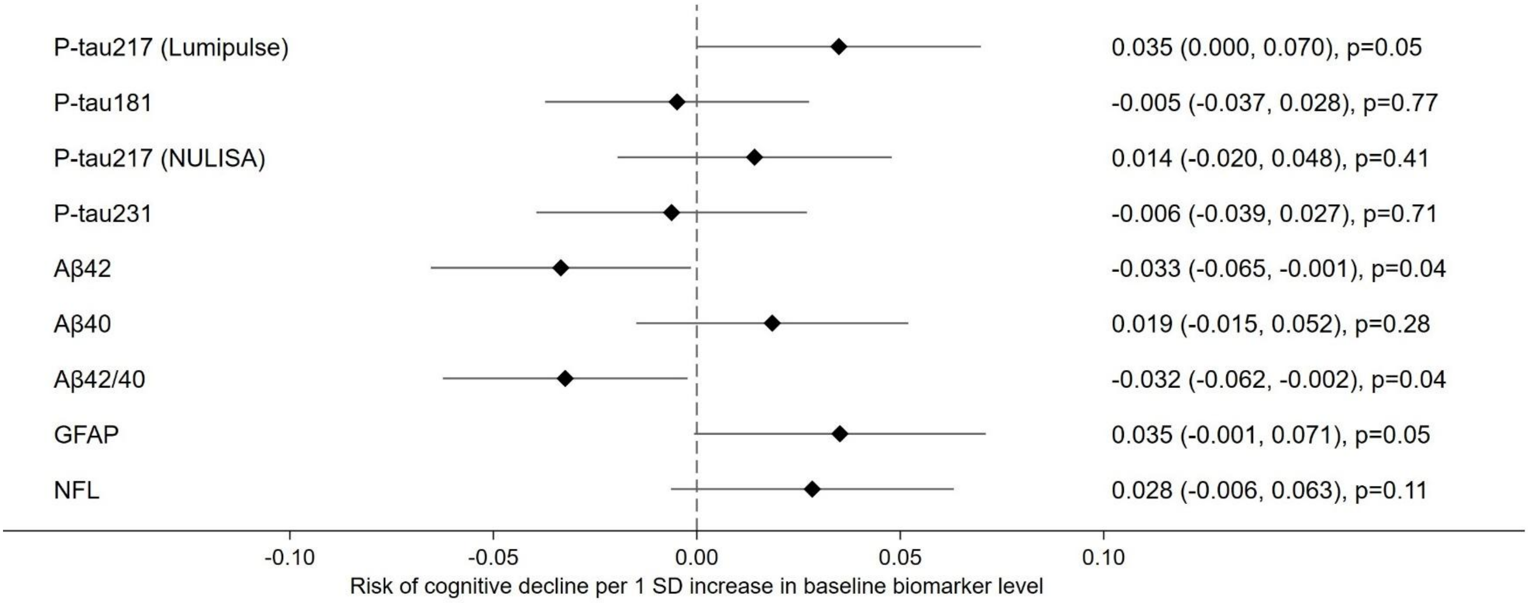

Cognitive decline was defined as a negative Neuropsychological Test Battery (NTB) z-score change from baseline to two years. Estimates (95% CI) were obtained from logistic regression models, using marginal prediction methods to derive interpretable effect estimates. Results are interpreted as differences in the probability of decline, per 1 SD higher baseline biomarker level, in the whole cohort (irrespective of randomization). Positive estimates indicate increased probability, with increasing baseline biomarker levels. E.g., 1SD increase in baseline p-tau217 (Lumipulse) was associated with 4.3% higher risk of experiencing decline in executive functioning (panel D). Analyses were adjusted for baseline cognitive score, age, sex, education, study site, *APOE* genotype, BMI, and eGFR.

Aβ=beta-amyloid; GFAP=glial fibrillary acidic protein; NFL=neurofilament light chain; NTB=neuropsychological test battery; p-tau=phosphorylated tau.

**Figure 11.** Baseline biomarker levels and intervention effects on cognition, by *APOE4* status

### A. Global cognition

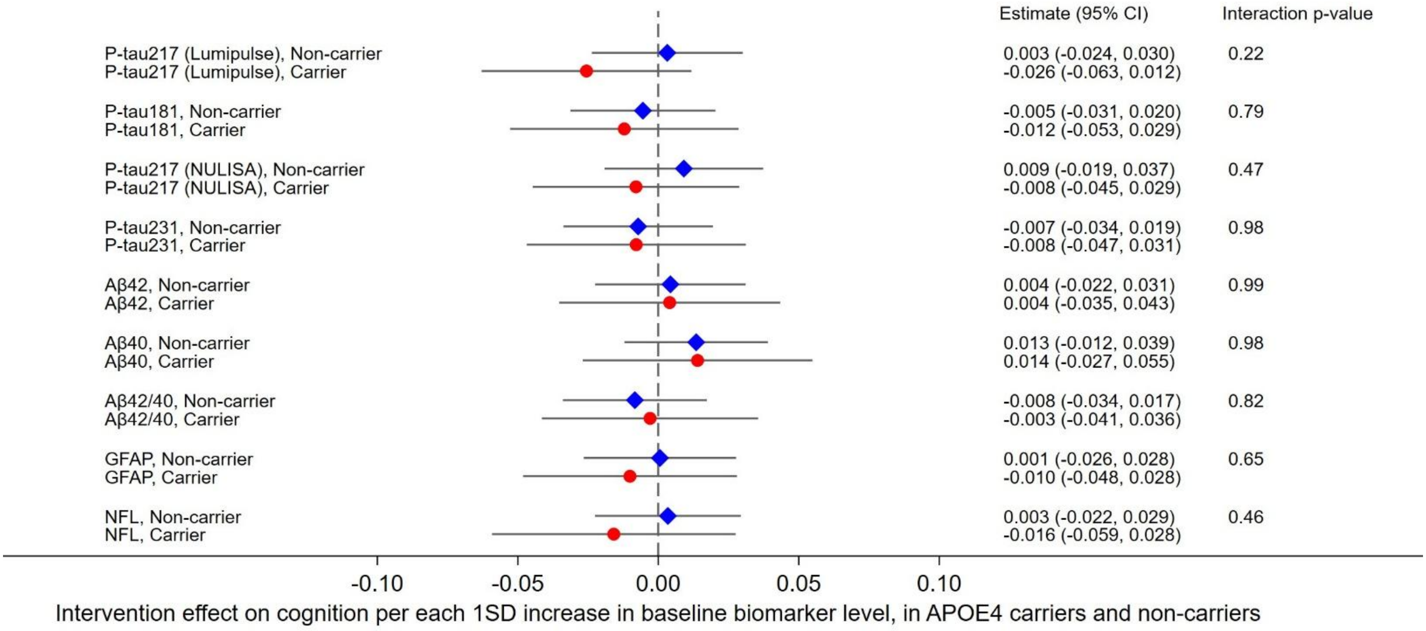

### B. Memory

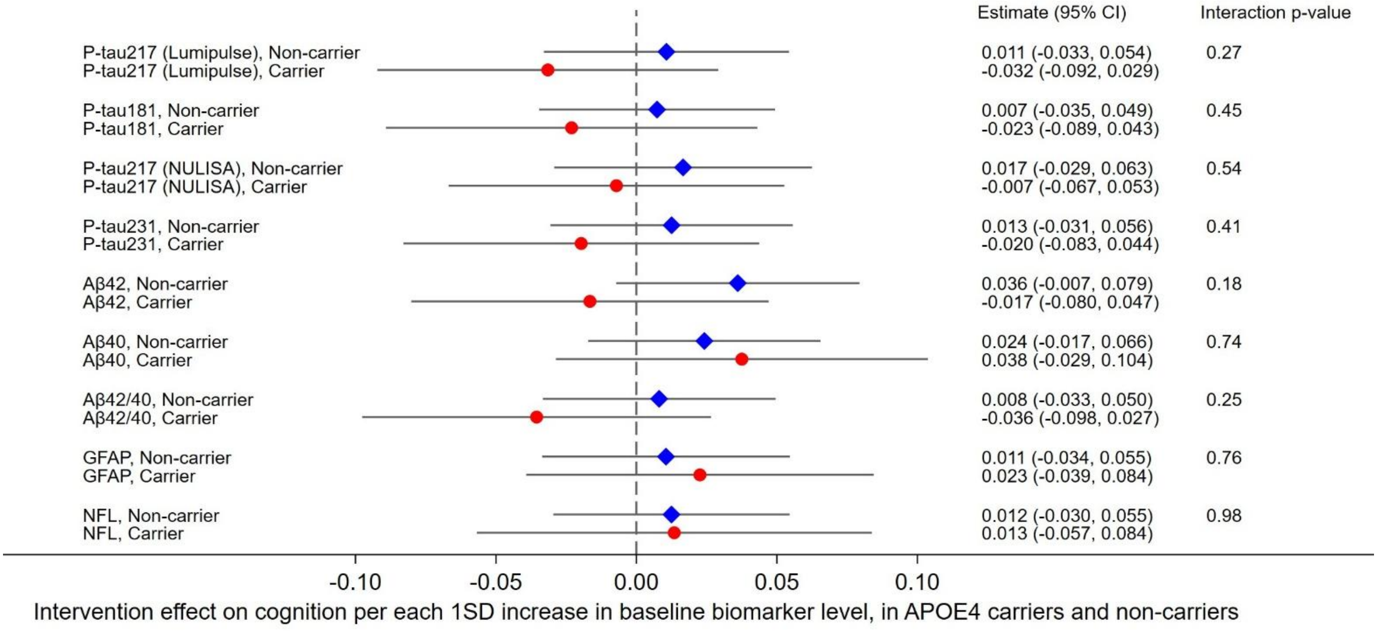

#### C. Abbreviated memory

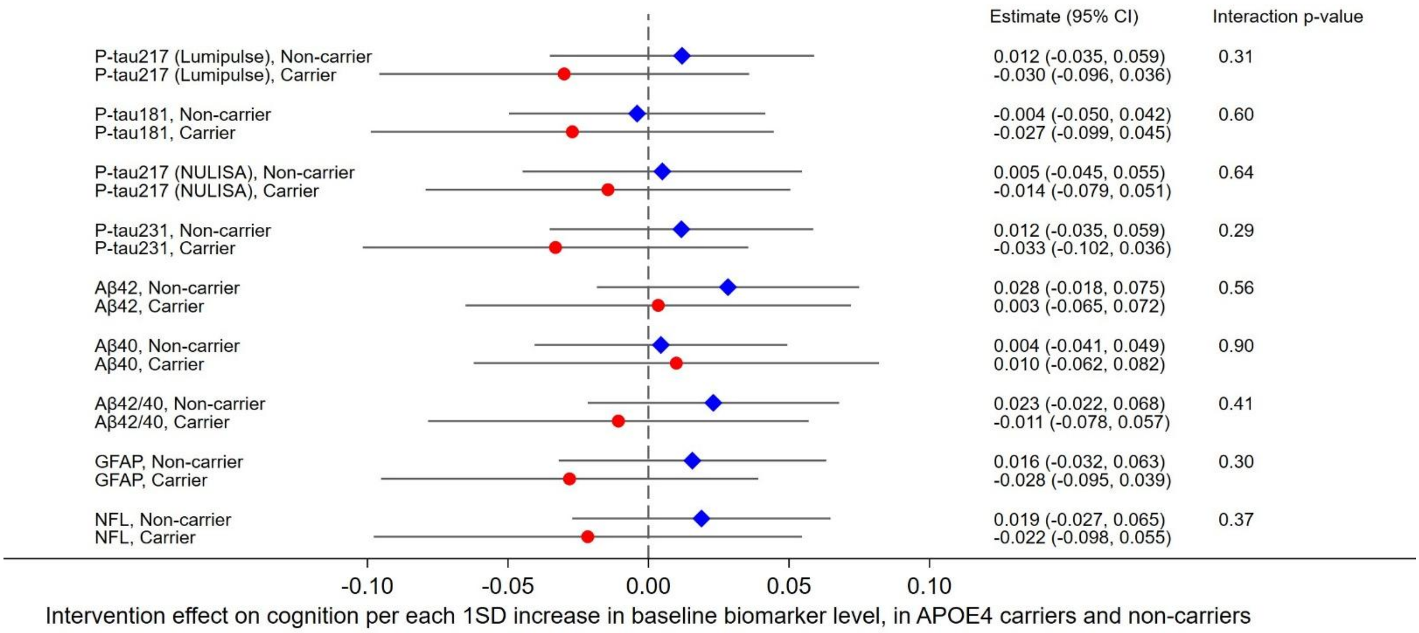

#### D. Executive functioning

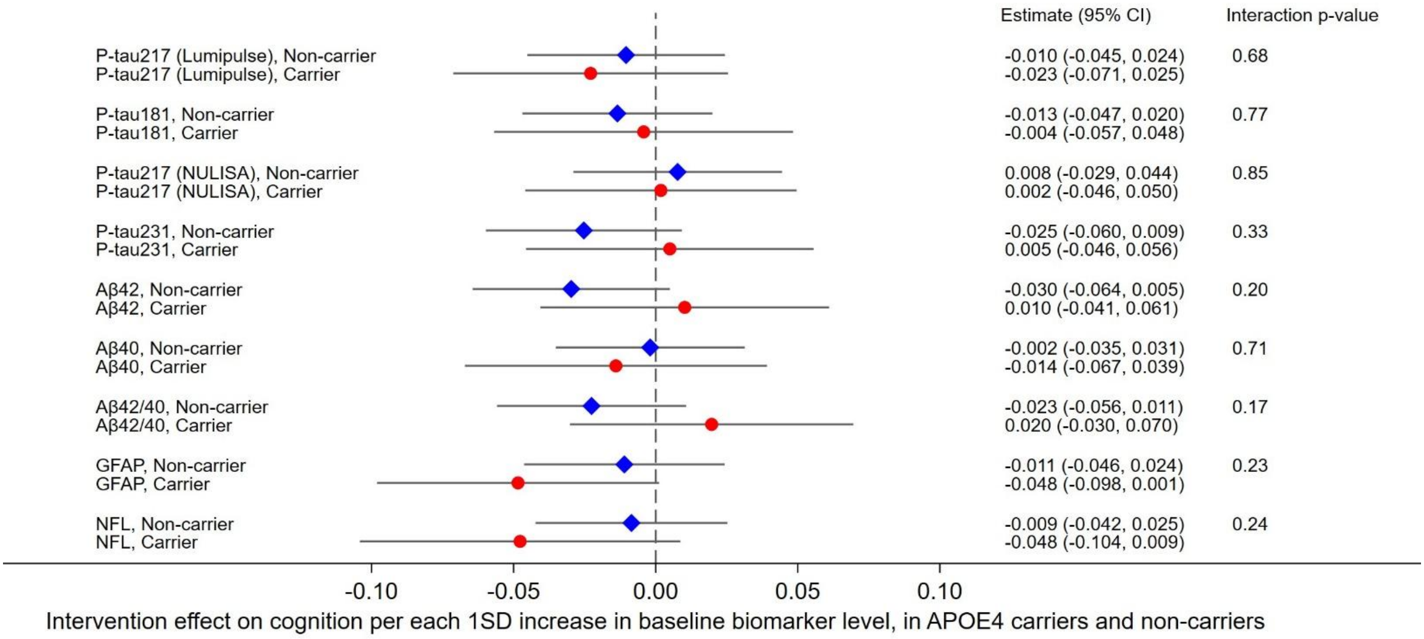

**E. Processing speed**

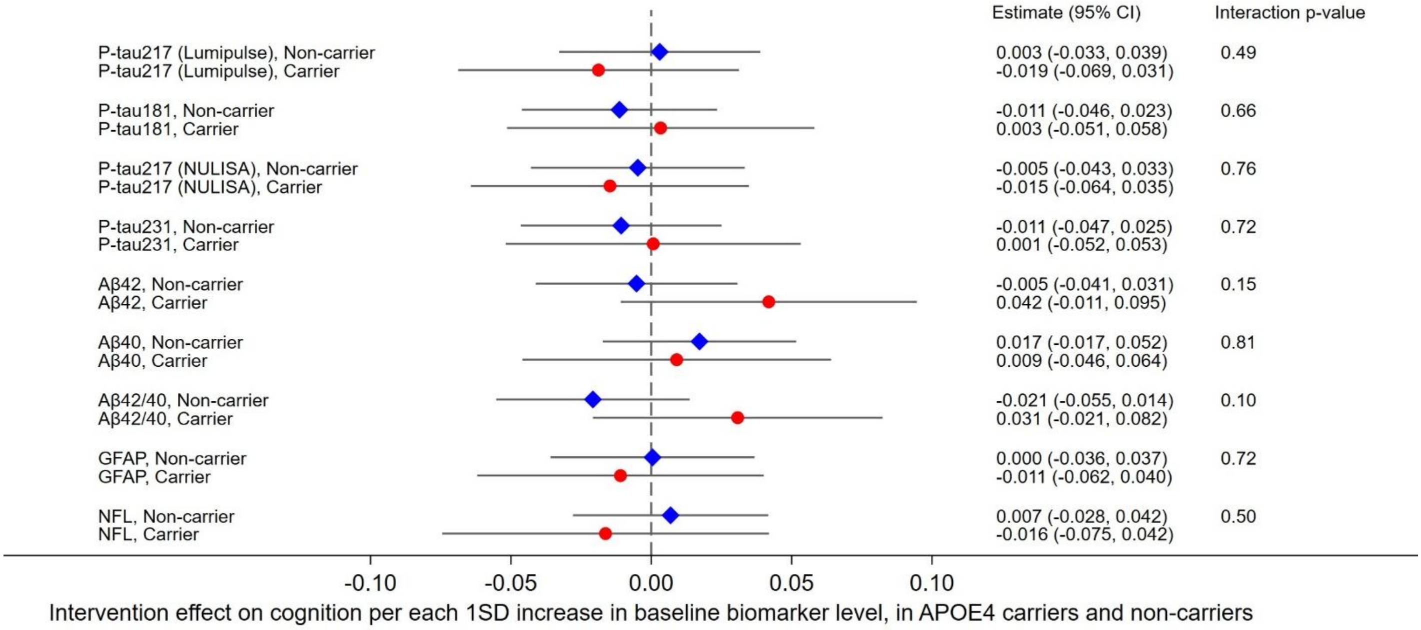

Estimates (95% CI) were obtained from linear mixed-effects models including the 4-way *APOE**biomarker*randomization group*time interaction, using marginal prediction methods to derive interpretable effect estimates. Estimates are interpreted as the impact of each 1SD increase in baseline biomarker level on the intervention effect on cognition, in *APOE4* carriers (red circle) and non-carriers (blue diamond). Positive estimates indicate more pronounced intervention benefit (larger intervention-control group difference in annual cognitive change) with increasing baseline biomarker levels. Negative estimates indicate attenuated intervention benefit with increasing baseline biomarker levels. Analyses were adjusted for baseline age, sex, education, study site, BMI, and eGFR. All *APOE**biomarker*randomization group*time interactions were statistically non-significant p>0.05, indicating that *APOE* genotype did not significantly modify the relationship between baseline biomarker levels and intervention effects on cognition.

Aβ=beta-amyloid; GFAP=glial fibrillary acidic protein; NFL=neurofilament light chain; NTB=neuropsychological test battery; p-tau=phosphorylated tau.
